# The prevalence of probable mental health disorders among hospital healthcare workers during COVID-19: A systematic review and meta-analysis

**DOI:** 10.1101/2022.11.16.22282426

**Authors:** Brian En Chyi Lee, Mathew Ling, Leanne Boyd, Craig Olsson, Jade Sheen

## Abstract

**Objectives:** The mental health impacts of the COVID-19 pandemic continue to be documented worldwide with systematic reviews playing a pivotal role. Here we present updated findings from our systematic review and meta-analysis on the mental health impacts among hospital healthcare workers during COVID-19.

**Methods:** We searched MEDLINE, CINAHL, PsycINFO, Embase and Web Of Science Core Collection between 1st January 2000 to 17^th^ February 2022 for studies using validated methods and reporting on the prevalence of diagnosed or probable mental health disorders in hospital healthcare workers during the COVID-19 pandemic. A meta-analysis of proportions and odds ratio was performed using a random effects model. Heterogeneity was investigated using test of subgroup differences and 95% prediction intervals.

**Results:** The meta-analysis included 401 studies, representing 458 754 participants across 58 countries. Pooled prevalence of depression was 28.5% (95%CI: 26.3-30.7), anxiety was 28.7% (95%CI: 26.5-31.0), PTSD was 25.5% (95%CI: 22.5-28.5), alcohol and substance use disorder was 25.3% (95%CI: 13.3-39.6) and insomnia was 24.4% (95%CI: 19.4-29.9). Prevalence rates were stratified by physicians, nurses, allied health, support staff and healthcare students, which varied considerably. There were significantly higher odds of probable mental health disorders in women, those working in high-risk units and those providing direct care.

**Limitations:** Majority of studies used self-report measures which reflected probable mental health disorders rather than actual diagnosis.

**Conclusions:** These updated findings have enhanced our understanding of at-risk groups working in hospitals. Targeted support and research towards these differences in mental health risks are recommended to mitigate any long-term consequences.

## Introduction

Despite stringent public health measures, the Coronavirus Disease 2019 (COVID-19) pandemic has continued to persist worldwide, with currently more than 600 million people infected and 6.5 million deaths across 233 countries (World Health Organisation, 2021). As different COVID-19 variants (World Health Organisation, 2022) continue to trigger multiple waves of outbreaks worldwide (Hale et al., 2021; Hassan and Mahmoud, 2021), many are concerned that its extended nature and impacts may leave long-lasting mental health consequences (Holmes et al., 2020; Pfefferbaum and North, 2020)

More specifically, there are concerns that hospital healthcare workers’ (HCW) mental health may be disproportionately affected as they continue to work in high-risk settings that are constantly overwhelmed with COVID-19 patients. Studies have shown that working in these settings early on during the pandemic have led to feelings of threat, uncertainty and fatigue among hospital HCWs as they face high infection risks, inadequate protective equipment, inconsistent communications, and increased workload (Digby et al., 2021; Ding et al., 2021; Hennein and Lowe, 2020; Joo and Liu, 2021; Liu et al., 2020). It is thus unsurprising that a number of early reviews have documented a wide range of mental health issues among hospital HCWs during the pandemic, such as depression (Salari et al., 2020b; Yan et al., 2021), anxiety (Raoofi et al., 2021; Salari et al., 2020b; Yan et al., 2021), post-traumatic stress disorder (PTSD)(Yan et al., 2021) and insomnia (Salari et al., 2020a; Salari et al., 2020b; Yan et al., 2021).

In view of these findings and the continuing spread of COVID-19, supporting hospital HCWs’ mental health is of the upmost importance to prevent long lasting consequences that may lead to chronic mental health disorders. To do so effectively, we need to better understand the mental health impacts of the COVID-19 pandemic on hospital HCWs and identify groups that may be at a risk of mental health disorders during the COVID-19 pandemic. This is, however, challenging given that prevalence estimates from previous reviews (Li et al., 2021a; Raoofi et al., 2021; Salari et al., 2020a; Salari et al., 2020b) have varied widely, with some reviews finding similar rates between HCWs and the general public (Cénat et al., 2021; Phiri et al., 2021; Raoofi et al., 2021), making it unclear to what extent working in hospitals during the pandemic have affected hospital HCWs’ mental health. There have also been inconsistent findings on at risk groups, with varied findings on whether mental health issues varies between profession (Olaya et al., 2021; Pappa et al., 2020; Santabárbara et al., 2021; Varghese et al., 2021; Wu et al., 2021), gender (Ching et al., 2021; Li et al., 2021b; Raoofi et al., 2021; Serrano-Ripoll et al., 2020; Varghese et al., 2021; Yuan et al., 2021b; Zhao et al., 2021) and frontline status (frontline HCWs defined as those providing direct care or working in high risk settings) (Bell and Wade, 2020; Carmassi et al., 2020; De Brier et al., 2020; Galli et al., 2020; Muller et al., 2020). The dynamic progression of the COVID-19 pandemic has also made it more important to consistently update findings and provide understanding on longitudinal trends to provide effective recommendations. Particularly for hospital HCWs, given that recent impacts of COVID-19 on hospitals have likely put them at high risk of mental health disorders, targeted recommendations are important to help focus and design effective interventions and strategies to protect their mental health.

## Aims and objectives

This systematic review and meta-analysis aim to provide updated findings and prevalence estimates on current mental health disorders experienced by all hospital HCWs and each professional group, as well as investigating whether prevalence of mental health disorders differed by gender and frontline status.

## Methods

This systematic review followed the Preferred Reporting Items for Systematic Review and Meta-Analysis (PRISMA) guidelines when conducting this study (Moher et al., 2009) (S1 Appendix). The protocol for this systematic review and meta-analysis was pre-registered and submitted to PROSPERO (ID:CRD42020219174). Due to the rapid publication of studies, the research questions and systematic review went through several iterations (detailed in PROSPERO) to ensure its relevance and avoid duplication of efforts.

### Search strategy

A systematic search of the following database was conducted: MEDLINE, CINAHL, PsycINFO via EBSCOhost, Embase via Emabase.com, and Web Of Science Core Collection via webofknowledge.com. Searches were restricted to peer review articles, from 1st January 2000 to 17^th^ February 2022, and English publications.

Search terms were derived from previous reviews and in consultation with a specialist librarian using the PRESS checklist (McGowan et al., 2016). Key concepts included healthcare workers, mental health, and coronavirus. Related terms, database specific MESH terms, truncation and proximity commands were used in the search. (A sample search strategy is shown in S2 Appendix). Endnote X9 was used to retrieve searches and remove duplicated citations.

### Inclusion and exclusion criteria

Prior to any screening, inclusion and exclusion criteria were pre-defined. Studies were included if they: (1) reported prevalence rates of any outcome related a mental health disorder defined by the diagnostic and statistical manual of mental disorders 5^th^ edition (DSM-V)(American Psychiatric Association, 2013), (2) reported data on any HCWs who have worked in a tertiary or secondary level hospital during the COVID-19 pandemic, (3) and used validated methods (self-report measurement tools or diagnostic interviews) to identify the presence of a mental health disorder. Studies were excluded if they: (1) were not published in the English language, (2) were not a primary study, (3) was published as an editorial, communication or brief report, (4) full-text were unobtainable (5) did not report the use of valid cut offs or scoring methods or (6) were not peer-reviewed.

### Operational definition-Presence of a mental health disorder

The presence of a mental health disorder in this systematic review is defined as those meeting the validated criteria on diagnostic interviews for mental health disorders as defined by the DSM-V(American Psychiatric Association, 2013). Where diagnostic interviews are not used, the presence of clinically significant symptoms that are likely to indicate a probable mental health disorder will be applied instead. This is defined as those meeting optimal cut-offs on measurement tools that have been validated against diagnostic interviews and using validated methods when estimating the likelihood of mental health disorder present in an individual.

### Study selection

The first author BL screened all studies’ title and abstract using the pre-defined criteria. When title and abstract were not clear, studies were reviewed at full text by BL. A random 10% of studies at the title and abstract stage and full text stage were screened independently by a second reviewer from the research team (DG, HS, CH, AJ, or BD). Conflicts were resolved by an independent third reviewer. After resolving conflicts, the first author’s inter-rater agreement with second reviewers was at 98.7% at the title and abstract and 98% at the full text stage.

### Data extraction

Following full-text screening, BL independently completed the risk of bias assessment and data extraction. Data extracted from the included studies are as follows: (1) author details, (2) study design, (3) coronavirus outbreak setting, (4) outcomes measured, (5) measurement tools, (6) recruitment dates, (7) mean or median age, (8) country of study, (9) overall sample size of hospital HCWs, (10) subgroup sample size (i.e., physicians, nurses, allied health staff, support staff, non-medical staff, healthcare students, hospital HCWs working in high risk or low risk units, hospital HCWs providing direct care or do not provide direct care, male and female), and the number of hospital HCWs overall and in subgroups who meet the criteria for a mental health disorder. Total and stratified prevalence rates were calculated as number of sample meeting criteria for a mental health disorder divided by total sample size. When only percentage was reported for prevalence rates, the number of participants with a mental health disorder was recalculated by multiplying point prevalence rates with total sample size. When multiple reports of the same dataset were found, they were merged and reported as one dataset to avoid double counting studies. When studies used both interviews and self-report measures, only prevalence data measured by interviews were extracted for analysis due to their increased validity and reliability. When studies reported incomplete or inconsistent data, authors were contacted for more information. All data extraction went through a second check by a second reviewer.

### Quality assessment

The checklist for prevalence studies (Munn et al., 2015) from the Joanna Briggs institute (JBI) was used to assess a study’s risk of bias to sampling, precision, reporting, reliability, validity, response, and coverage (S2B Appendix). When there were disagreements, they were resolved through discussions between reviewers until there was a consensus.

### Data synthesis

A meta-analysis of proportions was conducted using R (R Core Team, 2013), and the packages meta, metafor and dmetar (Balduzzi S et al., 2019; Harrer et al., 2019a; Harrer et al., 2019b; Viechtbauer, 2010; Wang, 2018). Due to the variability between studies, it is assumed that there is no one true effect size and thus a random effects model, using the restricted maximum likelihood method (REML) was used to pool prevalence rates (Borenstein and Higgins, 2013). Prevalence rates were transformed using the Freeman-Tukey Double arcsine transformation and back transformed for ease of interpretation, as recommended by Barendregt et al (Barendregt et al., 2013) to address, if any, imbalanced weighting from extreme estimates and non-normal distribution of prevalence rates in the meta-analysis. The variability in prevalence estimates that was due to heterogeneity was assessed using standard *χ*^2^ to test for significant heterogeneity and *I*^2^ to determine low (< 25%), moderate (25%-50%), and high (> 75%) heterogeneity among studies (Higgins et al., 2003).

### Subgroup analyses

#### Subgroup analyses by study level characteristics

Subgroup analyses were conducted on overall pooled prevalence rates when there were four or more studies (Fu et al., 2011), with test of subgroup differences using *Q* tests for categorical predictors (i.e. measurement tools, region and risk of bias). Test of subgroup difference between countries was considered inappropriate as there was insufficient studies (less than 4 studies) in the majority of countries. As such, the studies were categorised post-hoc according to regions categorised by the World Health Organisation regions (i.e., Africa, Americas, South-East Asia, Europe, Eastern Mediterranean, Western Pacific), as these regions have been shown to have significantly different prevalence of mental health issues when estimated for the general public (Nochaiwong et al., 2021).

#### Subgroup analyses by sub-populations

Stratified prevalence rates were estimated in individual professions to compare differences between prevalence rates among the different professions. To avoid false positives common in subgroup analyses where there is high heterogeneity (Higgins and Thompson, 2004), as well as to quantify the effect of working in high-risk units, providing direct care to COVID-19 patients and gender on prevalence rates, pooled odds ratios (OR) were estimated for these sub-populations. Studies were only included in the OR analyses if they reported prevalence rates for both groups of interest (e.g., prevalence rates for both male and females). To estimate pooled ORs, log odds ratios were calculated and derived from prevalence rates reported by individual studies, pooled using a random effects model (REML method) and then back transformed for ease of interpretation (Borenstein and Higgins, 2013). To reduce type 1 error from having low numbers of studies and heterogenous findings, the Hartung-Knapp method was used to estimate confidence intervals for ORs (Hartung and Knapp, 2001a, b; IntHout et al., 2014). HCWs in low-risk units, HCWs who do not provide direct care (referred as no direct care henceforth) and men were used as the reference group in their respective OR analyses. Significance thresholds of *p* <0.05 was used in all analysis.

### Sensitivity analysis

Sensitivity analysis was performed by excluding each included study to identify any influential outliers that have significant influence on overall estimates and heterogeneity by assessing externally standardised residuals, difference in fits values (DFFITS), Cook’s distance, covariance ratios, DFBETAS, leave one out statistics, hat values and weights (Viechtbauer and Cheung, 2010). Prediction intervals (PI) were estimated when there were four or more studies to predict where 95% of future estimates would lie (Borenstein et al., 2017).

### Publication bias

Publication bias was assessed by visually inspecting funnel plots in conjunction with Egger test of asymmetry. A *p* -value <0.05 in Egger’s test indicated significant publication bias.

## Results

### COVID-19-Overall

In total there were 401 included studies from 392 unique datasets, with 18 longitudinal studies that reported on the prevalence rates across different timepoints. The included studies represented a total of 458 754 hospital HCWs across 58 countries (refer to S3A and S3B Appendix for study characteristics). Overall, five DSM-V mental health disorder were identified and reported by the included studies: depression, anxiety, PTSD, insomnia, and alcohol or substance use disorder (A/SUD). Two studies used diagnostic interviews, and the rest used validated self-report measurement tools and validated cut-offs that reflected a probable mental health disorder (refer to S3C Appendix for details on measurement tools used by studies and the corresponding cut-offs, sensitivity and specificity).

A summary of overall pooled estimates is displayed in Figure 2 (details in S4A to S8A Appendix). Overall meta-analytic pooling of individual estimates yielded an overall summary estimate of 28.5% for depression (95%CI: 26.3%-30.7%), 28.7% for anxiety (95%CI: 26.5%-31.0%), 25.5% for PTSD (95%CI: 22.5%-28.6%), 24.4% for insomnia (95%CI: 19.4%-29.9%) and 25.3 % for A/SUD (95%CI: 13.3%-39.6%). In all estimates heterogeneity was high (*I*^2^ = 99%-100%) and significant (*p* < 0.001), and prediction intervals were wide. Studies conducted in China accounted for the most weight in all the estimates (24.3% to 25.4%), followed by studies conducted in the United States (5.6% to 16.9%) (S4B to S8B Appendix).

**Figure 1.**
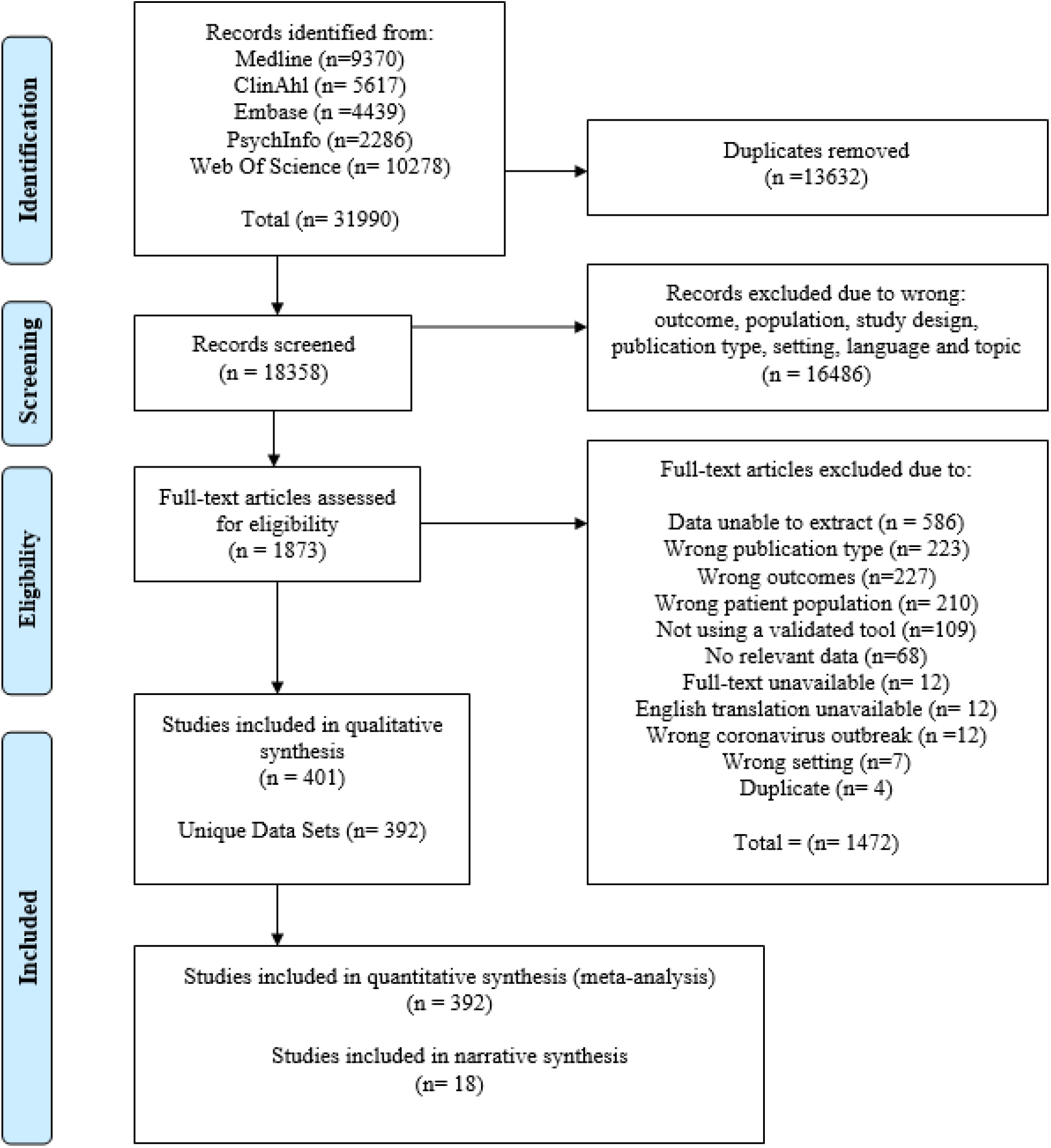
PRISMA flowchart.

**Figure 2.**
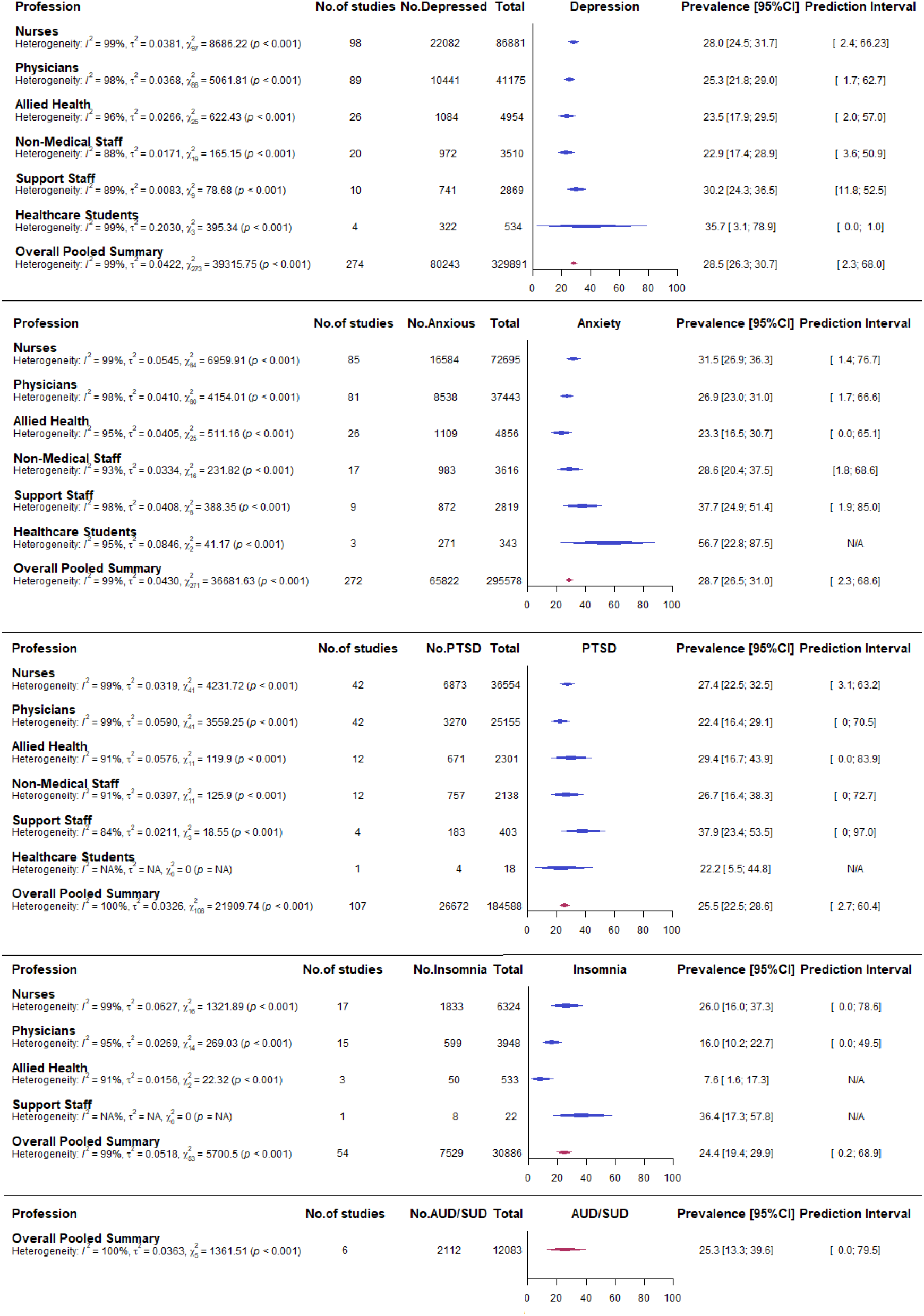
Forest plot of overall and stratified pooled prevalence estimates by individual professions.

### Subgroup analysis by study level characteristics

Subgroup analyses was conducted by measurement tools and WHO regions when there were more than four studies in each subgroup (Fig 3). Due to the lack of studies, subgroup analysis by measurement tools and region could not be conducted for A/SUD pooled rates. The test for subgroup differences showed statistically significant differences between estimates produced by the different measurement tools used for depression (*χ*^2^=56.7, df = 6, *p <* 0.001), anxiety (*χ*^2^=95.7, df = 5, *p <* 0.001), PTSD (*χ*^2^=21.7, df = 5, *p <* 0.001) and insomnia (*χ*^2^=47.9, df = 1, *p <* 0.001).

**Figure 3.**
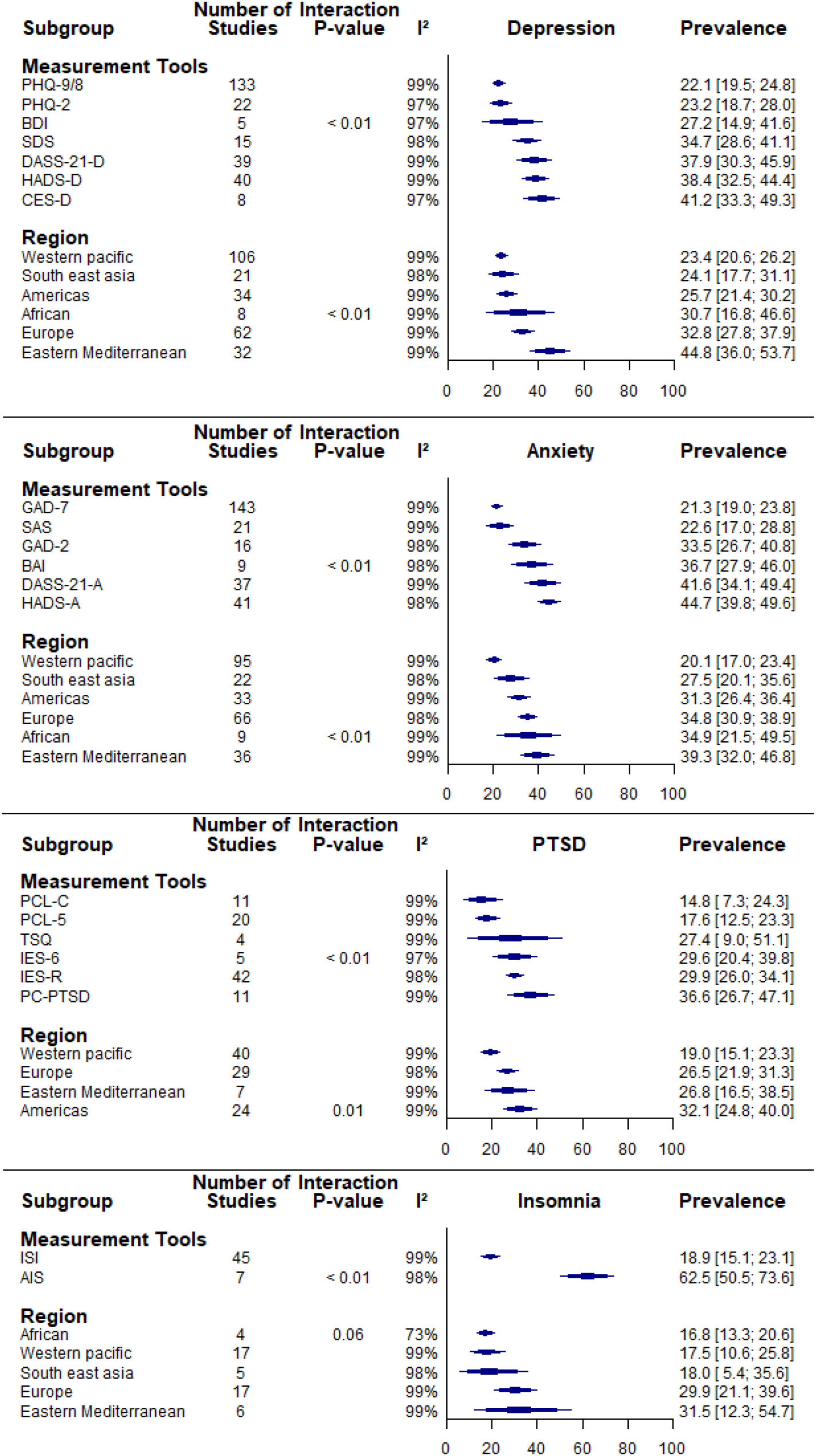
Forest plot of subgroup analysis by study level characteristics.

In terms of WHO regions, results showed statistically significant differences between the WHO regions’ overall pooled prevalence rates for depression (*χ*^2^=29.1, df = 5, *p <* 0.0001), anxiety (*χ*^2^=43.8, df = 5, *p <* 0.0001) and PTSD (*χ*^2^=11.3, df = 3, *p =* 0.01).

### Subgroup analysis by risk of bias

A summary of the overall risk of bias is displayed in Figure 4. There was low risk of bias in the sample frames used by 174 datasets (44%), sampling methods used by 140 datasets (36%), study’s sample size in 259 datasets (66%), study details reported in 342 datasets (87%), subgroup coverage in 14 datasets (4%), consistency in measurement across participants in 392 datasets (100%), statistical calculation of prevalence rates in 350 datasets (89%) and for response rate in 61 datasets (16%) (details in S3 Appendix). Overall, the studies show a mixed risk of bias across the results, with low risk of bias to accuracy and precision of the results but higher risk towards generalisability and representation of overall hospital HCW population. In the subgroup analysis, test of subgroup differences found no significant differences between estimates by JBI risk of bias criteria except for sample size, which significantly influenced depression (*χ*^2^=11.6, df = 1, *p <* 0.001), anxiety (*χ*^2^=14.5, df = 1, *p =* 0.0001), PTSD (*χ*^2^=7.41, df = 1, *p <* 0.01) and insomnia (*χ*^2^=5.1, df = 1, *p =* 0.02) rates (Fig5).

**Figure 4.**
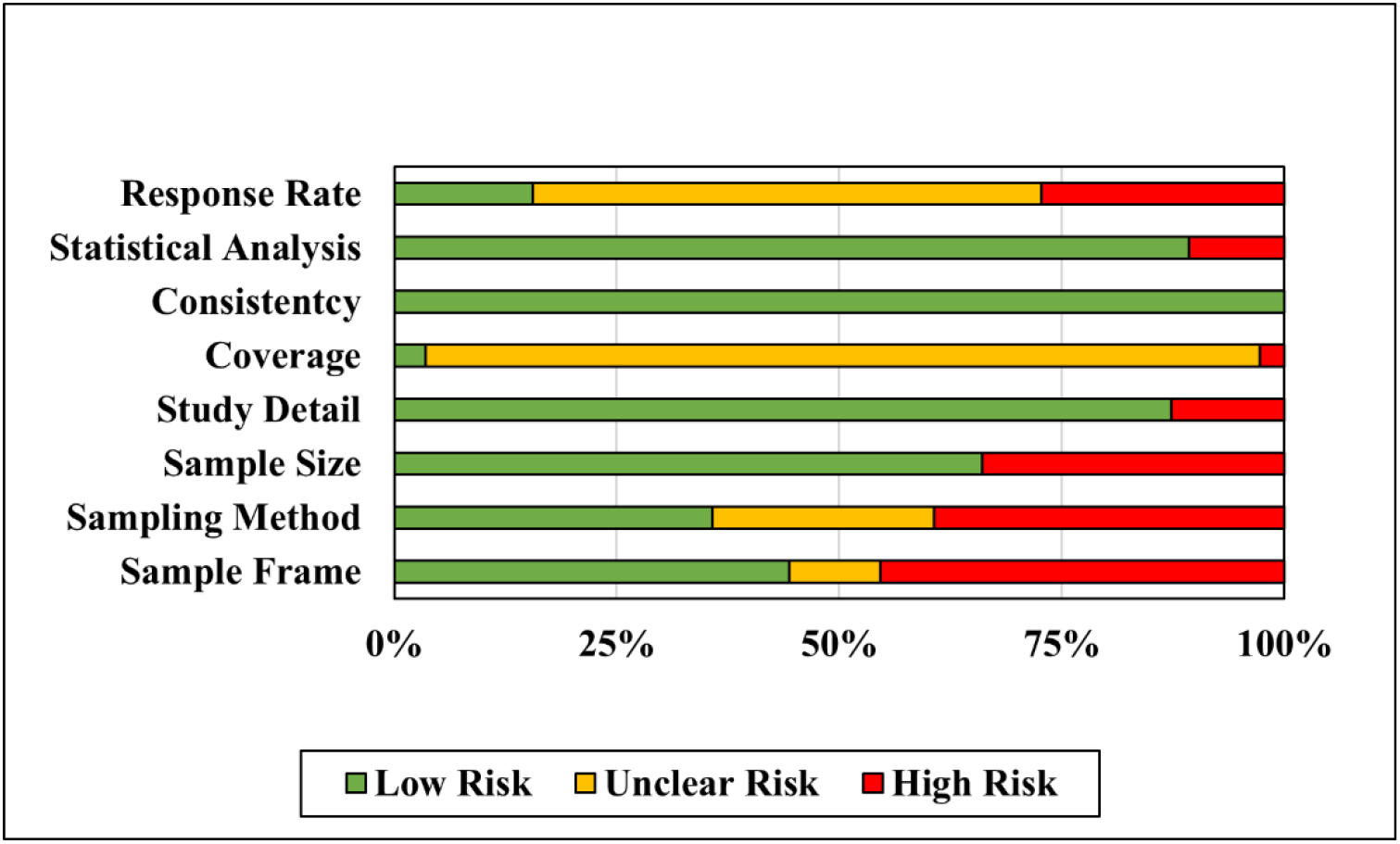
Risk of bias summary.

**Figure 5.**
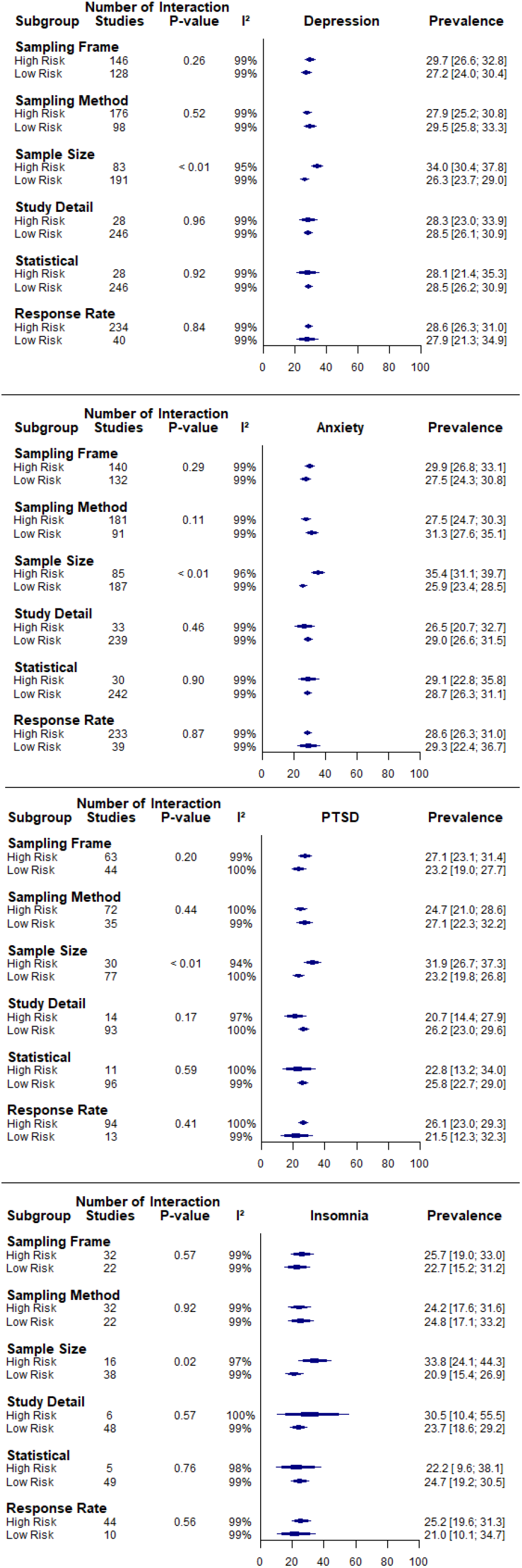
Forest plot of subgroup analysis by JBI risk of bias criteria.

### Prevalence estimates stratified by professional characteristics

A summary of pooled prevalence rates for all outcomes among individual professions are displayed in Figure 2.Results show that the pooled prevalence rates of depression was similar among nurses (28.0%, 95%CI: 24.5%-31.7%) and support staff (30.2%, 95%CI: 24.3%-36.5%), which was higher than physicians (25.3%, 95%CI: 21.8%-29.0%), allied health (23.5%, 95%CI: 17.9%-39.5%) and non-medical staff (22.9%, 95%CI: 17.4%-28.9%), but was lower than healthcare students (35.7%, 95%CI: 3.1%-78.9%) (S4C to S4H Appendix).

In terms of pooled anxiety rates, it was most prevalent among healthcare students (56.7%, 95%CI: 22.8%-87.5%), followed respectively by support staff (37.7%, 95%CI: 24.9%-51.4%), nurses (31.5%, 95%CI: 26.9%-36.3%), physicians (26.9%, 95%CI: 23.0%-31.0%), non-medical staff (28.6%, 95%CI: 20.4%-37.5%) and allied health (23.3%, 95%CI: 16.5%-30.7%) (S5C to S5H Appendix).

PTSD was most prevalent in support staff (37.9%, 95%CI: 23.4%-53.5%), followed by allied health (29.4%, 95%CI: 16.7%-43.9%), nurses (27.4%, 95%CI: 22.5%-32.5%), non-medical staff (26.7%, 95%CI: 16.4 %-38.3%), physicians (22.4%, 95%CI: 16.4%-29.1%) and healthcare students (22.2%, 95%CI: 5.5%-44.8%) (details in S6C to S6H Appendix).

Insomnia pooled prevalence rate was highest in support staff (36.4%, 95%CI: 17.3%-57.8%), followed by nurses (26.0%, 95%CI: 16.0%-37.3%), physicians (16.0%, 95%CI: 10.2%-22.7%) and lowest in allied health (7.6%, 95%CI: 1.6%-17.3%) (details in S7C to S7H Appendix).

No studies reported A/SUD rates by different professions.

Heterogeneity was high and significant for all estimates (*I*^2^= 84%-99%, *p <* 0.001). Prediction intervals were all wide and predicted wide ranges of pooled rates, indicating highly varied predicted rates.

### Odds ratio comparing prevalence rates of probable mental health disorders in Hospital HCWs working in high and low risk units

When comparing prevalence rates between hospital HCWs working in high risk and low risk department, high risk units had significantly higher pooled ORs for depression (OR=1.35,No. studies = 19, 95%CI:1.04-1.76, *t* = 2.39, *p* = 0.03), anxiety (OR=1.38, No. studies = 17, 95%CI:1.11-1.73, *t* = 3.12, *p* < 0.01), PTSD (OR= 1.59, No. studies = 12, 95%CI:1.26-2.02, *t*= 4.32, *p* < 0.01), and insomnia (OR= 1.33, No. studies =4, 95%CI=1.06-1.66, *t*= 3.99, *p* = 0.03) (Fig 6). There were no study reporting AUD rates by high or low risk units.

**Figure 6.**
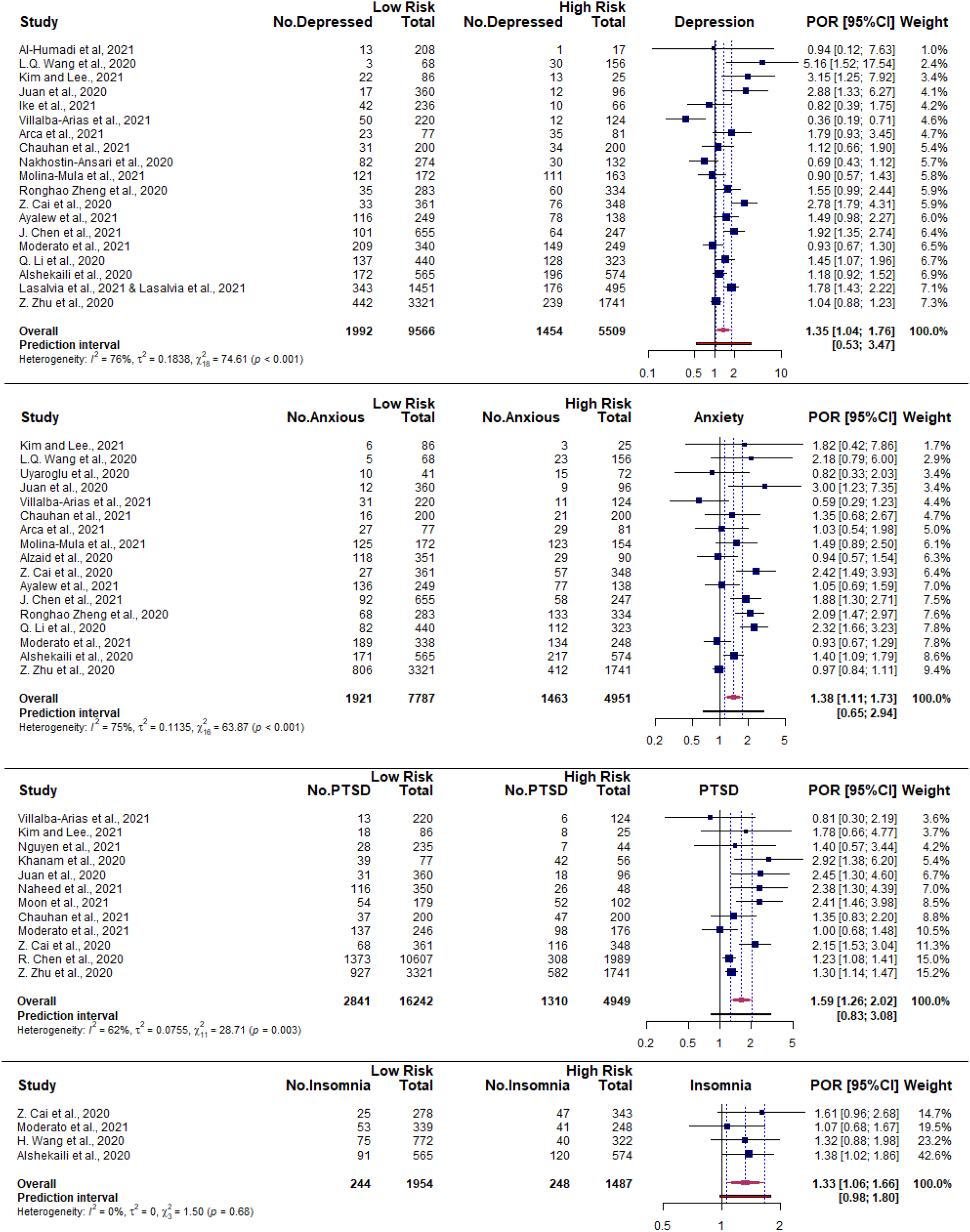
Forest plot of odds ratio comparing the prevalence of probable mental health disorders in Hospital HCWs working in high risk and low risk units.

Heterogeneity was high (*I*^2^ >75%) and significant (*p <*0.001) for depression and anxiety estimates comparing high and low risk units. Heterogeneity was moderate but significant for PTSD (*I*^2^= 62%, *p<* 0.01), whereas it was low and not significant for insomnia OR (*I*^2^= 0%, *p=*0.68). All prediction intervals predicted wide ranges of ORs and predicted non-significant effects in future estimates.

### Odds ratio comparing prevalence rates of probable mental health disorders in Hospital HCWs providing direct care to COVID-19 patients and those who do not

When comparing prevalence rates between Hospital HCWs who provide direct care and those who do not, those who provided direct care had significantly higher pooled ORs for depression (OR=1.48, No. studies = 36, 95%CI:1.24-1.77, *t* = 4.55, *p* < 0.0001), anxiety (OR=1.34, No. studies = 37, 95%CI:1.12-1.59, *t* = 3.38, *p* < 0.01), PTSD (OR=1.34, No. studies = 14, 95%CI:1.05-1.73, *t* = 2.56, *p* = 0.02) and insomnia (OR=2.17, No. studies = 10, 95%CI:1.74-2.71, *t* = 7.90, *p* = < 0.0001) (Figure 7). There was significant and high heterogeneity in OR estimates for depression and anxiety (*I*^2^ = 78%-79%, *p* <0.001). Heterogeneity was moderate but significant for PTSD (*I*^2^= 68%, *p <* 0.001), while insomnia pooled OR was not significantly heterogeneous and moderate (*p =* 0.12, *I*^2^= 37%). Prediction intervals for all OR estimates predicted non-significant effects, with the exception of insomnia, which had prediction intervals between 1.38 to 3.41

**Figure 7.**
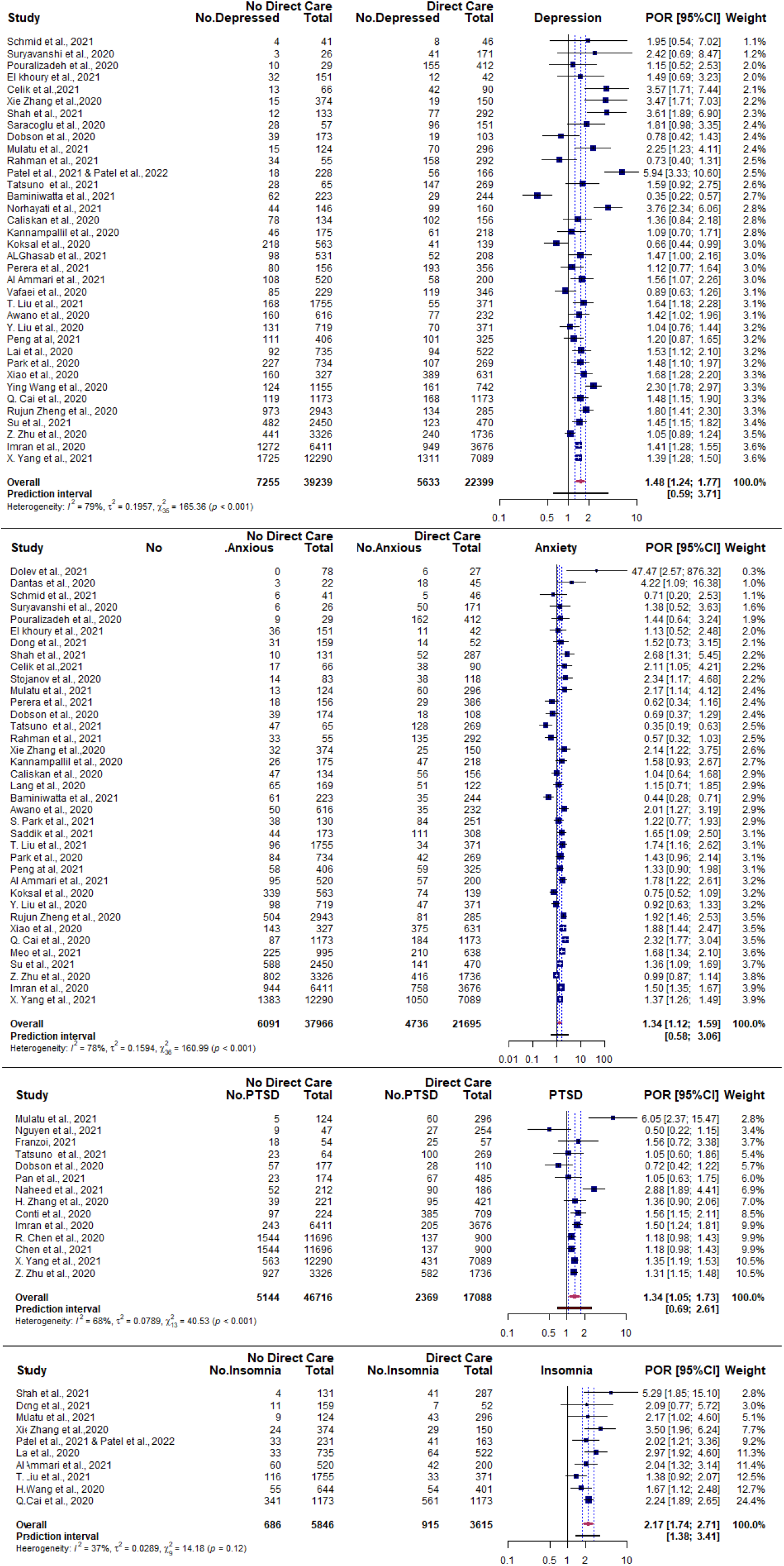
Forest plot of prevalence odds ratio comparing the prevalence of probable mental health disorders in Hospital HCWs providing direct care and no direct care.

### Odds ratio comparing prevalence rates of probable mental health disorders in men and women

When evaluating the OR for gender, pooled estimates show that women had significantly higher prevalence of depression (OR = 1.40, No. studies=79, 95%CI:1.26-1.56, *t* = 6.34, *p* < 0.0001), anxiety (OR = 1.50, No. studies=70, 95%CI:1.29-1.74, *t* = 5.47, *p* < 0.0001), PTSD (OR = 1.50, No. studies=40, 95%CI:1.30-1.74, *t* = 5.55, *p* < 0.0001), and insomnia (OR = 1.40, No. studies=15, 95%CI:1.23-1.60, *t* = 5.54, *p* < 0.0001).

As seen in Figure 8, there was significant heterogeneity and high (*p* < 0.01, *I*^2^> 77%) between studies in all estimates except insomnia OR, where it was low and not significant (*p =* 0.54, *I*^2^= 0%). Prediction intervals did not predict significantly higher rates of depression, anxiety and PTSD but predicted significantly higher rates of insomnia for women in future estimates.

**Figure 8.**
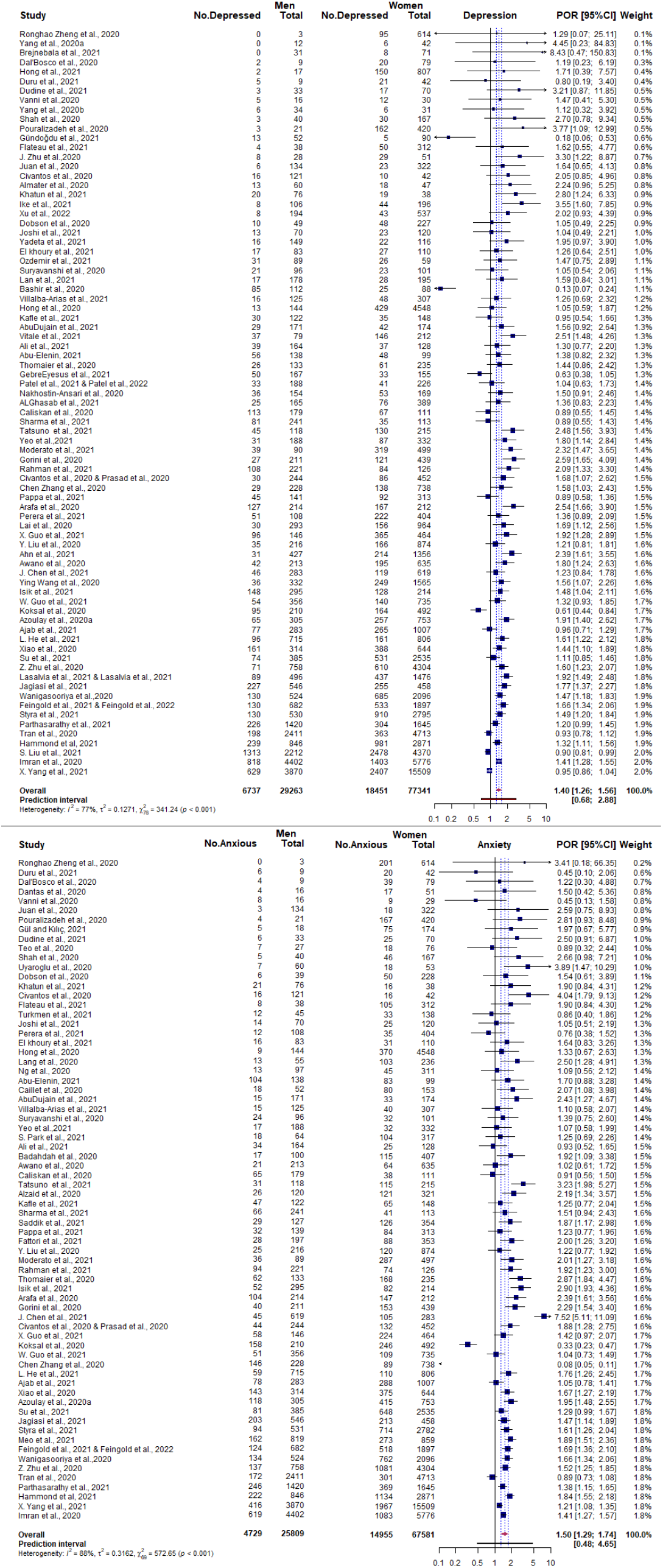
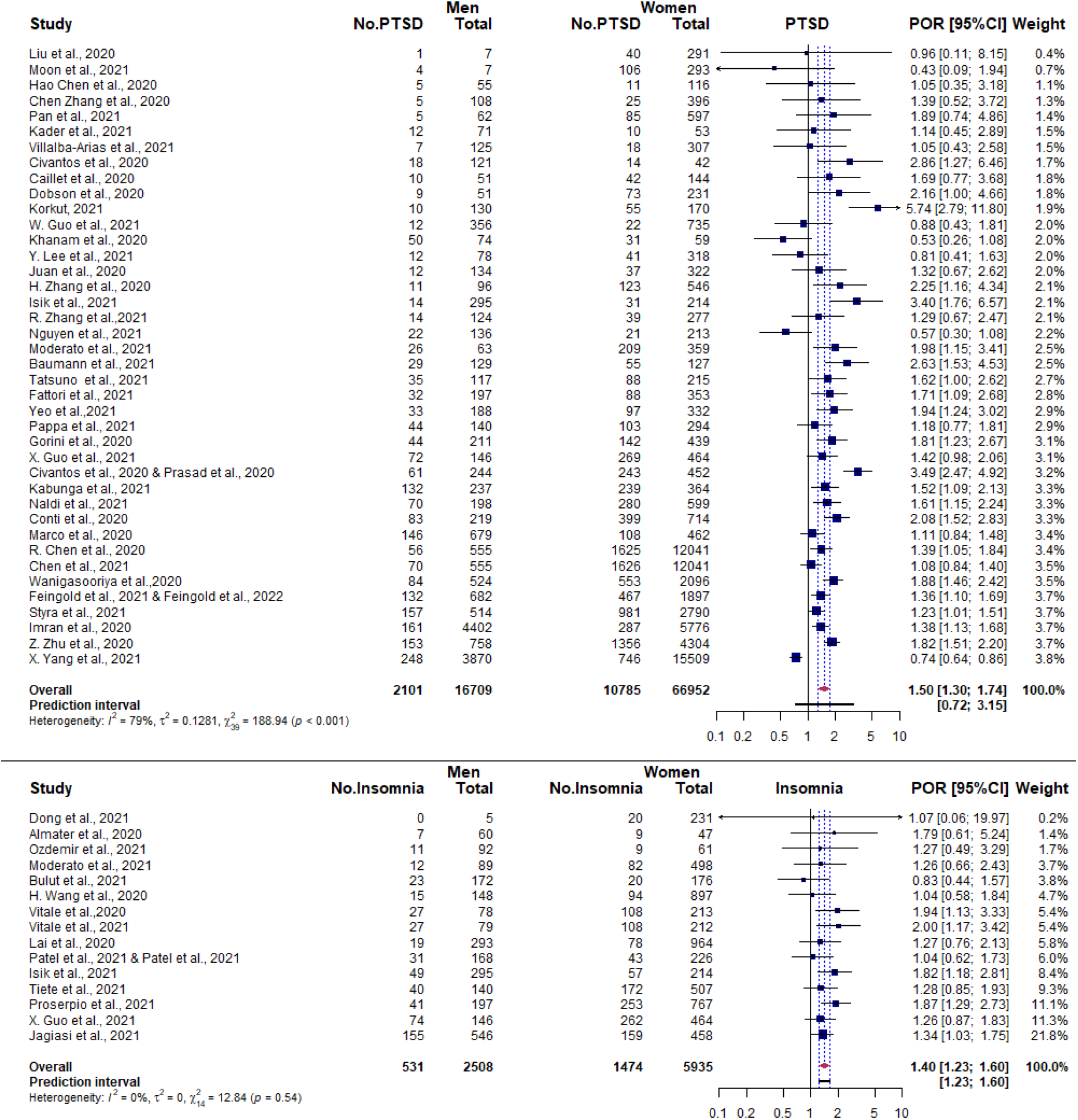
Forest plot of prevalence odds ratio comparing the prevalence of probable mental health disorders in females and males.

### Sensitivity analysis

In the overall pooled estimates, removal of influential outliers did not change any estimates by 4.7% and heterogeneity remained high (I^2 = 99%) and significant (p < 0.001).

In the stratified analysis for individual professions, with the exception of healthcare students’ pooled depression rates, removing outliers had no substantial or significant influence on pooled rates or their heterogeneity, with estimates differing by 3.1% at most and heterogeneity remaining high and significant (I^2= 85.0 %-99.2%, p < 0.001). With regards to healthcare students, removing the one outlier decreased pooled depression rates by 19.3%. The remaining three studies had a combined pooled rate of 16.4% (95%CI 6.6%-29.3%) but heterogeneity remained high and significant (*I*^2^ = 82.3%, *p <* 0.001), indicating that the high original pooled rates may be dependent on one study.

In the odds ratio analysis comparing prevalence rates between high risk and low risk units, one influential outlier was detected for insomnia OR but when removed did not change the significance of the effect, the overall effect size and heterogeneity (OR= 1.40, No. studies =3, 95%CI=1.14-1.72, t= 6.95, p = 0.02). No influential outliers were detected in depression, anxiety, and PTSD OR estimates.

No influential outliers were detected for any direct care OR estimates.

When influential outliers were removed in the OR estimates for gender, significance, effect size and prediction intervals were not affected substantially in depression (OR = 1.43, No. studies=78, 95%CI:1.31-1.56, t = 8.20, p < 0.0001), anxiety (OR = 1.53, No. studies=68, 95%CI:1.38-1.69, t = 8.24, p < 0.0001) and insomnia (OR = 1.42, No. studies=14, 95%CI:1.21-1.65, t = 4.82, p < 0.001). Heterogeneity remained significant but was reduced to a moderate level in depression (p < 0.0001, I^2= 73.5%) and anxiety estimates (p < 0.0001, I^2= 71.3%), which suggests that the high heterogeneity in the original estimate may be a result of extreme outliers. Heterogeneity in insomnia OR estimates remained low and non-significant (p = 0.47, I^2= 0%).

### Publication bias

There was significant publication bias detected in the overall prevalence estimates for depression (Egger’s test: *p* < 0.001), anxiety (Egger’s test: *p* < 0.001) and PTSD (Egger’s test: *p* < 0.001) (funnel plots displayed in S9 Appendix), with smaller studies producing higher prevalence rates. No publication bias was detected for insomnia (Egger’s test: *p* = 0.40) and AUD/SUD (Egger’s test: *p=* 0.12).

### Longitudinal COVID-19 studies

18 datasets conducted in 11 countries during COVID-19 reported the prevalence of probable mental health disorders across different timepoints (details in S10 Appendix) (Algattas et al., 2021; Baumann et al., 2021; Cai et al., 2020; Castioni et al., 2021; Chene et al., 2021; De Kock et al., 2022; Doulias et al., 2021; Gündoĝmuş et al., 2021; Katsuta et al., 2021; Lasalvia et al., 2021; Lasalvia et al., 2020; Li et al.; Magnavita et al., 2021a, b; Magnavita et al., 2020; Moore et al., 2021; Mosolova et al., 2021; Noaimi et al., 2021; Schmid et al., 2021; Shechter et al., 2021; Th’ng et al., 2021). Due to the variation in timepoints, differences in context, and the small number of studies, the temporal relationship between prevalence rates of mental health outcomes and the COVID-19 pandemic were narratively synthesised to avoid combining quantitative data inappropriately.

Two studies (Katsuta et al., 2021; Moore et al., 2021) compared pre-pandemic rates with COVID-19 rates in hospital HCWs. One study (Katsuta et al., 2021) (n-T1=2529, n-T2=2501) found no significant difference in the prevalence of depression pre-pandemic and during the COVID-19 pandemic among hospital HCWs. Similar results were found for the prevalence of anxiety in Moore et al. (2021) study among medical students (n-T1=3775, n-T2=836), which found no significant difference in both timepoints.

Eight datasets from 11 studies (Baumann et al., 2021; De Kock et al., 2022; Lasalvia et al., 2021; Lasalvia et al., 2020; Magnavita et al., 2021a, b; Magnavita et al., 2020; Noaimi et al., 2021; Schmid et al., 2021; Shechter et al., 2021; Th’ng et al., 2021), investigated the progression of mental health outcomes across different timepoints as part of follow up assessments. One study (Shechter et al., 2021) followed participants (T1 to T6-n= 230) over six timepoints over 10 weeks. They found that the high prevalence of PTSD found during baseline decreased by 34.9% over the first five timepoints but did not decrease further in the last timepoint. In another study (Baumann et al., 2021) with ED physicians (n-T1=262, n-T2=252), PTSD also decreased after two months but at a lower rate of 7%. With regards to depression, the prevalence of depression significantly increased by 11.4% over 8 months in one dataset (Magnavita et al., 2021a, b; Magnavita et al., 2020) (n-T1=180, n-T2=152, n-T3=120), and 14% over 12 months in another (Lasalvia et al., 2021; Lasalvia et al., 2020) (n-T1=2195, n-T2=917). In four other datasets (De Kock et al., 2022; Noaimi et al., 2021; Schmid et al., 2021; Th’ng et al., 2021), with an average follow up duration of 8.5 months (2-13 months), the prevalence of depression did not significantly change from baseline and remained high. In terms of the prevalence of anxiety, five datasets (De Kock et al., 2022; Magnavita et al., 2021a, b; Magnavita et al., 2020; Noaimi et al., 2021; Schmid et al., 2021; Th’ng et al., 2021) with an average follow up duration of 8 months (2-13 months), showed that it remained similar to baseline rates. Only one dataset (Magnavita et al., 2021a, b; Magnavita et al., 2020) reported on the prevalence of insomnia (n-T1=180, n-T2=152, n-T3=120), which showed significantly lower prevalence of insomnia after eight months.

Two studies reported prevalence rates of probable mental health disorders before and one month after COVID-19 outbreaks in their local area (Cai et al., 2020; Doulias et al., 2021). In Cai et al. (2020) study, they investigated two separate samples of nurses (n-T1=709, n-T2=621) recruited from the same sample frame during the two different timepoints. Their results shows that the prevalence of depression, anxiety, PTSD had a significant decrease after the outbreaks, but insomnia remained similar to baseline rates. Doulias et al. (2021) had similar results in their repeated measures study (n-T1:93, n-T2:103) with regards to the prevalence of depression and anxiety, showing a large decrease in prevalence rates of both outcomes.

One study (Li et al.) looked at hospital nurses’ (T1 & T2-n=356) levels of PTSD symptoms assessed before and after they worked in the COVID-19 units. Results showed that there was a significant and large increase in the prevalence of clinically significant PTSD symptoms after they have worked in COVID-19 units.

One study (Algattas et al., 2021) with a small sample of neuropsychology residents (n=17) found that the prevalence of depression during a COVID-19 outbreak was significantly lower than rates found before the outbreak.

Two repeated measures study (Gündoĝmuş et al., 2021; Mosolova et al., 2021) compared prevalence rates during the peaks of the first and second COVID-19 outbreak in their local area. One study (Gündoĝmuş et al., 2021) (n-T1:1051, n-T2:1409) reported that depression and anxiety prevalence rates among hospital HCWs was significantly higher during the second wave of COVID-19 outbreak compared to the first wave. This was consistent with Mosolova et al. (2021) study (n-T1:1090, n-T2:1105), which showed a significant increase in the prevalence of anxiety in the second wave of COVID-19.

Two studies (Castioni et al., 2021; Chene et al., 2021) compared prevalence rates before and after lockdown measures were implemented in their local area. Chene et al. (2021) results (n-T1:1565, n-T2:1109) showed that after lockdown, no significant changes were observed in the prevalence of depression and anxiety. Consistent with this, Castioni et al. (2021)’s result (n=272) showed that the prevalence of anxiety remained the same but the prevalence of depression significantly increased after lockdown.

## Discussion

To estimate the impact of the COVID-19 pandemic on hospital HCWs mental health, this study systematically reviewed the literature and conducted a meta-analysis of studies reporting on probable mental health disorders in hospital HCWs. The results from this study’s meta-analysis of 392 datasets during the COVID-19 pandemic suggest that a substantial proportion of hospital HCWs working during COVID-19 experienced clinically significant symptoms of depression (28.5%), anxiety (28.7%), PTSD (25.5%), insomnia (24.4%), and AUD (25.3%). With regards to published findings, preliminary estimates from early reviews on hospital HCWs have widely varied (Li et al., 2021a; Raoofi et al., 2021; Salari et al., 2020a; Salari et al., 2020b), likely due to the differences in the number of studies, sample size and methodology. Nonetheless, overall findings from this study echo those from these previous reviews, that the prevalence of mental health issues in hospital HCWs is high and concerning. In addition, findings from this study have provided an updated and more precise estimate of the overall estimated prevalence of mental health issues being experienced by hospital HCWs during COVID-19, which has remained high even with newer studies, additional countries, a substantially larger sample size and the addition of clinically significant thresholds. Given these findings and the ongoing nature of the COVID-19 pandemic, it is evident that support is imminently needed to prevent, reduce, and treat these clinically significant symptoms of mental health disorders found in hospital HCWs working during COVID-19.

When compared to the most recent estimates for the general public (GP) during COVID-19, our findings indicate that hospital HCWs overall had a higher prevalence of depression (HCWS: 28.5.0% vs GP: 22.6%) and anxiety (HCWS: 28.7% vs GP: 22.4%) (Phiri et al., 2021). This is not surprising given the increases in workloads, infection risks, uncertainty, and feelings of threat during the pandemic (Ding et al., 2021; Joo and Liu, 2021; Liu et al., 2020), which may have increased the likelihood of developing depression and anxiety in hospital HCWs during this time. The same, however, could not be said about the prevalence rates of PTSD (HCWs: 25.5% vs GP: 23.2%) and insomnia (HCWS: 24.4% vs GP: 22.9) in hospital HCWs, which have been comparable to the general public (Phiri et al., 2021). Given these findings it cannot be disregarded that the prevalence of mental health issues in hospital HCWs, such as PTSD and insomnia, may be explained partly, if not fully, by distressing experiences in the community that have been associated with mental health issues during the COVID-19 pandemic, such as the risk of infections in the community, lockdowns, quarantines, impacts on families and social distancing (Brooks et al., 2020; Sheen et al., 2022; Yuan et al., 2021a).Thus, when developing mental health support strategies for hospital HCWs it is important to consider the impact of a pandemic across the different domains of their lives as well as the direct impacts of their healthcare roles.

When interpreting findings, it is also important to recognise there was high heterogeneity in the overall estimates, which is common to meta-analyses of prevalence and consistent with previous reviews (Li et al., 2021a; Raoofi et al., 2021; Salari et al., 2020a; Salari et al., 2020b). To further investigate the extent of this heterogeneity, prediction intervals were estimated, providing insight into the distribution and predicted range of future estimates (Barker et al., 2021; Borenstein et al., 2017). Our analysis identified wide ranges of predicted prevalence rates that indicate, while on average there may be a high prevalence of probable mental health disorders in hospital HCWs during the COVID-19 pandemic, it is likely that prevalence rates vary substantially between different settings, groups, or study design. In our findings, sample size was found to significantly affect all effect sizes, with lower risk studies and smaller studies showing a lower average in prevalence rates.

Prevalence estimates were also found to significantly differ between different WHO regions and measurement tools, which is likely explained by the considerable differences in optimal specificity and sensitivity in the measurement tools used (S2 Appendix) as well as national differences in COVID-19’s spread and responses. As such, it is important to consider these factors when interpreting findings. However, while these factors may impact findings, there is low certainty they had a large and significant impact as there were still large amounts of unexplained heterogeneity. Another important factor to consider when interpreting findings is clinical heterogeneity, (e.g., participant characteristics) which our findings indicate could also explain differences in outcomes given consistent findings in specific estimates.

When analysed by healthcare professions, findings demonstrate that clinical symptoms of depression, anxiety, PTSD, and insomnia were highly prevalent across all studied healthcare professions. There was, however, variations in prevalence rates of different mental health outcomes among different professions, suggesting that different professions may be susceptible to different mental health impacts. In terms of depression only physicians, nurses, support staff and healthcare students had higher rates than estimates for the general public (Phiri et al., 2021). With regards to physicians, hospital nurses and healthcare students, pre-pandemic studies largely supports that mental health issues have been evident in this cohort since pre-pandemic (Chen and Meier, 2021; Dyrbye et al., 2006; Gheshlagh et al., 2017; Rotenstein et al., 2016; Tung et al., 2018; Xie et al., 2020), due to their high workload, work-related stress and high levels of burnout associated with their work (Almutairi et al., 2022; Chen and Meier, 2021; Koutsimani et al., 2019). This makes it unclear to what extent pandemic factors alone have affected their rates of depression symptoms while working in hospitals during this time. Nevertheless, it is likely that pandemic factors exacerbated their existing vulnerabilities, placing them at a higher risk of developing depression symptoms during the COVID-19 pandemic. In contrast, support staff and other healthcare professionals have largely been left out of pre-pandemic research. This is concerning given that translating evidence from other professions may not be effective due to the differences in work related stress, roles, and tasks. Based on these findings it is recommended that further research examine profession specific factors that may be influencing the prevalence of depression symptoms in hospital HCWs to support further development of targeted mental health support strategies within hospital systems.

With regards to findings on anxiety, there is little pre-pandemic research indicating this was a pre-existing issue for hospital HCWs overall, suggesting that the high rates of anxiety found in this study may be explained by pandemic factors. This is affirmed by the literature showing that sources of anxiety in hospital HCWs during coronavirus outbreaks, including COVID-19, largely revolves around nosocomial infection risks, such as fear of being infected (De Brier et al., 2020; Serrano-Ripoll et al., 2020), fear of infecting others (i.e., family and community members) (Barello et al., 2020; Kisely et al., 2020), and PPE adequacy (Kisely et al., 2020; Troglio Da Silva and Neto, 2021). The trends in our findings also support this pattern of anxiety, showing the highest rates of anxiety in nurses, physicians, support staff and healthcare students, all of whom have patient facing intensive roles and thus more risks of infection. In contrast, allied health staff who have less patient contact, such as pharmacists and lab technicians, were found to have the lowest rates of anxiety, comparable to the general public (Phiri et al., 2021). Of note, non-medical staff had higher rates of anxiety than the general public even though they were likely to have little patient contact. This could be a reflection of their lack of specialised training and experience, which have been suggested to increase worries of infections and mental health issues during coronavirus outbreak (De Brier et al., 2020; Kisely et al., 2020). This re-emphasises the need for healthcare organisations to incorporate strategies that help all hospital HCWs feel safe during this time. Though, it is important to note that concerns around high infection risks among HCWs were raised in the early stages of COVID-19 (Erdem and Lucey, 2021) and infection risks may have reduced with improvements in the healthcare system over the course of the COVID-19 pandemic. Early evidence from the United States have demonstrated that HCWs’ risk of infection in the workplace have reduced as hospitals improve their infection control procedures, PPE use and vaccination rates (Braun et al., 2021; Damluji et al., 2021; Dunbar et al., 2021; Jacob et al., 2021; Moghadas et al., 2021). While new variants of COVID-19 may create a resurgence of infections again, it is not certain whether these high anxiety rates persist, and if they do, whether nosocomial infection risks still play a significant role. Nevertheless, given that anxiety symptoms can be a precursor to depression (Batterham et al., 2013), support strategies will still benefit from focusing on reducing anxiety symptoms in addition to depression symptoms to reduce the overall high rates of mental health issues in hospital HCWs during the COVID-19 pandemic and future infectious disease outbreaks.

With PTSD findings, interpretation needs to consider that findings may not be reflective of post-traumatic stress reactions but rather hospital HCWs’ ongoing and acute experiences with traumatic stress as assessment of symptoms largely occurred during outbreaks, suggesting that delayed onset of PTSD symptoms may yet occur. In the findings, while overall rates of PTSD in hospital HCWs were similar to those in the general public, nurses, allied health, non-medical staff and support staff were more likely to experience PTSD symptoms than the general public (Phiri et al., 2021). Again, infection risks may be playing an important role here, however, indirectly. Due to increased infection risks, rates of quarantine have been significantly higher in hospitals HCWs compared to the general public (Kumar et al., 2020) and have been found to be associated with PTSD symptoms (Brooks et al., 2020; Brophy et al., 2020; Carmassi et al., 2020). While an effective measure to reduce nosocomial outbreaks of COVID-19 in hospitals (Grzelakowska and Kryś, 2021; Huang et al., 2021), studies have demonstrated that quarantine procedures can have severe psychological impact due to feelings of threat, restrain and social isolation (Basso et al., 2021; Brooks et al., 2020; Gómez-Durán et al., 2020), which were likely intensified as hospital HCWs face repeated quarantine and its’ consequences due to their work in hospitals. Another contributing factor could be the social stigma and social rejection (Ding et al., 2021; Gómez-Durán et al., 2020; Schubert et al., 2021; Yuan et al., 2021b) faced by these professional groups due to their work, which have been found to increase experiences of PTSD symptoms (Yuan et al., 2021a). Nurses, allied health, non-medical staff and support staff may have been exposed more frequently to these negative social experiences given their increased contact with patients and the public outside of clinical activities.

In addition to professional differences, it is also important to consider the impacts of occupational factors on hospital HCWs’ mental health, such as working in high-risk units and providing direct care to infected patients. Our findings suggest that both these factors have led to significantly higher odds of developing depression, anxiety, PTSD and insomnia symptoms compared to their counterparts, with highly consistent evidence for insomnia and moderately consistent for PTSD. Overall, these findings re-enforce that proximity to risk is likely to be a determinant of distress during traumatic events (May and Wisco, 2016) and highlights the need for strategies and interventions to support these at-risk groups during the COVID-19 pandemic and in future infectious disease outbreaks. However, while these findings affirm those from previous coronavirus outbreaks (De Brier et al., 2020; Kisely et al., 2020; Preti et al., 2020; Serrano-Ripoll et al., 2020), it has contradicted recent results from a living meta-analysis (Bell and Wade, 2020) that have shown exposure to COVID-19 patients do not significantly increase risk of PTSD. This could likely be a result of conflating hospital HCWs with general HCWs and severe symptoms with mild in their meta-analysis (Bell and Wade, 2020) and thus increasing the rates of PTSD symptoms in their non-exposed group, leading to a non-significant effect. This suggests that the effect found in our meta-analysis may only be reflective of those working in hospitals and PTSD symptoms that are at a clinically significant threshold. Nevertheless, our overall findings still echoes Bell and Wade’s (Bell and Wade, 2020) recommendations that trauma focused support may not be a pertinent focus in pandemic responses in the overarching healthcare system. When we look at the overall impact on PTSD symptoms in hospital HCWs, there is some basis to their recommendations as overall rates are comparable to the general public, which may mean that reducing traumatic stress during COVID-19 would be more effective on a national level and will likely have a significant cascading effect in reducing traumatic stress in hospital HCWs overall. Within the healthcare system our findings suggest that trauma focused support, such as psychological first aid (Ruzek et al., 2007), may instead be more appropriate for specific groups at-risk due to their profession or proximity to infection risks, as opposed to a macro level response from healthcare organisations.

Attention should also be given to hospital HCWs who are women. Our findings clearly indicate they have been impacted more than men across multiple outcomes, with consistent evidence in regard to insomnia and with the exception of extreme cases, depression, and anxiety. However, while this supports evidence from previous coronavirus outbreaks (Carmassi et al., 2020; Kisely et al., 2020; Serrano-Ripoll et al., 2020), and the COVID-19 pandemic (Batra et al., 2020; Zhao et al., 2021), it is again unclear to what extent working in hospitals and pandemic factors are impacting their mental health. For instance, women make up a large proportion of the healthcare workforce, such as nurses (World Health Organisation, 2020) who have been implicated to have higher rates of mental health issues in this study’s findings and may explain women’s higher likelihood of experiencing mental health issues. In addition, the mental health of women has also been found to be impacted more in the general public during pandemics (Xiong et al., 2020) and out of pandemics (Wang et al., 2016). Nonetheless, while it is not certain that the hospital system or pandemic factors are the main causal factor, pandemic factors likely played a significant role in exacerbating existing mental health vulnerabilities among hospital HCWs who are women. It is also important to note that findings have rejected that biological vulnerabilities, such as hormones, explain the gender differences in mental health outcomes (Wang et al., 2016), which underscores the importance of examining and addressing social factors that are influencing disparities in mental health outcomes between men and women working in hospitals.

Turning to longitudinal trends, findings were weak due to paucity of research, but they suggested that probable mental health disorders were most prevalent in hospital HCWs leading into and during an outbreak but decreased after outbreaks. Similar findings have been identified amongst the general public during COVID-19 (Bourmistrova et al., 2022; Robinson et al., 2022), showing rates of mental health issues decreasing as the COVID-19 pandemic progressed, implying that acute mental health responses are likely to be more severe than long-term mental health responses. Though, despite the decrease, prevalence rates remained at concerning rates and persisted among hospital HCWs over time. Previous findings suggest that this particularly occurs for those working in high-risk settings that may be at risk of long-term mental health issues after infectious disease outbreaks (Chau et al., 2021). Given these findings, mental health strategies should consider strategies to mitigate any long-term impacts. Longitudinal findings from a small number of studies, also supports that pre-pandemic mental health issues were likely present and translated into higher mental health risks during the COVID-19 pandemic. This highlights the importance of long-term strategies to address systemic issues that leave hospital HCWs vulnerable to mental health issues during traumatic events such as infectious disease outbreaks. Given these findings and the low certainty in findings due to a paucity in longitudinal research, follow-up research is recommended, specifically on those who are at higher risk, such as those providing direct care to COVID-19 patients and HCWs working high risk departments.

## Limitations

While this review has attempted to strengthen confidence when interpreting findings using conservative methodology, it is not without its limitations. First, the findings are only as robust as the studies included, which have mostly used self-report measures, reducing the certainty that estimates truly reflect mental health diagnoses. However, we have minimised uncertainty with this by only including studies that use measurement tools and cut offs that have been validated against diagnostic interviews. While not reflective of actual mental health diagnoses, our findings still show for the first time the proportion of hospital HCWs experiencing mental health issues at a clinical level that have a high probability to lead to a mental health disorder, if not present already, and requires immediate clinical assessment (Bressler et al., 2018; Kroenke et al., 2007; Nieuwenhuijsen et al., 2003).

It also cannot be rejected that studies were missed in the selection process. Due to the lack of multilingual reviewers in this study, non-English studies were not included. In addition, not all studies were screened twice, however, evidence suggests that partial dual screening can still be an acceptable procedure (Taylor-Phillips et al., 2017; Waffenschmidt et al., 2019). Despite these limitations, our review with 401 studies is the most comprehensive to date on mental health outcomes of hospital HCWs during COVID-19. As such, there is very low risk of missing studies significantly impacting results.

## Conclusion

In sum, findings are overall still emerging and there is growing certainty in the evidence to suggest that there are specific groups of hospital HCWs at higher risk of mental health issues. Thus, it is certain that further research is needed to inform the development of mental health support strategies that are targeted. This is especially true for those outside of the nursing and physician professions, such as allied health, non-medical and support staff in hospital settings, where there is a lack of evidence to effectively rely on. At this time, findings also highlight that those working in direct proximity of COVID-19 patients, working in high-risk units and women hospital HCWs are at significant risk of mental health issues and should be a priority in staff support within hospitals. More importantly, with re-occurring outbreaks from new COVID-19 variants, mental health issues in hospital HCWs may continue to persist and should not be neglected. As such, we echo the many pervious calls for further research, development of effective interventions and initiatives to support hospital HCWs as they continue to support their communities while facing the many challenges of the COVID-19 pandemic.

## Data Availability

All data produced in the present work are contained in the manuscript

## Contribution

Formal analysis, data curation, investigation, writing – Original Draft Preparation: Brian Lee En Chyi

Methodology: Brian Lee En Chyi, Mathew Ling, Leanne Boyd, Jade Sheen

Conceptualisation, supervision, writing – review & editing: Mathew Ling, Leanne Boyd, Craig Olsson, Jade Sheen

All authors have contributed and approved the final manuscript

## Acknowledgements

The authors would like to acknowledge and thank Dawson Peter Grace, Mikhayl von Riebon, Gayatri Kumar, Caitlyn Ann Herrick, Hiba Sidiqui, Anuradhi Jayasinghe and Britney Dellios for their assistance with screening, data extraction and data checking.

## Supporting information

**S1 Appendix. Prisma checklist**

**S2 Appendix. Methods supplement**

**S3A Appendix. Summary table of study characteristics**

**S3B Appendix. Included studies references**

**S3C Appendix. Measurement tools and cut off scores**

**S4 Appendix. Depression forest plots**

**S5 Appendix. Anxiety forest plots S6 Appendix. PTSD forest plots**

**S7 Appendix. Insomnia forest plots S8 Appendix. AUD forest plot**

**S9 Appendix. Funnel plots**

**S10 Appendix. COVID-19 longitudinal studies summary table**

