## Supplements for "The prevalence of probable mental health disorders among hospital healthcare workers during COVID-19: A systematic review and meta-analysis": S2 Appendix.Methods.docx

| **S2A Appendix.** **Sample search strategy in Embase** | |
| --- | --- |
| Key Concept | Similar search terms and truncations |
| Healthcare worker  AND | 'healthcare worker*':ti, ab OR 'healthcare personnel*':ti, ab OR 'healthcare professional*':ti, ab OR 'medical professional*':ti, ab OR 'medical worker*':ti, ab OR 'medical staff*':ti, ab OR 'medical personnel*':ti, ab OR 'hospital personnel*':ti, ab OR 'hospital staff*':ti, ab OR 'hospital worker*':ti, ab OR 'hospital employee*':ti, ab OR 'medical employee*':ti, ab OR 'healthcare employee*':ti, ab OR 'health care worker*':ti, ab OR 'health care personnel*':ti, ab OR 'health care employee*':ti, ab OR 'health care staff*':ti, ab OR 'healthcare staff*':ti, ab OR 'allied health staff*':ti, ab OR 'allied health professional*':ti, ab OR 'allied health personnel*':ti, ab OR 'allied health worker*':ti, ab OR 'doctor*':ti, ab OR 'nurse*':ti, ab OR 'physician*':ti, ab OR 'clinician*':ti, ab OR 'clinical assistant*':ti, ab OR 'ward clerk*':ti, ab OR 'hospital volunteer*':ti, ab OR 'hospital administrator*':ti, ab OR 'medical practitioner*':ti, ab OR 'healthcare practitioner*':ti, ab OR 'health care practitioner*':ti, ab OR 'allied health practitioner*':ti, ab OR 'health worker*':ti, ab OR 'health professional*':ti, ab OR 'health practitioner*':ti, ab OR 'health employee*':ti, ab OR 'health staff*':ti, ab OR 'emergency department staff*':ti, ab OR 'icu staff*':ti, ab OR 'intensive care unit staff*':ti, ab OR 'infectious disease unit*':ti, ab OR 'infectious disease ward*':ti, ab OR 'health care personnel'/exp OR 'health care personnel' |
| Mental health  AND | 'mental health':ti, ab OR 'wellbeing':ti, ab OR 'emotional':ti, ab OR 'psycholog*':ti, ab OR 'psychoso*':ti, ab OR 'depression*':ti, ab OR 'depressive*':ti, ab OR 'anxi*':ti, ab OR 'psychological distress*':ti, ab OR 'post-traumatic stress*':ti, ab OR 'insomnia*':ti, ab OR 'burnout*':ti, ab OR 'burn-out*':ti, ab OR 'stress*':ti, ab OR 'stress'/exp OR 'stress' OR 'depression'/exp OR 'depression' OR 'anxiety'/exp OR 'anxiety' OR 'posttraumatic stress disorder'/exp OR 'posttraumatic stress disorder' OR 'anxiety disorder'/exp OR 'anxiety disorder' OR 'professional burnout'/exp OR 'professional burnout' OR 'insomnia'/exp OR 'insomnia' OR 'mental health'/exp OR 'psychological adjustment'/de OR 'wellbeing'/de OR 'emotional stress'/de |
| Coronavirus | '2019-ncov':ti, ab OR 'coronavirus':ti, ab OR 'COVID19':ti, ab OR 'COVID':ti, ab OR 'COVID-19':ti, ab OR 'ncov':ti, ab OR 'severe acute respiratory syndrome*':ti, ab OR 'sars-cov-2':ti, ab OR 'sars':ti, ab OR 'middle east respiratory syndrome*':ti, ab OR 'mers':ti, ab OR 'sars-related coronavirus'/exp OR 'sars-related coronavirus' OR 'middle east respiratory syndrome coronavirus'/exp OR 'middle east respiratory syndrome coronavirus' OR 'middle east respiratory syndrome'/exp OR 'middle east respiratory syndrome' OR 'severe acute respiratory syndrome'/exp OR 'severe acute respiratory syndrome' OR 'coronavirus disease 2019'/exp OR 'coronavirus disease 2019' |

| **S2B Appendix.** **Risk of Bias Assessment For Prevalence Studies** | |  |
| --- | --- | --- |
| JBI Assessment criteria | Criteria for low risk of bias | Criteria for high risk of bias |
| Was the sample frame appropriate to address the target population? | Recruited participants from multicentre hospitals | Recruited participants from social media or a single centre only |
| Were study participants sampled in an appropriate way? | Used a probabilistic sampling method | Used a convenience sampling method |
| Was the sample size adequate? | Attained required sample $\mathrm{size}^{a}$ | Did not meet the required sample size |
| Were the study subjects and the setting described in detail? | Described in participant characteristics (i.e., age and gender characteristics, hospital setting, recruitment dates and country of setting) | Had missing information on participant characteristics (i.e., age and gender characteristics, hospital setting, recruitment dates and country of setting) |
| Were valid methods used for the identification of the condition? | Used a validated measurement tool and method or optimal cut offs recommended by the literature | Used a measurement tool that has not been validated or used a cut off or method that has not been validated to identify individuals with a mental health disorder |
| Was the condition measured in a standard, reliable way for all participants? | Used the same measurement tool and method for all participants | Inconsistent methods or measurement tools used to identify participants with a mental health disorder |
| Was there appropriate statistical analysis? | Reported the numerator and denominator used to calculate prevalence rates | Reported prevalence rates without raw values used to estimate prevalence rates |
| Was the response rate adequate, and if not, was the low response rate managed appropriately? | Response rate was 80% or above | Response rate was below 80% |
| Was the data analysis conducted with sufficient coverage of the identified sample? | All subgroups had comparable response rates | Had an under or overrepresented subgroup |
