## Supplements for "The prevalence of probable mental health disorders among hospital healthcare workers during COVID-19: A systematic review and meta-analysis": S3B Appendix.References.docx

Abdellah, M.M., Khalil, M.F., Alhafiz, A., 2021. Prevalence of Poor Sleep Quality Among Physicians During the COVID-19 Pandemic. Cureus 13, e12948.

Abdoli, N., Farnia, V., Jahangiri, S., Radmehr, F., Alikhani, M., Abdoli, P., Davarinejad, O., Dürsteler, K.M., Brühl, A.B., Sadeghi-Bahmani, D., Brand, S., 2021. Sources of Sleep Disturbances and Psychological Strain for Hospital Staff Working during the COVID-19 Pandemic. International journal of environmental research and public health 18.

AbuDujain, N.M., Almuhaideb, Q.A., Alrumaihi, N.A., Alrabiah, M.A., Alanazy, M.H., Abdulghani, H., 2021. The Impact of the COVID-19 Pandemic on Medical Interns' Education, Training, and Mental Health: A Cross-Sectional Study. Cureus 13, e19250.

Abu-Elenin, M.M., 2021. Immediate psychological outcomes associated with COVID-19 pandemic in frontline physicians: A cross-sectional study in Egypt. BMC Psychiatry 21.

Abu-Snieneh, H.M., 2021. Psychological factors associated with the spread of Coronavirus disease 2019 (COVID-19) among nurses working in health sectors in Saudi Arabia. Perspectives in psychiatric care 57, 1399-1408.

Acar Sevinc, S., Metin, S., Balta Basi, N., Cinar, A.S., Turkel Ozkan, M., Oba, S., 2021. Anxiety and burnout in anesthetists and intensive care unit nurses during the COVID-19 pandemic: a cross-sectional study. Brazilian journal of anesthesiology (Elsevier).

Aggar, C., Samios, C., Penman, O., Whiteing, N., Massey, D., Rafferty, R., Bowen, K., Stephens, A., 2021. The impact of covid‐19 pandemic‐related stress experienced by australian nurses. International Journal of Mental Health Nursing.

Ahn, M.H., Shin, Y.-W., Suh, S., Kim, J.H., Kim, H.J., Lee, K.-U., Chung, S., 2021. High Work-Related Stress and Anxiety as a Response to COVID-19 Among Health Care Workers in South Korea: Cross-sectional Online Survey Study. JMIR public health and surveillance 7, e25489.

Ajab, S., Ádam, B., Al Hammadi, M., Al Bastaki, N., Al Junaibi, M., Al Zubaidi, A., Hegazi, M., Grivna, M., Kady, S., Koornneef, E., Neves, R., Uva, A.S., Sheek-Hussein, M., Loney, T., Serranheira, F., Paulo, M.S., 2021. Occupational Health of Frontline Healthcare Workers in the United Arab Emirates during the COVID-19 Pandemic: A Snapshot of Summer 2020. International journal of environmental research and public health 18.

Al Ammari, M., Sultana, K., Thomas, A., Al Swaidan, L., Al Harthi, N., 2020. Mental Health Outcomes Amongst Health Care Workers During COVID 19 Pandemic in Saudi Arabia. Frontiers in Psychiatry 11.

Al Hariri, M., Hamade, B., Bizri, M., Salman, O., Tamim, H., Al Jalbout, N., 2022. Psychological impact of COVID-19 on emergency department healthcare workers in a tertiary care center during a national economic crisis. AMERICAN JOURNAL OF EMERGENCY MEDICINE 51, 342-347.

Al Maqbali, M., 2021. Sleep disturbance among frontline nurses during the COVID-19 pandemic. Sleep and Biological Rhythms 19, 467-473.

Alan, H., Eskin Bacaksiz, F., Tiryaki Sen, H., Taskiran Eskici, G., Gumus, E., Harmanci Seren, A.K., 2020. 'i'm a hero, but…': An evaluation of depression, anxiety, and stress levels of frontline healthcare professionals during covid‐19 pandemic in turkey. Perspectives in Psychiatric Care.

AlAteeq, D.A., Aljhani, S., Althiyabi, I., Majzoub, S., 2020. Mental health among healthcare providers during coronavirus disease (COVID-19) outbreak in Saudi Arabia. Journal of Infection and Public Health 13, 1432-1437.

Algattas, H., Roy, S., Agarwal, N., Maroon, J., 2021. COVID-19 Impact in Neurosurgery Residency: Grit During Pandemic. World neurosurgery 151, e395-e398.

Alghasab, N.S., Aljadani, A.H., Almesned, S.S., Hersi, A.S., 2021. Depression among physicians and other medical employees involved in the COVID-19 outbreak: A cross-sectional study. Medicine 100, e25290-e25290.

Alhalaiqa, F.N., Khalifeh, A.H., Al Omari, O., Yehia, D.B., Khalil, M.M.H., 2021. Psychological Problems in a Sample of Jordanian Healthcare Workers Involved in Caring for Patients With COVID-19: A Cross-Sectional Study. Frontiers in psychology 12, 679785.

Al-Humadi, S., Bronson, B., Muhlrad, S., Paulus, M., Hong, H., Cáceda, R., 2021. Depression, suicidal thoughts, and burnout among physicians during the covid-19 pandemic: A survey-based cross-sectional study. Academic Psychiatry.

Al-Humadi, S.M., Cáceda, R., Bronson, B., Paulus, M., Hong, H., Muhlrad, S., 2021. Orthopaedic Surgeon Mental Health During the COVID-19 Pandemic. Geriatric orthopaedic surgery & rehabilitation 12, 21514593211035230.

Ali, M., Uddin, Z., Ahsan, N.F., Haque, M.Z., Bairagee, M., Khan, S.A., Hossain, A., 2021. Prevalence of mental health symptoms and its effect on insomnia among healthcare workers who attended hospitals during COVID-19 pandemic: A survey in Dhaka city. Heliyon 7, e06985.

Ali, S., Maguire, S., Marks, E., Doyle, M., Sheehy, C., 2020. Psychological impact of the COVID-19 pandemic on healthcare workers at acute hospital settings in the South-East of Ireland: An observational cohort multicentre study. BMJ Open 10.

Alkhamees, A.A., Assiri, H., Alharbi, H.Y., Nasser, A., Alkhamees, M.A., 2021. Burnout and depression among psychiatry residents during COVID-19 pandemic. Human Resources for Health 19, 1-9.

Almater, A., Tobaigy, M., Younis, A., Alaqeel, M., Abouammoh, M., 2020. Effect of 2019 coronavirus pandemic on ophthalmologists practicing in Saudi Arabia: A psychological health assessment. Middle East African Journal of Ophthalmology 27, 79-85.

Alonso, J., Vilagut, G., Mortier, P., Ferrer, M., Alayo, I., Aragón-Peña, A., Aragonès, E., Campos, M., Cura-González, I.D., Emparanza, J.I., Espuga, M., Forjaz, M.J., González-Pinto, A., Haro, J.M., López-Fresneña, N., Salázar, A.D.M.d., Molina, J.D., Ortí-Lucas, R.M., Parellada, M., Pelayo-Terán, J.M., Pérez-Zapata, A., Pijoan, J.I., Plana, N., Puig, M.T., Rius, C., Rodríguez-Blázquez, C., Sanz, F., Serra, C., Kessler, R.C., Bruffaerts, R., Vieta, E., Pérez-Solà, V., 2021. Mental health impact of the first wave of COVID-19 pandemic on Spanish healthcare workers: A large cross-sectional survey. Revista de psiquiatria y salud mental 14, 90-105.

Alsairafi, Z., Naser, A.Y., Alsaleh, F.M., Awad, A., Jalal, Z., 2021. Mental Health Status of Healthcare Professionals and Students of Health Sciences Faculties in Kuwait during the COVID-19 Pandemic. International journal of environmental research and public health 18.

Alshekaili, M., Hassan, W., Al Said, N., Al Sulaimani, F., Jayapal, S.K., Al-Mawali, A., Chan, M.F., Mahadevan, S., Al-Adawi, S., 2020. Factors associated with mental health outcomes across healthcare settings in Oman during COVID-19: Frontline versus non-frontline healthcare workers. BMJ Open 10.

Altmayer, V., Weiss, N., Cao, A., Marois, C., Demeret, S., Rohaut, B., Le Guennec, L., 2021. Coronavirus disease 2019 crisis in Paris: A differential psychological impact between regular intensive care unit staff members and reinforcement workers. Australian Critical Care 34, 142-145.

Aly, H.M., Nemr, N.A., Kishk, R.M., Elsaid, N.M.A.B., 2021. Stress, anxiety and depression among healthcare workers facing COVID-19 pandemic in Egypt: a cross-sectional online-based study. BMJ open 11, e045281.

Alzaid, E.H., Alsaad, S.S., Alshakhis, N., Albagshi, D., Albesher, R., Aloqaili, M., 2020. Prevalence of COVID-19-related anxiety among healthcare workers: A cross-sectional study. Journal of family medicine and primary care 9, 4904-4910.

Amer, S.A.A.M., Fouad, A.M., El-Samahy, M., Hashem, A.A., saati, A.A., Sarhan, A.A., Anani, M., 2021. Mental Stress, Anxiety and Depressive Symptoms and Interleuken-6 Level among Healthcare Workers during the COVID-19 Pandemic. Journal of Primary Care & Community Health, 1-8.

Amra, B., Salmasi, M., Soltaninejad, F., Sami, R., Nickpour, M., Mansourian, M., Ghasemi, K., Morin, C.M., 2021. Healthcare workers' sleep and mood disturbances during COVID-19 outbreak in an Iranian referral center. Sleep & breathing = Schlaf & Atmung 25, 2197-2204.

An, Y., Yang, Y., Wang, A., Li, Y., Zhang, Q., Cheung, T., Ungvari, G.S., Qin, M.Z., An, F.R., Xiang, Y.T., 2020. Prevalence of depression and its impact on quality of life among frontline nurses in emergency departments during the COVID-19 outbreak. Journal of Affective Disorders 276, 312-315.

Appiani, F.J., Rodríguez Cairoli, F., Sarotto, L., Yaryour, C., Basile, M.E., Duarte, J.M., 2021. Prevalence of stress, burnout syndrome, anxiety and depression among physicians of a teaching hospital during the COVID-19 pandemic. Archivos argentinos de pediatria 119, 317-324.

Arafa, A., Mohammed, Z., Mahmoud, O., Elshazley, M., Ewis, A., 2020. Depressed, anxious, and stressed: What have healthcare workers on the frontlines in Egypt and Saudi Arabia experienced during the COVID-19 pandemic? Journal of affective disorders 278, 365-371.

Arca, M., Dönmezdil, S., Durmaz, E.D., 2021. The effect of the COVID-19 Pandemic on anxiety, depression, and musculoskeletal system complaints in healthcare workers. Work (Reading, Mass.) 69, 47-54.

Arnetz, J.E., Goetz, C.M., Sudan, S., Arble, E., Janisse, J., Arnetz, B.B., 2020. Personal Protective Equipment and Mental Health Symptoms Among Nurses During the COVID-19 Pandemic. Journal of occupational and environmental medicine 62, 892-897.

Asnakew, S., Legas, G., Muche Liyeh, T., Belete, A., Haile, K., Yitbarek, G.Y., Bayih, W.A., Feleke, D.G., Birhane, B.M., Amha, H., Shumet, S., Chanie, E.S., 2021. Prevalence of post-traumatic stress disorder on health professionals in the era of COVID-19 pandemic, Northwest Ethiopia, 2020: A multi-centered cross-sectional study. PloS one 16, e0255340.

Awano, N., Oyama, N., Akiyama, K., Inomata, M., Kuse, N., Tone, M., Takada, K., Muto, Y., Fujimoto, K., Akagi, Y., Mawatari, M., Ueda, A., Kawakami, J., Komatsu, J., Izumo, T., 2020. Anxiety, depression, and resilience of healthcare workers in japan during the coronavirus disease 2019 outbreak. Internal Medicine 59, 2693-2699.

Ayalew, M., Deribe, B., Abraham, Y., Reta, Y., Tadesse, F., Defar, S., Hoyiso, D., Ashegu, T., 2021. Prevalence and determinant factors of mental health problems among healthcare professionals during COVID-19 pandemic in southern Ethiopia: multicentre cross-sectional study. BMJ open 11, e057708.

Azizoddin, D.R., Kvaternik, N., Beck, M., Zhou, G., Hasdianda, M.A., Jones, N., Johnsky, L., Im, D., Chai, P.R., Boyer, E.W., 2021. Heal the Healers: A pilot study evaluating the feasibility, acceptability, and exploratory efficacy of a Transcendental Meditation intervention for emergency clinicians during the coronavirus disease 2019 pandemic. Journal of the American College of Emergency Physicians open 2, e12619.

Azoulay, E., Cariou, A., Bruneel, F., Demoule, A., Kouatchet, A., Reuter, D., Souppart, V., Combes, A., Klouche, K., Argaud, L., Barbier, F., Jourdain, M., Reignier, J., Papazian, L., Guidet, B., Géri, G., Resche-Rigon, M., Guisset, O., Labbé, V., Mégarbane, B., Van Der Meersch, G., Guitton, C., Friedman, D., Pochard, F., Darmon, M., Kentish-Barnes, N., 2020. Symptoms of Anxiety, Depression, and Peritraumatic Dissociation in Critical Care Clinicians Managing Patients with COVID-19. A Cross-Sectional Study. American journal of respiratory and critical care medicine 202, 1388-1398.

Azoulay, E., De Waele, J., Ferrer, R., Staudinger, T., Borkowska, M., Povoa, P., Iliopoulou, K., Artigas, A., Schaller, S.J., Hari, M.S., Pellegrini, M., Darmon, M., Kesecioglu, J., Cecconi, M., 2020. Symptoms of burnout in intensive care unit specialists facing the COVID-19 outbreak. Annals of Intensive Care 10.

Azoulay, E., Pochard, F., Reignier, J., Argaud, L., Bruneel, F., Courbon, P., Cariou, A., Klouche, K., Labbé, V., Barbier, F., Guitton, C., Demoule, A., Kouatchet, A., Guisset, O., Jourdain, M., Papazian, L., Van Der Meersch, G., Reuter, D., Souppart, V., Resche-Rigon, M., 2021. Symptoms of Mental Health Disorders in Critical Care Physicians Facing the Second COVID-19 Wave: A Cross-Sectional Study. CHEST 160, 944-955.

Badahdah, A., Khamis, F., Al Mahyijari, N., Al Balushi, M., Al Hatmi, H., Al Salmi, I., Albulushi, Z., Al Noomani, J., 2020. The mental health of health care workers in Oman during the COVID-19 pandemic. The International journal of social psychiatry, 20764020939596.

Bahadirli, S., Sagaltici, E., 2021. Burnout, Job Satisfaction, and Psychological Symptoms Among Emergency Physicians During COVID-19 Outbreak: A Cross-Sectional Study. PSYCHIATRY AND CLINICAL PSYCHOPHARMACOLOGY 31, 67-76.

Bahadirli, S., Sagaltici, E., 2021. Post-traumatic stress disorder in healthcare workers of emergency departments during the pandemic: A cross-sectional study. American Journal of Emergency Medicine 50, 251-255.

Bajo, M., Gallego, P., Stavraki, M., Lamprinakos, G., Luna, P., Díaz, D., 2021. Anxiety, trauma and well-being in health-care professionals during COVID-19 first wave in Spain: the moderating role of personal protection equipment availability. Health & Quality of Life Outcomes 19, 1-9.

Balay-Odao, E.M., Alquwez, N., Inocian, E.P., Alotaibi, R.S., 2021. Hospital Preparedness, Resilience, and Psychological Burden Among Clinical Nurses in Addressing the COVID-19 Crisis in Riyadh, Saudi Arabia. Frontiers in public health 8, 573932.

Baminiwatta, A., De Silva, S., Hapangama, A., Basnayake, K., Abayaweera, C., Kulasinghe, D., Kaushalya, D., Williams, S., 2021. Impact of COVID-19 on the mental health of frontline and non-frontline healthcare workers in Sri Lanka. Ceylon Medical Journal 66, 16-31.

Bani Issa, W., Al Nusair, H., AlTamimi, A., Rababa, M., Saqan, R., Hijazi, H., Al-Marzouqi, A., Abdul Rahman, H., Naing, L., 2021. Posttraumatic stress disorders and influencing factors during the COVID-19 pandemic: A cross-sectional study of frontline nurses. International nursing review.

Bashir, M., Ahluwalia, H., Sayeed, S.I., Salroo, I.N., 2020. A Study of Depressive Symptoms in Doctors Working at COVID-19 Hospitals: An Online Survey. Medeniyet medical journal 35, 310-314.

Bates, A., Ottaway, J., Moyses, H., Perrrow, M., Rushbrook, S., Cusack, R., 2020. Psychological impact of caring for critically ill patients during the Covid-19 pandemic and recommendations for staff support. Journal of the Intensive Care Society.

Baumann, B.M., Cooper, R.J., Medak, A.J., Lim, S., Chinnock, B., Frazier, R., Roberts, B.W., Epel, E.S., Rodriguez, R.M., Mycyk, M.B., 2021. Emergency physician stressors, concerns, and behavioral changes during COVID‐19: A longitudinal study. Academic Emergency Medicine 28, 314-324.

Besirli, A., Erden, S.C., Atilgan, M., Varlihan, A., Habaci, M.F., Yeniceri, T., Isler, A.C., Gumus, M., Kizileroglu, S., Ozturk, G., Ozer, O.A., Ozdemir, H.M., 2021. The Relationship between Anxiety and Depression Levels with Perceived Stress and Coping Strategies in Health Care Workers during the COVID-19 Pandemic. Sisli Etfal Hastanesi tip bulteni 55, 1-11.

Brejnebøl, M.W., Walvik, L., Christensen, A.K., Jensen, R.G., von Buchwald, C., 2021. Stress reactions in a tertiary oto-rhino-laryngological department during the first wave of the COVID-19 pandemic in the Danish Capital region. Acta Oto-Laryngologica 141, 791-795.

Bulut, D., Sefa Sayar, M., Koparal, B., Cem Bulut, E., Çelik, S., 2021. Which of us were more affected by the pandemic? The psychiatric impacts of the COVID-19 pandemic on healthcare professionals in the province where the first quarantine units were established in Turkey. International journal of clinical practice 75, e14235.

Cag, Y., Erdem, H., Gormez, A., Ankarali, H., Hargreaves, S., Ferreira-Coimbra, J., Rubulotta, F., Belliato, M., Berger-Estilita, J., Pelosi, P., Blot, S., Lefrant, J.Y., Mardani, M., Darazam, I.A., Cag, Y., Rello, J., 2021. Anxiety among front-line health-care workers supporting patients with COVID-19: A global survey. General Hospital Psychiatry 68, 90-96.

Cai, Q., Feng, H., Huang, J., Wang, M., Wang, Q., Lu, X., Xie, Y., Wang, X., Liu, Z., Hou, B., Ouyang, K., Pan, J., Li, Q., Fu, B., Deng, Y., Liu, Y., 2020. The mental health of frontline and non-frontline medical workers during the coronavirus disease 2019 (COVID-19) outbreak in China: A case-control study. Journal of Affective Disorders 275, 210-215.

Cai, Z., Cui, Q., Liu, Z., Li, J., Gong, X., Liu, J., Wan, Z., Yuan, X., Li, X., Chen, C., Wang, G., 2020. Nurses endured high risks of psychological problems under the epidemic of COVID-19 in a longitudinal study in Wuhan China. Journal of Psychiatric Research 131, 132-137.

Caillet, A., Coste, C., Sanchez, R., Allaouchiche, B., 2020. Psychological Impact of COVID-19 on ICU Caregivers. Anaesthesia Critical Care and Pain Medicine.

Caliandro, M., Fabiana, G., Surgo, A., Carbonara, R., Ciliberti, M.P., Bonaparte, I., Caputo, S., Fiorentino, A., 2022. Impact on mental health of the COVID-19 pandemic in a radiation oncology department. RADIOLOGIA MEDICA 127, 220-224.

Çalişkan, F., Dost, B., 2020. The evaluation of knowledge, attitudes, depression and anxiety levels among emergency physicians during the COVID-19 pandemic. Signa Vitae 16, 163-171.

Carmassi, C., Bui, E., Bertelloni, C.A., Dell'Oste, V., Pedrinelli, V., Corsi, M., Baldanzi, S., Cristaudo, A., Dell'Osso, L., Buselli, R., 2021. Validation of the Italian version of the Peritraumatic Distress Inventory: Validity, reliability and factor analysis in a sample of healthcare workers. European Journal of Psychotraumatology 12.

Carmassi, C., Pedrinelli, V., Dell'Oste, V., Bertelloni, C.A., Cordone, A., Bouanani, S., Corsi, M., Baldanzi, S., Malacarne, P., Dell'Osso, L., Buselli, R., 2021. Work and social functioning in frontline healthcare workers during the covid-19 pandemic in Italy: role of acute post-traumatic stress, depressive and anxiety symptoms. Rivista di psichiatria 56, 189-197.

Castioni, D., Galasso, O., Rava, A., Massè, A., Gasparini, G., Mercurio, M., 2021. Has the COVID-19 Pandemic Changed the Daily Practices and Psychological State of Orthopaedic Residents? Clinical Orthopaedics & Related Research®, 1947-1954.

ÇElİK, F., DaĞLi, R., 2021. Comparison of the Mental Status of COVID-19 Intensive Care Unit and General Intensive Care Unit Staff. Duzce Medical Journal 23, 197-204.

Chapa-Koloffon, G.D.C., Jean-Tron, M.G., Ávila-Hernández, A.V., Márquez-González, H., Garduño-Espinosa, J., 2021. Frequency of acute stress disorder in health care workers of a tertiary level pediatric hospital during the National Safe Distance Strategy for COVID-19 prevention. Boletin medico del Hospital Infantil de Mexico 78, 10-17.

Chauhan, V.S., Chatterjee, K., Yadav, A.K., Srivastava, K., Prakash, J., Yadav, P., Dangi, A., 2021. Mental health impact of COVID-19 among health-care workers: An exposure-based cross-sectional study. Industrial psychiatry journal 30, S63-S68.

Chen, H., Wang, B., Cheng, Y., Muhammad, B., Li, S., Miao, Z., Wan, B., Abdul, M., Zhao, Z., Geng, D., Xu, X., 2020. Prevalence of posttraumatic stress symptoms in health care workers after exposure to patients with COVID-19. Neurobiology of Stress 13.

Chen, J., Liu, X.H., Wang, D.K., Jin, Y., He, M., Ma, Y.L., Zhao, X.L., Song, S.N., Zhang, L., Xiang, X.L., Yang, L., Song, J., Bai, T., Hou, X.H., Risk factors for depression and anxiety in healthcare workers deployed during the COVID-19 outbreak in China. Social Psychiatry and Psychiatric Epidemiology.

Chen, R., Sun, C., Chen, J.-J., Jen, H.-J., Kang, X.L., Kao, C.-C., Chou, K.-R., 2021. A Large-Scale Survey on Trauma, Burnout, and Posttraumatic Growth among Nurses during the COVID-19 Pandemic. International journal of mental health nursing 30, 102-116.

Chene, G., Nohuz, E., Cerruto, E., Moret, S., Atallah, A., Saoud, M., 2021. Psychological impact on healthcare workers in obstetrics and gynecology in France in 18 French University Hospitals during the first Covid-19 lockdown: a prospective observational study. Journal of psychosomatic obstetrics and gynaecology, 1-8.

Cheng, F.F., Zhan, S.H., Xie, A.W., Cai, S.Z., Hui, L., Kong, X.X., Tian, J.M., Yan, W.H., 2020. Anxiety in Chinese pediatric medical staff during the outbreak of Coronavirus Disease 2019: A cross-sectional study. Translational Pediatrics 9, 231-236.

Cheng, P., Xu, L.Z., Zheng, W.H., Ng, R.M.K., Zhang, L., Li, L.J., Li, W.H., 2020. Psychometric property study of the posttraumatic stress disorder checklist for DSM-5 (PCL-5) in Chinese healthcare workers during the outbreak of corona virus disease 2019. Journal of Affective Disorders 277, 368-374.

Chew, N.W.S., Lee, G.K.H., Tan, B.Y.Q., Jing, M., Goh, Y., Ngiam, N.J.H., Yeo, L.L.L., Ahmad, A., Ahmed Khan, F., Napolean Shanmugam, G., Sharma, A.K., Komalkumar, R.N., Meenakshi, P.V., Shah, K., Patel, B., Chan, B.P.L., Sunny, S., Chandra, B., Ong, J.J.Y., Paliwal, P.R., Wong, L.Y.H., Sagayanathan, R., Chen, J.T., Ying Ng, A.Y., Teoh, H.L., Tsivgoulis, G., Ho, C.S., Ho, R.C., Sharma, V.K., 2020. A multinational, multicentre study on the psychological outcomes and associated physical symptoms amongst healthcare workers during COVID-19 outbreak. Brain, Behavior, and Immunity 88, 559-565.

Chew, N.W.S., Ngiam, J.N., Tan, B.Y.Q., Tham, S.M., Tan, C.Y.S., Jing, M.X., Sagayanathan, R., Chen, J.T., Wong, L.Y.H., Ahmad, A., Khan, F.A., Marmin, M., Hassan, F.B., Sharon, T.M.L., Lim, C.H., Bin Mohaini, M.I., Danuaji, R., Nguyen, T.H., Tsivgoulis, G., Tsiodras, S., Fragkou, P.C., Dimopoulou, D., Sharma, A.K., Shah, K., Patel, B., Sharma, S., Komalkumar, R.N., Meenakshi, R.V., Talati, S., Teoh, H.L., Ho, C.S., Ho, R.C., Sharma, V.K., 2020. Asian-Pacific perspective on the psychological well-being of healthcare workers during the evolution of the COVID-19 pandemic. Bjpsych Open 6.

Chou, D.W., Staltari, G., Mullen, M., Chang, J., Durr, M., 2021. Otolaryngology Resident Wellness, Training, and Education in the Early Phase of the COVID-19 Pandemic. Annals of Otology, Rhinology & Laryngology 130, 904-914.

Chow, S.K., Francis, B., Ng, Y.H., Naim, N., Beh, H.C., Ariffin, M.A.A., Yusuf, M.H.M., Lee, J.W., Sulaiman, A.H., 2021. Religious Coping, Depression and Anxiety among Healthcare Workers during the COVID-19 Pandemic: A Malaysian Perspective. Healthcare (Basel, Switzerland) 9.

Chung, S., Kim, H.J., Ahn, M.H., Yeo, S., Lee, J., Kim, K., Kang, S., Suh, S., Shin, Y.-W., 2021. Development of the Stress and Anxiety to Viral Epidemics-9 (SAVE-9) Scale for Assessing Work-related Stress and Anxiety in Healthcare Workers in Response to Viral Epidemics. Journal of Korean medical science 36, e319.

Cigiloglu, A., Ozturk, E., Ganidagli, S., Ozturk, Z.A., 2021. Different reflections of the face mask: sleepiness, headache and psychological symptoms. International journal of occupational safety and ergonomics : JOSE, 1-6.

Civantos, A.M., Bertelli, A., Gonçalves, A., Getzen, E., Chang, C., Long, Q., Rajasekaran, K., 2020. Mental health among head and neck surgeons in Brazil during the COVID-19 pandemic: A national study. American Journal of Otolaryngology - Head and Neck Medicine and Surgery 41.

Civantos, A.M., Byrnes, Y., Chang, C., Prasad, A., Chorath, K., Poonia, S.K., Jenks, C.M., Bur, A.M., Thakkar, P., Graboyes, E.M., Seth, R., Trosman, S., Wong, A., Laitman, B.M., Harris, B.N., Shah, J., Stubbs, V., Choby, G., Long, Q., Rassekh, C.H., Thaler, E., Rajasekaran, K., 2020. Mental health among otolaryngology resident and attending physicians during the COVID-19 pandemic: National study. Head and Neck 42, 1597-1609.

Collantoni, E., Saieva, A.M., Meregalli, V., Girotto, C., Carretta, G., Boemo, D.G., Bordignon, G., Capizzi, A., Contessa, C., Nesoti, M.V., Donato, D., Flesia, L., Favaro, A., 2021. Psychological Distress, Fear of COVID-19, and Resilient Coping Abilities among Healthcare Workers in a Tertiary First-Line Hospital during the Coronavirus Pandemic. Journal of clinical medicine 10.

Collins, C., Mahuron, K., Bongiovanni, T., Lancaster, E., Sosa, J.A., Wick, E., 2020. Stress and the Surgical Resident in the COVID-19 Pandemic. Journal of surgical education.

Conti, C., Fontanesi, L., Lanzara, R., Rosa, I., Doyle, R.L., Porcelli, P., 2021. Burnout Status of Italian Healthcare Workers during the First COVID-19 Pandemic Peak Period. Healthcare (Basel, Switzerland) 9.

Conti, C., Fontanesi, L., Lanzara, R., Rosa, I., Porcelli, P., 2020. Fragile heroes. The psychological impact of the COVID-19 pandemic on health-care workers in Italy. PloS one 15, e0242538.

Cornelius, T., Duran, A.T., Diaz, F., Bramley, S., Shaw, K., Schwartz, J.E., Edmondson, D., Shechter, A., Abdalla, M., 2021. The association of transmission concerns and social distance from loved ones with distress in medical professionals providing care during the COVID-19 pandemic in New York City. Families, Systems, & Health 39, 499-504.

Crotty, T.J., Corbett, M., Gary, S., Davey, M.G., Hughes, J.P., Keogh, I.J., Patil, N.P., Doherty, E., 2022. The psychological impact of COVID-19 on ear, nose and throat (ENT) specialists. IRISH JOURNAL OF MEDICAL SCIENCE 191, 51-57.

Crowe, S., Howard, A.F., Vanderspank-Wright, B., Gillis, P., McLeod, F., Penner, C., Haljan, G., 2021. The effect of COVID-19 pandemic on the mental health of Canadian critical care nurses providing patient care during the early phase pandemic: A mixed method study. Intensive & Critical Care Nursing 63, N.PAG-N.PAG.

Cui, Q., Cai, Z., Li, J., Liu, Z., Sun, S., Chen, C., Wang, G., 2020. The Psychological Pressures of Breast Cancer Patients During the COVID-19 Outbreak in China—A Comparison With Frontline Female Nurses. Frontiers in Psychiatry 11.

Cui, S., Jiang, Y., Shi, Q., Zhang, L., Kong, D., Qian, M., Chu, J., 2021. Impact of COVID-19 on Anxiety, Stress, and Coping Styles in Nurses in Emergency Departments and Fever Clinics: A Cross-Sectional Survey. Risk management and healthcare policy 14, 585-594.

Dal'Bosco, E.B., Floriano, L.S.M., Skupien, S.V., Arcaro, G., Martins, A.R., Anselmo, A.C.C., 2020. Mental health of nursing in coping with COVID-19 at a regional university hospital. Revista Brasileira De Enfermagem 73.

Dantas, E.S.O., Araújo Filho, J.d.D.d., Silva, G.W.D.S., Silveira, M.Y.M., Dantas, M.N.P., Meira, K.C., 2021. Factors associated with anxiety in multiprofessional health care residents during the COVID-19 pandemic. Revista brasileira de enfermagem 74, e20200961.

Das, K., Ryali, V.S.S.R., Bhavyasree, R., Sekhar, C.M., 2021. Postexposure psychological sequelae in frontline health workers to COVID-19 in Andhra Pradesh, India. Industrial psychiatry journal 30, 123-130.

Daviskiba, S.E., MacKenzie, M.A., Dow, M., Johnston, P., Balon, R., Javanbakht, A., Arfken, C.L., 2021. Rapid assessment of mental health of Detroit-area health care workers during the COVID-19 pandemic. Annals of clinical psychiatry : official journal of the American Academy of Clinical Psychiatrists 33, 101-107.

De Kock, J.H., Latham, H.A., Cowden, R.G., Cullen, B., Narzisi, K., Jerdan, S., Munoz, S.-A., Leslie, S.J., McNamara, N., Boggon, A., Humphry, R.W., 2022. The mental health of NHS staff during the COVID-19 pandemic: two-wave Scottish cohort study. BJPSYCH OPEN 8.

Debnath, P.R., Islam, M.S., Karmakar, P.K., Sarker, R., Zhai, Z.W., Potenza, M.N., 2021. Mental health concerns, insomnia, and loneliness among intern doctors amidst the covid-19 pandemic: Evidence from a large tertiary care hospital in bangladesh. International Journal of Mental Health and Addiction.

Debski, M., Abdelaziz, H.K., Sanderson, J., Wild, S., Assaf, O., Wiper, A., Nabi, A., Abdelrahman, A., Eichhofer, J., Skailes, G., Gardner, J., Moynes, K., Goode, G., Pathan, T., Patel, B., Kumar, S., Taylor, R., Galasko, G., More, R., Chalil, S., Choudhury, T., 2021. Mental Health Outcomes Among British Healthcare Workers-Lessons From the First Wave of the Covid-19 Pandemic. Journal of occupational and environmental medicine 63, e549-e555.

Dehon, E., Zachrison, K.S., Peltzer-Jones, J., Tabatabai, R.R., Clair, E., Puskarich, M.A., Ondeyka, A., Dixon-Gordon, K., Walter, L.A., Situ-LaCasse, E.H., Fix, M.L., 2021. Sources of Distress and Coping Strategies Among Emergency Physicians During COVID-19. Western Journal of Emergency Medicine: Integrating Emergency Care with Population Health 22, 1240-1252.

Diaz, F., Bramley, S., Venner, H., Shaw, K., McMurry, C., Cornelius, T., Schwartz, J., Shechter, A., Abdalla, M., 2021. THE ASSOCIATION BETWEEN SLEEP AND PSYCHOLOGICAL DISTRESS AMONG NEW YORK HEALTHCARE WORKERS DURING THE COVID-19 PANDEMIC. SLEEP 44, A254-A254.

Doan, Q.-H., Tran, N.-N., Than, M.-H., Nguyen, H.-T., Bui, V.-S., Nguyen, D.-H., Vo, H.-L., Do, T.-T., Pham, N.-T., Nguyen, T.-K., Cao, D.-C., Nguyen, V.-T., Tran, T.-M.T., Pham, B.-H., Tran, A.-L., Nguyen, V.-T., Nguyen, V.-T., Tran, X.-T., Lai, D.-T., Vu, Q.-H., Otsu, S., 2021. Depression, Anxiety and Associated Factors among Frontline Hospital Healthcare Workers in the Fourth Wave of COVID-19: Empirical Findings from Vietnam. Tropical medicine and infectious disease 7.

Dobson, H., Malpas, C.B., Burrell, A.J.C., Gurvich, C., Chen, L., Kulkarni, J., Winton-Brown, T., 2020. Burnout and psychological distress amongst Australian healthcare workers during the COVID-19 pandemic. Australasian Psychiatry.

Dolev, T., Zubedat, S., Brand, Z., Bloch, B., Mader, E., Blondheim, O., Avital, A., 2021. Physiological parameters of mental health predict the emergence of post-traumatic stress symptoms in physicians treating COVID-19 patients. Translational psychiatry 11, 169.

Dong, H.-S., Gao, J.-J., Dong, Y.-X., Han, C.-X., Sun, L., 2021. Prevalence of insomnia and anxiety among healthcare workers during the COVID-19 pandemic in Jilin Province. Brazilian journal of medical and biological research = Revista brasileira de pesquisas medicas e biologicas 54, e10602.

Doo, E.Y., Kim, M., Lee, S., Lee, S.Y., Lee, K.Y., 2021. Influence of anxiety and resilience on depression among hospital nurses: A comparison of nurses working with confirmed and suspected patients in the COVID‐19 and non‐COVID‐19 units. Journal of Clinical Nursing (John Wiley & Sons, Inc.) 30, 1990-2000.

Doulias, T., Thrikandiyur, A.A., Titus, N., Soundararasha, K., Coxon, A., Amarantidis, E., Arulampalam, T., 2021. Junior doctors' wellbeing at peak and post-peak pandemic: a repeated cross-sectional study. Annals of the Royal College of Surgeons of England.

Dudine, L., Canaletti, C., Giudici, F., Lunardelli, A., Abram, G., Santini, I., Baroni, V., Paris, M., Pesavento, V., Manganotti, P., Ronchese, F., Gregoretti, B., Negro, C., 2021. Investigation on the Loss of Taste and Smell and Consequent Psychological Effects: A Cross-Sectional Study on Healthcare Workers Who Contracted the COVID-19 Infection. Frontiers in public health 9, 666442.

Duru, H., 2021. The Continuing Effect of COVID-19 Pandemic on Physical Well-Being and Mental Health of ICU Healthcare Workers in Turkey: A Single-Centre Cross-Sectional Later-Phase Study. Journal of intensive care medicine, 8850666211070740.

Elgohary, H.M., Sehlo, M.G., Bassiony, M.M., Youssef, U.M., Elrafey, D.S., Amin, S.I., 2021. Depression among health workers caring for patients with COVID-19 in Egypt. The Egyptian journal of neurology, psychiatry and neurosurgery 57, 139.

Elhadi, M., Msherghi, A., Elgzairi, M., Alhashimi, A., Bouhuwaish, A., Biala, M., Abuelmeda, S., Khel, S., Khaled, A., Alsoufi, A., Elhadi, A., BenGhatnsh, A., 2020. The Mental Well-Being of Frontline Physicians Working in Civil Wars Under Coronavirus Disease 2019 Pandemic Conditions. Frontiers in Psychiatry 11.

Elkholy, H., Tawfik, F., Ibrahim, I., El-din, W.S., Sabry, M., Mohammed, S., Hamza, M., Alaa, M., Fawzy, A., Ashmawy, R., Sayed, M., Omar, A.N., Mental health of frontline healthcare workers exposed to COVID-19 in Egypt: A call for action. International Journal of Social Psychiatry.

Ezzat, A., Li, Y., Holt, J., Komorowski, M., 2021. The global mental health burden of COVID-19 on critical care staff. British Journal of Nursing 30, 634-642.

Fang, X.-H., Wu, L., Lu, L.-S., Kan, X.-H., Wang, H., Xiong, Y.-J., Ma, D.-C., Wu, G.-C., 2021. Mental health problems and social supports in the COVID-19 healthcare workers: A Chinese explanatory study. BMC Psychiatry 21.

Fari, G., de Sire, A., Giorgio, V., Rizzo, L., Bruni, A., Bianchi, F.P., Zonno, A., Pierucci, P., Ranieri, M., Megna, M., 2022. Impact of COVID-19 on the mental health in a cohort of Italian rehabilitation healthcare workers. JOURNAL OF MEDICAL VIROLOGY 94, 110-118.

Fattori, A., Cantù, F., Comotti, A., Tombola, V., Colombo, E., Nava, C., Bordini, L., Riboldi, L., Bonzini, M., Brambilla, P., 2021. Hospital workers mental health during the COVID-19 pandemic: methods of data collection and characteristics of study sample in a university hospital in Milan (Italy). BMC Medical Research Methodology 21, 1-12.

Feingold, J.H., Hurtado, A., Feder, A., Peccoralo, L., Southwick, S.M., Ripp, J., Pietrzak, R.H., 2022. Posttraumatic growth among health care workers on the frontlines of the COVID-19 pandemic. JOURNAL OF AFFECTIVE DISORDERS 296, 35-40.

Feingold, J.H., Peccoralo, L., Chan, C.C., Kaplan, C.A., Kaye-Kauderer, H., Charney, D., Verity, J., Hurtado, A., Burka, L., Syed, S.A., Murrough, J.W., Feder, A., Pietrzak, R.H., Ripp, J., 2021. Psychological Impact of the COVID-19 Pandemic on Frontline Health Care Workers During the Pandemic Surge in New York City. Chronic stress (Thousand Oaks, Calif.) 5, 2470547020977891.

Flateau, C., Noël, C., Bonnafoux, A., Fuentes, E., de Pontfarcy, A., Diamantis, S., 2021. Psychological impact of the SARS-CoV-2 outbreak on the staff of a French hospital. Infectious diseases now 51, 187-193.

Florin, M., Pinar, U., Chavigny, E., Bouaboula, M., Jarboui, L., Coulibaly, A., Lemogne, C., Fournier, L., 2020. Socio-economic and psychological impact of the COVID-19 outbreak on private practice and public hospital radiologists. European Journal of Radiology 132.

Franzoi, I.G., Granieri, A., Sauta, M.D., Agnesone, M., Gonella, M., Cavallo, R., Lochner, P., Bragazzi, N.L., Naldi, A., 2021. Anxiety, Post-Traumatic Stress, and Burnout in Health Professionals during the COVID-19 Pandemic: Comparing Mental Health Professionals and Other Healthcare Workers. Healthcare (Basel, Switzerland) 9.

Fu, M., Han, D., Xu, M., Mao, C., Wang, D., 2021. The psychological impact of anxiety and depression on Chinese medical staff during the outbreak of the COVID-19 pandemic: a cross-sectional study. Annals of palliative medicine 10, 7759-7774.

Fukunaga, A., Inoue, Y., Yamamoto, S., Miki, T., Nanri, A., Ishiwari, H., Ishii, M., Miyo, K., Konishi, M., Ohmagari, N., Mizoue, T., 2021. Association Between Adherence to Healthy Lifestyles and Depressive Symptoms Among Japanese Hospital Workers During the COVID-19 Pandemic. Asia-Pacific Journal of Public Health 33, 847-853.

García-Hedrera, F.J., Gil-Almagro, F., Carmona-Monge, F.J., Peñacoba-Puente, C., Catalá-Mesón, P., Velasco-Furlong, L., 2021. Intensive care unit professionals during the COVID-19 pandemic in Spain: social and work-related variables, COVID-19 symptoms, worries, and generalized anxiety levels. Acute and critical care 36, 232-241.

Garg, S., Yadav, M., Chauhan, A., Verma, D., Bansal, K., 2020. Prevalence of psychological morbidities and their influential variables among nurses in a designated COVID-19 tertiary care hospital in India: A cross-sectional study. Industrial psychiatry journal 29, 237-244.

GebreEyesus, F.A., Tarekegn, T.T., Amlak, B.T., Shiferaw, B.Z., Emeria, M.S., Geleta, O.T., Terefe, T.F., Mammo Tadereregew, M., Jimma, M.S., Degu, F.S., Abdisa, E.N., Eshetu, M.A., Misganaw, N.M., Chanie, E.S., 2021. Levels and predictors of anxiety, depression, and stress during COVID-19 pandemic among frontline healthcare providers in Gurage zonal public hospitals, Southwest Ethiopia, 2020: A multicenter cross-sectional study. PloS one 16, e0259906.

Geng, S., Zhou, Y., Zhang, W., Lou, A., Cai, Y., Xie, J., Sun, J., Zhou, W., Liu, W., Li, X., 2021. The influence of risk perception for COVID‐19 pandemic on posttraumatic stress disorder in healthcare workers: A survey from four designated hospitals. Clinical Psychology & Psychotherapy 28, 1146-1159.

Ghaleb, Y., Lami, F., Al Nsour, M., Rashak, H.A., Samy, S., Khader, Y.S., Al Serouri, A., BahaaEldin, H., Afifi, S., Elfadul, M., Ikram, A., Akhtar, H., Hussein, A.M., Barkia, A., Hakim, H., Taha, H.A., Hijjo, Y., Kamal, E., Ahmed, A.Y., Rahman, F., Islam, K.M., Hussein, M.H., Ramzi, S.R., 2021. Mental health impacts of COVID-19 on healthcare workers in the Eastern Mediterranean Region: a multi-country study. Journal of public health (Oxford, England) 43, iii34-iii42.

Ghio, L., Patti, S., Piccinini, G., Modafferi, C., Lusetti, E., Mazzella, M., Del Sette, M., 2021. Anxiety, Depression and Risk of Post-Traumatic Stress Disorder in Health Workers: The Relationship with Burnout during COVID-19 Pandemic in Italy. International journal of environmental research and public health 18.

Gorini, A., Fiabane, E., Sommaruga, M., Barbieri, S., Sottotetti, F., La Rovere, M.T., Tremoli, E., Gabanelli, P., 2020. Mental health and risk perception among Italian healthcare workers during the second month of the Covid-19 pandemic. Archives of Psychiatric Nursing 34, 537-544.

Grover, S., Mehra, A., Sahoo, S., Avasthi, A., Sathyanarayana Rao, T.S., Vaishnav, M., Dalal, P.K., Saha, G., Singh, O.P., Chakraborty, K., Janardran Reddy, Y.C., Rao, N.P., Tripathi, A., Chadda, R.K., Mishra, K.K., Rao, G.P., Kumar, V., Gautam, S., Sarkar, S., Krishnan, V., 2021. Evaluation of Psychological Impact of COVID-19 on Health-Care Workers. Indian Journal of Psychiatry 63, 222-227.

Guiroy, A., Gagliardi, M., Coombes, N., Landriel, F., Zanardi, C., Camino Willhuber, G., Guyot, J.P., Valacco, M., 2020. COVID-19 Impact Among Spine Surgeons in Latin America. Global Spine Journal.

Gül, Ş., Kılıç, S.T., 2021. Determining anxiety levels and related factors in operating room nurses during the COVID-19 pandemic: A descriptive study. Journal of nursing management 29, 1934-1945.

Gundogdu, A., Ozsoy, S., Koyuncu, S., Uysal, C., Kocyigit, I., Sipahioglu, M.H., Tokgoz, B., Oymak, O., 2021. Psychological Effect of Coronavirus Disease-19 on Healthcare Workers: A Cross-Sectional Study in Kayseri. ERCIYES MEDICAL JOURNAL 43, 600-605.

Gündoğmuş, İ., Ünsal, C., Bolu, A., Takmaz, T., Ökten, S.B., Aydın, M.B., Uçar, H., Gündüz, A., Kul, A.T., 2021. The comparison of anxiety, depression and stress symptoms levels of healthcare workers between the first and second COVID-19 peaks. Psychiatry research 301, 113976.

Guo, W.-P., Min, Q., Gu, W.-W., Yu, L., Xiao, X., Yi, W.-B., Li, H.-L., Huang, B., Li, J.-L., Dai, Y.-J., Xia, J., Liu, J., Li, B., Zhou, B.-H., Li, M., Xu, H.-X., Wang, X.-B., Shi, W.-Y., 2021. Prevalence of mental health problems in frontline healthcare workers after the first outbreak of COVID-19 in China: a cross-sectional study. Health & Quality of Life Outcomes 19, 1-10.

Guo, X., McCutcheon, R., Pillinger, T., Arumuham, A., Chen, J., Ma, S., Yang, J., Wang, Y., Hu, S., Wang, G., Liu, Z.-C., 2021. Acute psychological impact of coronavirus disease 2019 outbreak among psychiatric professionals in China: a multicentre, cross-sectional, web-based study. BMJ open 11, e047828.

Gupta, P., B K, A., Ramakrishna, K., 2021. Prevalence of depression and anxiety among medical students and house staff during the covid-19 health-care crisis. Academic Psychiatry.

Hammami, A.S., Jellazi, M., Mahjoub, L., Fedhila, M., Ouali, S., 2021. Psychological Impact of the COVID-19 Pandemic on Healthcare Professionals in Tunisia: Risk and Protective Factors. Frontiers in psychology 12, 754047.

Hammond, N.E., Crowe, L., Abbenbroek, B., Elliott, R., Tian, D.H., Donaldson, L.H., Fitzgerald, E., Flower, O., Grattan, S., Harris, R., Sayers, L., Delaney, A., 2021. Impact of the coronavirus disease 2019 pandemic on critical care healthcare workers' depression, anxiety, and stress levels. Australian Critical Care 34, 146-154.

Han, L., Wong, F.K.Y., She, D.L., Li, S.Y., Yang, Y.F., Jiang, M.Y., Ruan, Y., Su, Q., Ma, Y.X., Chung, L.Y.F., 2020. Anxiety and Depression of Nurses in a North West Province in China During the Period of Novel Coronavirus Pneumonia Outbreak. Journal of Nursing Scholarship 52, 564-573.

Han, S., Choi, S., Cho, S.H., Lee, J., Yun, J.-Y., 2021. Associations between the working experiences at frontline of COVID-19 pandemic and mental health of Korean public health doctors. BMC Psychiatry 21.

Hasan, S.R., Hamid, Z., Jawaid, M.T., Ali, R.K., 2020. Anxiety among doctors during COVID-19 pandemic in secondary and tertiary care hospitals. Pakistan Journal of Medical Sciences 36, 1360-1365.

Hassamal, S., Dong, F., Hassamal, S., Lee, C., Ogunyemi, D., Neeki, M.M., 2021. The Psychological Impact of COVID-19 on Hospital Staff. Western Journal of Emergency Medicine: Integrating Emergency Care with Population Health 22, 346-352.

HavlİOĞLu, S., DemİR, H.A., 2020. Determining the Anxiety Levels of Emergency Service Employees' Working During The Covid-19 Pandemic. Journal of Harran University Medical Faculty 17, 251-255.

He, L., Wang, J., Zhang, L., Wang, F., Dong, W., Zhao, W., 2021. Risk Factors for Anxiety and Depressive Symptoms in Doctors During the Coronavirus Disease 2019 Pandemic. Frontiers in psychiatry 12, 687440.

He, Q., Ren, J., Wang, G., Zhang, J., Xiang, J., He, D., 2021. Psychological effects of the COVID-19 outbreak on nurses working in tertiary women's and children's hospitals from Sichuan, China: A cross-sectional study. International journal of disaster risk reduction : IJDRR 58, 102188.

Heesakkers, H., Zegers, M., van Mol, M.M.C., van den Boogaard, M., 2021. The impact of the first COVID-19 surge on the mental well-being of ICU nurses: A nationwide survey study. Intensive & Critical Care Nursing 65, N.PAG-N.PAG.

Hennein, R., Lowe, S., 2020. A hybrid inductive-abductive analysis of health workers' experiences and wellbeing during the COVID-19 pandemic in the United States. PLoS ONE 15.

Hennein, R., Mew, E.J., Lowe, S.R., 2021. Socio-ecological predictors of mental health outcomes among healthcare workers during the COVID-19 pandemic in the United States. PloS one 16, e0246602.

Hernandez, J.M., Munyan, K., Kennedy, E., Kennedy, P., Shakoor, K., Wisser, J., 2021. Traumatic stress among frontline American nurses during the COVID-19 pandemic: A survey study. Traumatology 27, 413-418.

Hilmi, M., Boilève, A., Ducousso, A., Michalet, M., Turpin, A., Neuzillet, C., Naoun, N., 2020. Professional and Psychological Impacts of the COVID-19 Pandemic on Oncology Residents: A National Survey. JCO global oncology 6, 1674-1683.

Hoang, D.V., Yamamoto, S., Miki, T., Fukunaga, A., Islam, Z., Konishi, M., Mizoue, T., 2021. Is there an association between ABO blood types and depressive symptoms among Japanese healthcare workers during the COVID-19 pandemic? PloS one 16, e0256441.

Holton, S., Wynter, K., Rothmann, M.J., Skjoeth, M.M., Considine, J., Street, M., Hutchinson, A., Khaw, D., Hutchinson, A.M., Ockerby, C., Crowe, S., Trueman, M., Sweeney, S., Bruce, S., Rasmussen, B., 2021. Australian and Danish nurses' and midwives' wellbeing during COVID-19: a comparison study. Collegian (Royal College of Nursing, Australia).

Holton, S., Wynter, K., Trueman, M., Bruce, S., Sweeney, S., Crowe, S., Dabscheck, A., Eleftheriou, P., Booth, S., Hitch, D., Said, C.M., Haines, K.J., Rasmussen, B., Psychological well-being of Australian hospital clinical staff during the COVID-19 pandemic. Australian Health Review.

Hong, S., Ai, M., Xu, X., Wang, W., Chen, J., Zhang, Q., Wang, L., Kuang, L., 2020. Immediate psychological impact on nurses working at 42 government-designated hospitals during COVID-19 outbreak in China: A cross-sectional study. Nursing outlook.

Hong, Y., Lee, J., Lee, H.J., Kim, K., Cho, I.-K., Ahn, M.H., Shin, Y.-W., Park, J., Chung, S., 2021. Resilience and Work-Related Stress May Affect Depressive Symptoms in Nursing Professionals during the COVID-19 Pandemic Era. Psychiatry investigation 18, 357-363.

Hou, T., Yin, Q., Cai, W., Song, X., Deng, W., Zhang, J., Deng, G., 2021. Posttraumatic stress symptoms among health care workers during the covid-19 epidemic: The roles of negative coping and fatigue. Psychology, Health & Medicine.

Hu, D., Kong, Y., Li, W., Han, Q., Zhang, X., Zhu, L.X., Wan, S.W., Liu, Z., Shen, Q., Yang, J., He, H.G., Zhu, J., 2020. Frontline nurses’ burnout, anxiety, depression, and fear statuses and their associated factors during the COVID-19 outbreak in Wuhan, China: A large-scale cross-sectional study. EClinicalMedicine 24.

Huang, H.L., Chen, R.C., Teo, I., Chaudhry, I., Heng, A.L., Zhuang, K.D., Tan, H.K., Tan, B.S., 2021. A survey of anxiety and burnout in the radiology workforce of a tertiary hospital during the COVID-19 pandemic. Journal of medical imaging and radiation oncology 65, 139-145.

Huang, L., Wang, Y., Liu, J., Ye, P., Chen, X., Xu, H., Qu, H., Ning, G., 2020. Factors Influencing Anxiety of Health Care Workers in the Radiology Department with High Exposure Risk to COVID-19. Medical science monitor : international medical journal of experimental and clinical research 26, e926008.

Ike, I.D., Durand-Hill, M., Elmusharaf, E., Asemota, N., Silva, E., White, E., Awad, W.I., 2021. NHS staff mental health status in the active phase of the COVID-19 era: A staff survey in a large London hospital. General Psychiatry 34.

Ilias, I., Mantziou, V., Vamvakas, E., Kampisiouli, E., Theodorakopoulou, M., Vrettou, C., Douka, E., Vassiliou, A.G., Orfanos, S., Kotanidou, A., Dimopoulou, I., 2021. Post-Traumatic Stress Disorder and Burnout in Healthcare Professionals During the SARS-CoV-2 Pandemic: a Cross-Sectional Study. Journal of critical care medicine (Universitatea de Medicina si Farmacie din Targu-Mures) 7, 14-20.

Imran, N., Masood, H.M.U., Ayub, M., Gondal, K.M., 2020. Psychological impact of COVID-19 pandemic on postgraduate trainees: a cross-sectional survey. Postgraduate medical journal.

Inoue, Y., Yamamoto, S., Fukunaga, A., Hoang, D.V., Miki, T., Islam, Z., Miyo, K., Ishii, M., Ishiwari, H., Konishi, M., Ohmagari, N., Mizoue, T., 2021. Association between engagement in COVID-19-related work and depressive symptoms among hospital workers in a designated COVID-19 hospital in Japan: a cross-sectional study. BMJ open 11, e049996.

Işik, M., Kirli, U., Özdemir, P.G., 2021. The Mental Health of Healthcare Professionals During the COVID-19 Pandemic. Turk psikiyatri dergisi = Turkish journal of psychiatry 32, 225-234.

Jagiasi, B.G., Chanchalani, G., Nasa, P., Tekwani, S., 2021. Impact of COVID-19 Pandemic on the Emotional Well-being of Healthcare Workers: A Multinational Cross-sectional Survey. Indian journal of critical care medicine : peer-reviewed, official publication of Indian Society of Critical Care Medicine 25, 499-506.

Jain, A., Singariya, G., Kamal, M., Kumar, M., Solanki, R.K., 2020. COVID-19 pandemic: Psychological impact on anaesthesiologists. Indian Journal of Anaesthesia 64, 774-783.

Jambunathan, P., Jindal, M., Patra, P., Madhusudan, T., 2020. COVID-warriors: Psychological impact of the severe acute respiratory syndrome coronavirus 2 pandemic on health-care professionals. JOURNAL OF MARINE MEDICAL SOCIETY 22, 57-61.

Jang, O.-J., Chung, Y.-I., Lee, J.-W., Kim, H.-C., Seo, J.S., 2021. Emotional Distress of the COVID-19 Cluster Infection on Health Care Workers Working at a National Hospital in Korea. Journal of Korean medical science 36, e324.

Javadekar, A., Javadekar, S., Chaudhury, S., Saldanha, D., 2021. Depression, anxiety, stress, and sleep disturbances in doctors and general population during COVID-19 pandemic. Industrial psychiatry journal 30, S20-S24.

Jemal, K., Deriba, B.S., Geleta, T.A., 2021. Psychological Distress, Early Behavioral Response, and Perception Toward the COVID-19 Pandemic Among Health Care Workers in North Shoa Zone, Oromiya Region. Frontiers in psychiatry 12, 628898.

Jemal, K., Deriba, B.S., Geleta, T.A., Tesema, M., Awol, M., Mengistu, E., Annous, Y., 2021. Self-Reported Symptoms of Depression, Anxiety, and Stress Among Healthcare Workers in Ethiopia During the COVID-19 Pandemic: A Cross-Sectional Study. Neuropsychiatric disease and treatment 17, 1363-1373.

Jiang, M., Li, S., She, D., Yan, F., Chung, Y.F., Han, L., 2020. The psychological effect of 2019 coronavirus disease outbreak on nurses living in Islamic culture dominant region, China. Archives of Psychiatric Nursing 34, 513-519.

Jo, S.H., Koo, B.H., Seo, W.S., Yun, S.H., Kim, H.G., 2020. The psychological impact of the coronavirus disease pandemic on hospital workers in Daegu, South Korea. Comprehensive Psychiatry 103.

Johnson, A.P., Wohlauer, M.V., Mouawad, N.J., Malgor, R.D., Coogan, S.M., Sheahan, M.G., 3rd, Singh, N., Cuff, R.F., Woo, K., Coleman, D.M., Shalhub, S., 2020. The impact of the COVID-19 pandemic on vascular surgery trainees in the United States. Annals of vascular surgery.

Joshi, R.G., Joshi, P., Shrestha, P., Basnet, P.S., Mahat, P., 2021. Psychological Reactions among Staffs of a Tertiary Eye Hospital in Eastern Nepal during COVID-19 Pandemic. NEPALESE JOURNAL OF OPHTHALMOLOGY 13, 39-50.

Juan, Y., Yuanyuan, C., Qiuxiang, Y., Cong, L., Xiaofeng, L., Yundong, Z., Jing, C., Peifeng, Q., Yan, L., Xiaojiao, X., Yujie, L., 2020. Psychological distress surveillance and related impact analysis of hospital staff during the COVID-19 epidemic in Chongqing, China. Comprehensive Psychiatry 103.

Kabunga, A., Okalo, P., 2021. Frontline Nurses' Post-Traumatic Stress Disorder and Associated Predictive Factors During the Second Wave of COVID-19 in Central, Uganda. Neuropsychiatric disease and treatment 17, 3627-3633.

Kader, N., Elhusein, B., Chandrappa, N.S.K., Nashwan, A.J., Chandra, P., Khan, A.W., Alabdulla, M., 2021. Perceived stress and post-traumatic stress disorder symptoms among intensive care unit staff caring for severely ill coronavirus disease 2019 patients during the pandemic: a national study. Annals of General Psychiatry 20, 1-8.

Kafle, B., Bagale, Y., Kafle, S., Parajuli, A., Pandey, S., 2021. Depression, Anxiety and Stress among Healthcare Workers during COVID-19 Pandemic in a Tertiary Care Centre of Nepal: A Descriptive Cross-sectional Study. JNMA; journal of the Nepal Medical Association 59, 239-242.

Kalyanaraman, M., Sankar, A., Timpo, E., McQueen, D., Morparia, K., Bergel, M., Rosenblatt, J., 2021. Posttraumatic Stress Namong Pediatric Critical Care Physicians in the United States in Association with Coronavirus Disease 2019 Patient Care Experiences. Journal of intensive care medicine, 8850666211059385.

Kandemir, D., Temiz, Z., Ozhanli, Y., Erdogan, H., Kanbay, Y., 2021. Analysis of mental health symptoms and insomnia levels of intensive care nurses during the covid‐19 pandemic with a structural equation model. Journal of Clinical Nursing.

Kannampallil, T.G., Goss, C.W., Evanoff, B.A., Strickland, J.R., McAlister, R.P., Duncan, J., 2020. Exposure to COVID-19 patients increases physician trainee stress and burnout. PLoS ONE 15.

Katsuta, N., Ito, K., Fukuda, H., Seyama, K., Hori, S., Shida, Y., Nagura, R., Nojiri, S., Sato, H., 2021. Elevated depressive symptoms among newer and younger healthcare workers in Japan during the COVID-19 pandemic. Neuropsychopharmacology reports 41, 544-547.

Khan, H., Srivastava, R., Tripathi, N., Uraiya, D., Singh, A., Verma, R., 2021. Level of anxiety and depression among health-care professionals amidst of coronavirus disease: A web-based survey from India. Journal of education and health promotion 10, 408.

Khan, T.M., Tahir, H., Salman, M., Mustafa, Z.U., Raza, M.H., Asif, N., Shehzadi, N., Hussain, K., Al-Worafi, Y.M., Biag, M.R., 2021. General Anxiety Predictors among Frontline Warriors of COVID: Cross-Sectional Study among Nursing Staff in Punjab, Pakistan. ARCHIVES OF PHARMACY PRACTICE 12, 40-44.

Khanal, P., Devkota, N., Dahal, M., Paudel, K., Joshi, D., 2020. Mental health impacts among health workers during COVID-19 in a low resource setting: A cross-sectional survey from Nepal. Globalization and Health 16.

Khanam, A., Dar, S.A., Wani, Z.A., Shah, N.N., Haq, I., Kousar, S., 2020. Healthcare Providers on the Frontline: A Quantitative Investigation of the Stress and Recent Onset Psychological Impact of Delivering Health Care Services During COVID-19 in Kashmir. Indian journal of psychological medicine 42, 359-367.

Khatun, M.F., Parvin, M.F., Rashid, M.M.-U., Alam, M.S., Kamrunnahar, M., Talukder, A., Rahman Razu, S., Ward, P.R., Ali, M., 2021. Mental Health of Physicians During COVID-19 Outbreak in Bangladesh: A Web-Based Cross-Sectional Survey. Frontiers in public health 9, 592058.

Kibret, S., Teshome, D., Fenta, E., Hunie, M., Tamire, T., 2020. Prevalence of anxiety towards COVID-19 and its associated factors among healthcare workers in a Hospital of Ethiopia. PLoS ONE 15.

Kim, H.J., Lee, G.H., 2021. A comparative study of the psychological impacts of tasks related and unrelated to coronavirus disease 2019 (COVID-19) on nurses. Yeungnam University journal of medicine.

Kim, M.-Y., Yang, Y.-Y., 2021. Mental Health Status and Its Influencing Factors: The Case of Nurses Working in COVID-19 Hospitals in South Korea. International journal of environmental research and public health 18.

Koksal, E., Dost, B., Terzi, O., Ustun, Y.B., Ozdin, S., Bilgin, S., 2020. Evaluation of Depression and Anxiety Levels and Related Factors Among Operating Theater Workers During the Novel Coronavirus (COVID-19) Pandemic. Journal of Perianesthesia Nursing 35, 472-477.

Korkmaz, S., Kazgan, A., Çekiç, S., Tartar, A.S., Balcı, H.N., Atmaca, M., 2020. The anxiety levels, quality of sleep and life and problem-solving skills in healthcare workers employed in COVID-19 services. Journal of Clinical Neuroscience 80, 131-136.

Korkut, S., 2021. Research on the frequency of post-traumatic stress disorder in healthcare workers during the COVID-19 pandemic. Irish journal of medical science.

Kovner, C., Raveis, V.H., Van Devanter, N., Yu, G., Glassman, K., Ridge, L.J., 2021. The psychosocial impact on frontline nurses of caring for patients with COVID-19 during the first wave of the pandemic in New York City. Nursing Outlook 69, 744-754.

Kumar, D., Saghir, T., Ali, G., Yasin, U., Furnaz, S., Karim, M., Hussain, M., Kumari, R., Bai, R., Kumar, H., 2021. Psychosocial Impact of COVID-19 on Healthcare Workers at a Tertiary Care Cardiac Center of Karachi Pakistan. Journal of occupational and environmental medicine 63, e59-e62.

Lai, J., Ma, S., Wang, Y., Cai, Z., Hu, J., Wei, N., Wu, J., Du, H., Chen, T., Li, R., Tan, H., Kang, L., Yao, L., Huang, M., Wang, H., Wang, G., Liu, Z., Hu, S., 2020. Factors associated with mental health outcomes among health care workers exposed to coronavirus disease 2019. JAMA Network Open 3.

Lamb, D., Gnanapragasam, S., Greenberg, N., Bhundia, R., Carr, E., Hotopf, M., Razavi, R., Raine, R., Cross, S., Dewar, A., Docherty, M., Dorrington, S., Hatch, S., Wilson-Jones, C., Leightley, D., Madan, I., Marlow, S., McMullen, I., Rafferty, A.-M., Parsons, M., Polling, C., Serfioti, D., Gaunt, H., Aitken, P., Morris-Bone, J., Simela, C., French, V., Harris, R., Stevelink, S.A.M., Wessely, S., 2021. Psychosocial impact of the COVID-19 pandemic on 4378 UK healthcare workers and ancillary staff: initial baseline data from a cohort study collected during the first wave of the pandemic. Occupational and environmental medicine 78, 801-808.

Lamiani, G., Borghi, L., Poli, S., Razzini, K., Colosio, C., Vegni, E., 2021. Hospital Employees' Well-Being Six Months after the COVID-19 Outbreak: Results from a Psychological Screening Program in Italy. International journal of environmental research and public health 18.

Lan, V.T.H., Dzung, L.T., Quyen, B.T.T., Nha, P.B., Linh, N.T., Hoang, L.T., Nghia, N.Q., Hung, D.D., 2021. Impact of Central Quarantine Inside a Lockdown Hospital Due to COVID-19 Pandemic on Psychological Disorders among Health Care Staffs in Central Hospitals of Hanoi, Vietnam, 2020. Health Services Insights, 1-7.

Lang, Q., Liu, X., He, Y., Lv, Q., Xu, S., 2020. Association between working hours and anxiety/depression of medical staff during large-scale epidemic outbreak of COVID-19: A cross-sectional study. Psychiatry Investigation 17, 1167-1174.

Lasalvia, A., Bodini, L., Amaddeo, F., Porru, S., Carta, A., Poli, R., Bonetto, C., 2021. The Sustained Psychological Impact of the COVID-19 Pandemic on Health Care Workers One Year after the Outbreak-A Repeated Cross-Sectional Survey in a Tertiary Hospital of North-East Italy. International journal of environmental research and public health 18.

Lasalvia, A., Bonetto, C., Porru, S., Carta, A., Tardivo, S., Bovo, C., Ruggeri, M., Amaddeo, F., 2020. Psychological impact of COVID-19 pandemic on healthcare workers in a highly burdened area of north-east Italy. Epidemiology and Psychiatric Sciences 30.

Laurent, A., Fournier, A., Lheureux, F., Poujol, A.-L., Deltour, V., Ecarnot, F., Meunier-Beillard, N., Loiseau, M., Binquet, C., Quenot, J.-P., 2022. Risk and protective factors for the possible development of post-traumatic stress disorder among intensive care professionals in France during the first peak of the COVID-19 epidemic. EUROPEAN JOURNAL OF PSYCHOTRAUMATOLOGY 13.

Lee, J., Lee, H.J., Hong, Y., Shin, Y.-W., Chung, S., Park, J., 2021. Risk Perception, Unhealthy Behavior, and Anxiety Due to Viral Epidemic Among Healthcare Workers: The Relationships With Depressive and Insomnia Symptoms During COVID-19. Frontiers in psychiatry 12, 615387.

Lei, L., Tang, J., Su, D., Deng, D., Huang, X., 2021. Relevant Factors and Intervention Measures of Psychological Stress-Induced Hyperthermia among Medical Staff in Temporary COVID-19 Negative Pressure Wards. IRANIAN JOURNAL OF PUBLIC HEALTH 50, 2526-2535.

Li, J., Xu, J., Zhou, H., You, H., Wang, X., Li, Y., Liang, Y., Li, S., Ma, L., Zeng, J., Cai, H., Xie, J., Pan, C., Hao, C., Gilmour, S., Lau, J.T.-f., Hao, Y., Xu, D.R., Gu, J., 2021. Working conditions and health status of 6,317 front line public health workers across five provinces in China during the COVID-19 epidemic: a cross-sectional study. BMC Public Health 21, 1-14.

Li, J., Zhang, Y., Li, L., Yi, W., Hao, Y., Bi, Y., 2021. Predictive Analysis of Factors Influencing Depression Status of Nurses in the COVID-19 Pandemic Intensive Care Unit. Frontiers in psychiatry 12, 596428.

Li, L., Wang, X., Tan, J., Li, J., Yuan, Y., 2021. Influence of sleep difficulty on post-traumatic stress symptoms among frontline medical staff during covid-19 pandemic in china. Psychology, Health & Medicine.

Li, Q., Chen, J.L., Xu, G., Zhao, J., Yu, X.Q., Wang, S.Y., Liu, L., Liu, F., 2020. The Psychological Health Status of Healthcare Workers During the COVID-19 Outbreak: A Cross-Sectional Survey Study in Guangdong, China. Frontiers in Public Health 8.

Li, T.-M., Pien, L.-C., Kao, C.-C., Kubo, T., Cheng, W.-J., 2022. Effects of work conditions and organisational strategies on nurses' mental health during the COVID-19 pandemic. JOURNAL OF NURSING MANAGEMENT 30, 71-78.

Li, X., Li, S., Xiang, M., Fang, Y., Qian, K., Xu, J., Li, J., Zhang, Z., Wang, B., 2020. The prevalence and risk factors of PTSD symptoms among medical assistance workers during the COVID-19 pandemic. J Psychosom Res 139, 110270-110270.

Li, X., Yu, H., Yang, W., Mo, Q., Yang, Z., Wen, S., Zhao, F., Zhao, W., Tang, Y., Ma, L., Zeng, R., Zou, X., Lin, H., 2021. Depression and Anxiety Among Quarantined People, Community Workers, Medical Staff, and General Population in the Early Stage of COVID-19 Epidemic. Frontiers in psychology 12, 638985.

Li, X.C., Zhou, Y., Xu, X.Y., Factors associated with the psychological well-being among front-line nurses exposed to COVID-2019 in China: A predictive study. Journal of Nursing Management.

Liang, Y., Wu, K., Zhou, Y., Huang, X., Zhou, Y., Liu, Z., 2020. Mental Health in Frontline Medical Workers during the 2019 Novel Coronavirus Disease Epidemic in China: A Comparison with the General Population. International Journal of Environmental Research and Public Health 17, 6550.

Liu, C.Y., Yang, Y.Z., Zhang, X.M., Xu, X., Dou, Q.L., Zhang, W.W., Cheng, A.S.K., 2020. The prevalence and influencing factors in anxiety in medical workers fighting COVID-19 in China: A cross-sectional survey. Epidemiology and Infection 148.

Liu, S., Han, W., Shen, C., Zhu, C., Wang, Q., Liang, X., He, X., Xie, Q., Wei, J., Wu, M., Zhao, X., Liu, H., Liu, D., Guo, X., Nie, S., Cao, L., Lu, L., Fang, Y., Lu, Z., Wu, Y., Zhao, M., Han, J., Zhang, X., Chang, J., Xu, S., Ma, W., Si, J., Qi, S., Peng, P., Chai, Y., Cao, Y., Jiang, Y., Yin, W., Wang, Y., Zhan, H., Huang, Y., Deng, Y., Song, J., Yang, L., Wu, J., Ding, B., Zheng, D., Qian, C., Huang, R., Lin, J., Xu, Z., Zhang, G., Hu, Y., Dou, Q., Zhang, X., Tian, Y., Yao, D., Walline, J.H., Zhu, H., Xu, J., Li, Y., Yu, X., 2021. Depressive State in the Emergency Department During COVID-19: A National Cross-Sectional Survey in China. Frontiers in psychiatry 12, 566990.

Liu, S., Xu, R., Liu, L., 2021. Influencing Factors of Acute Stress Disorder Among Frontline Nurses in Wuhan, China. Journal of Psychosocial Nursing & Mental Health Services 59, 38-47.

Liu, T., Zheng, Z., Sha, X., Liu, H., Zheng, W., Su, H., Xu, G., Su, K.P., So, K.F., Lin, K., 2020. Psychological impact in non-infectious disease specialists who had direct contact with patients with COVID-19. BJPsych Open 7.

Liu, Y., Chen, H., Zhang, N., Wang, X., Fan, Q., Zhang, Y., Huang, L., Hu, B., Li, M., 2020. Anxiety and depression symptoms of medical staff under COVID-19 epidemic in China. Journal of affective disorders 278, 144-148.

Londoño-Ramírez, A.C., García-Pla, S., Bernabeu-Juan, P., Pérez-Martínez, E., Rodríguez-Marín, J., van-der Hofstadt-Román, C.J., 2021. Impact of COVID-19 on the Anxiety Perceived by Healthcare Professionals: Differences between Primary Care and Hospital Care. International journal of environmental research and public health 18.

Lu, M.-Y., Ahorsu, D.K., Kukreti, S., Strong, C., Lin, Y.-H., Kuo, Y.-J., Chen, Y.-P., Lin, C.-Y., Chen, P.-L., Ko, N.-Y., Ko, W.-C., 2021. The Prevalence of Post-traumatic Stress Disorder Symptoms, Sleep Problems, and Psychological Distress Among COVID-19 Frontline Healthcare Workers in Taiwan. Frontiers in psychiatry 12, 705657.

Lu, P., Li, X., Lu, L., Zhang, Y., 2020. The psychological states of people after Wuhan eased the lockdown. PLoS ONE 15.

Luo, D., Liu, Q., Chen, Q., Huang, R., Chen, P., Yang, B.X., Liu, Z., 2021. Mental Health Status of the General Public, Frontline, and Non-frontline Healthcare Providers in the Early Stage of COVID-19. Frontiers in psychiatry 12, 553021.

Magnavita, N., Soave, P.M., Antonelli, M., 2021. A One-Year Prospective Study of Work-Related Mental Health in the Intensivists of a COVID-19 Hub Hospital. International journal of environmental research and public health 18.

Magnavita, N., Soave, P.M., Antonelli, M., 2021. Prolonged Stress Causes Depression in Frontline Workers Facing the COVID-19 Pandemic-A Repeated Cross-Sectional Study in a COVID-19 Hub-Hospital in Central Italy. International journal of environmental research and public health 18.

Magnavita, N., Soave, P.M., Ricciardi, W., Antonelli, M., 2020. Occupational Stress and Mental Health among Anesthetists during the COVID-19 Pandemic. International journal of environmental research and public health 17.

Magnavita, N., Tripepi, G., Di Prinzio, R.R., 2020. Symptoms in health care workers during the covid-19 epidemic. A cross-sectional survey. International Journal of Environmental Research and Public Health 17, 1-15.

Maqbali, M.A., Khadhuri, J.A., 2021. Psychological impact of the coronavirus 2019 (COVID-19) pandemic on nurses. Japan Journal of Nursing Science 18, 1-10.

Marco, C.A., Larkin, G.L., Feeser, V.R., Monti, J.E., Vearrier, L., 2020. Post-traumatic stress and stress disorders during the COVID-19 pandemic: Survey of emergency physicians. Journal of the American College of Emergency Physicians open 1, 1594-1601.

Marcomini, I., Agus, C., Milani, L., Sfogliarini, R., Bona, A., Castagna, M., 2021. COVID-19 and post-traumatic stress disorder among nurses: a descriptive cross-sectional study in a COVID hospital. La Medicina del lavoro 112, 241-249.

Marthoenis, Maskur, Fathiariani, L., Nassimbwa, J., 2021. Investigating the burden of mental distress among nurses at a provincial COVID-19 referral hospital in Indonesia: a cross-sectional study. BMC Nursing 20, 1-8.

Martínez-Caballero, C.M., Cárdaba-García, R.M., Varas-Manovel, R., García-Sanz, L.M., Martínez-Piedra, J., Fernández-Carbajo, J.J., Pérez-Pérez, L., Madrigal-Fernández, M.A., Barba-Pérez, M.Á., Olea, E., Durantez-Fernández, C., Herrero-Frutos, M.T., 2021. Analyzing the Impact of COVID-19 Trauma on Developing Post-Traumatic Stress Disorder among Emergency Medical Workers in Spain. International journal of environmental research and public health 18.

Matsumoto, Y., Fujino, J., Shiwaku, H., Miyajima, M., Doi, S., Hirai, N., Jitoku, D., Takagi, S., Tamura, T., Maruo, T., Shidei, Y., Kobayashi, N., Ichihashi, M., Noguchi, S., Oohashi, K., Takeuchi, T., Sugihara, G., Okada, T., Fujiwara, T., Takahashi, H., 2021. Factors affecting mental illness and social stress in hospital workers treating COVID-19: Paradoxical distress during pandemic era. Journal of Psychiatric Research 137, 298-302.

Mattila, E., Peltokoski, J., Neva, M.H., Kaunonen, M., Helminen, M., Parkkila, A.-K., 2021. COVID-19: anxiety among hospital staff and associated factors. Annals of medicine 53, 237-246.

Mehta, S., Yarnell, C., Shah, S., Dodek, P., Parsons-Leigh, J., Maunder, R., Kayitesi, J., Eta-Ndu, C., Priestap, F., LeBlanc, D., Chen, J., Honarmand, K., 2021. The impact of the COVID-19 pandemic on intensive care unit workers: a nationwide survey. Canadian journal of anaesthesia = Journal canadien d'anesthesie.

Mei, S., Liang, L., Ren, H., Hu, Y., Qin, Z., Cao, R., Li, C., Fei, J., Yuan, T., Meng, C., Guo, X., Lv, J., Hu, Y., 2021. Association Between Perceived Stress and Post-Traumatic Stress Disorder Among Medical Staff During the COVID-19 Epidemic in Wuhan City. Frontiers in public health 9, 666460.

Mendonça, V.S., Steil, A., Góis, A.F.T., 2021. Mental health and the COVID-19 pandemic: a study of medical residency training over the years. Clinics (Sao Paulo, Brazil) 76, e2907.

Mendonça, V.S., Steil, A., Teixeira de Gois, A.F., 2021. COVID-19 pandemic in São Paulo: a quantitative study on clinical practice and mental health among medical residency specialties. Sao Paulo medical journal = Revista paulista de medicina 139, 489-495.

Mensinger, J.L., Brom, H., Havens, D.S., Costello, A., D'Annunzio, C., Durning, J.D., Bradley, P.K., Copel, L., Maldonado, L., Smeltzer, S., Yost, J., Kaufmann, P., 2022. Psychological responses of hospital-based nurses working during the COVID-19 pandemic in the United States: A cross-sectional study. APPLIED NURSING RESEARCH 63.

Meo, S.A., Alkhalifah, J.M., Alshammari, N.F., Alnufaie, W.S., 2021. Comparison of Generalized Anxiety and Sleep Disturbance among Frontline and Second-Line Healthcare Workers during the COVID-19 Pandemic. International journal of environmental research and public health 18.

Moderato, L., Lazzeroni, D., Oppo, A., Dell'Orco, F., Moderato, P., Presti, G., 2021. Acute Stress Response Profiles in Health Workers Facing SARS-CoV-2. Frontiers in psychology 12, 660156.

Molina-Mula, J., González-Trujillo, A., Perelló-Campaner, C., Tortosa-Espínola, S., Tera-Donoso, J., la Rosa, L.O.D., Romero-Franco, N., 2021. THE EMOTIONAL IMPACT OF COVID-19 ON SPANISH NURSES AND POTENTIAL STRATEGIES TO REDUCE IT. Collegian (Royal College of Nursing, Australia).

Moon, D.-J., Han, M.A., Park, J., Ryu, S.Y., 2021. Post-traumatic stress and related factors among hospital nurses during the covid-19 outbreak in korea. Psychiatric Quarterly.

Moore, C.J.S., Blencowe, N.S., Hollén, L., van Hamel, C., 2021. Interim Foundation Year One (FiY1) and preparedness for foundation year 1: A national survey of UK foundation doctors. Medical teacher, 1-7.

Morawa, E., Schug, C., Geiser, F., Beschoner, P., Jerg-Bretzke, L., Albus, C., Weidner, K., Hiebel, N., Borho, A., Erim, Y., 2021. Psychosocial burden and working conditions during the COVID-19 pandemic in Germany: The VOICE survey among 3678 health care workers in hospitals. Journal of Psychosomatic Research 144, N.PAG-N.PAG.

Mosheva, M., Gross, R., Hertz-Palmor, N., Hasson-Ohayon, I., Kaplan, R., Cleper, R., Kreiss, Y., Gothelf, D., Pessach, I.M., 2021. The association between witnessing patient death and mental health outcomes in frontline COVID-19 healthcare workers. Depression and anxiety 38, 468-479.

Mosolova, E., Sosin, D., Mosolov, S., 2021. Stress, anxiety, depression and burnout in frontline healthcare workers during two peaks of COVID-19 pandemic in Russia. Psychiatry research 306, 114226.

Motahedi, S., Aghdam, N.F., Khajeh, M., Baha, R., Aliyari, R., Bagheri, H., Mardani, A., 2021. Anxiety and depression among healthcare workers during COVID-19 pandemic: A cross-sectional study. Heliyon 7, e08570.

Msheik El Khoury, F., Talih, F., Khatib, M.F.E., Abi Younes, N., Siddik, M., Siddik-Sayyid, S., 2021. Factors Associated with Mental Health Outcomes: Results from a Tertiary Referral Hospital in Lebanon during the COVID-19 Pandemic. The Libyan journal of medicine 16, 1901438.

Mulatu, H.A., Tesfaye, M., Woldeyes, E., Bayisa, T., Fisseha, H., Kassu, R.A., 2021. The prevalence of common mental disorders among healthcare professionals during the COVID-19 pandemic at a tertiary Hospital in Addis Ababa, Ethiopia. Journal of affective disorders reports 6, 100246.

Naheed, A., Ahmed, A., Choudhary, Z.I., Fatima, S., Naseem, S., Ghias, M., 2021. COVID-19 Pandemic: Psychological Stress and Coping Strategies among Doctors. PAKISTAN JOURNAL OF MEDICAL & HEALTH SCIENCES 15, 2504-2507.

Nakhostin-Ansari, A., Sherafati, A., Aghajani, F., Khonji, M., Aghajani, R., Shahmansouri, N., 2020. Depression and Anxiety among Iranian Medical Students during COVID-19 Pandemic. Iranian Journal of Psychiatry.

Naldi, A., Vallelonga, F., Di Liberto, A., Cavallo, R., Agnesone, M., Gonella, M., Sauta, M.D., Lochner, P., Tondo, G., Bragazzi, N.L., Botto, R., Leombruni, P., 2020. COVID-19 pandemic-related anxiety, distress and burnout: Prevalence and associated factors in healthcare workers of North-West Italy. BJPsych Open 7.

Nayak, B.S., Sahu, P.K., Ramsaroop, K., Maharaj, S., Mootoo, W., Khan, S., Extravour, R.M., 2021. Prevalence and factors associated with depression, anxiety and stress among healthcare workers of Trinidad and Tobago during COVID-19 pandemic: a cross-sectional study. BMJ open 11, e044397.

Ng, K.Y.Y., Zhou, S., Tan, S.H., Ishak, N.D.B., Goh, Z.Z.S., Chua, Z.Y., Chia, J.M.X., Chew, E.L., Shwe, T., Mok, J.K.Y., Si Leong, S., Si Ying Lo, J., Li Ting Ang, Z., Leow, J.L., Lam, C.W.J., Kwek, J.W., Dent, R., Tuan, J., Lim, S.T., Hwang, W.Y.K., Griva, K., Ngeow, J., 2020. Understanding the psychological impact of COVID-19 pandemic on patients with cancer, their caregivers, and health care workers in Singapore. JCO Global Oncology, 1494-1509.

Nguyen, T.T., Le, X.T.T., Nguyen, N.T.T., Nguyen, Q.N., Le, H.T., Pham, Q.T., Ta, N.K.T., Nguyen, Q.T., Nguyen, A.N., Hoang, M.T., Pham, H.Q., Vu, L.G., Luong, A.M., Koh, D., Nguyen, T.H., Tran, B.X., Latkin, C.A., Ho, C.S.H., Ho, R.C.M., 2021. Psychosocial Impacts of COVID-19 on Healthcare Workers During the Nationwide Partial Lockdown in Vietnam in April 2020. Frontiers in psychiatry 12, 562337.

Ni, M.Y., Yang, L., Leung, C.M.C., Li, N., Yao, X.I., Wang, Y.S., Leung, G.M., Cowling, B.J., Liao, Q.Y., 2020. Mental Health, Risk Factors, and Social Media Use During the COVID-19 Epidemic and Cordon Sanitaire Among the Community and Health Professionals in Wuhan, China: Cross-Sectional Survey. Jmir Mental Health 7.

Noaimi, H.M.A., Noaimi, M.M.A., Fayez, F.M.A., Mushkhes, H.Q.A., Al Ani, W., 2021. The Impact of COVID-19 Pandemic on Mental Health of Health Care Workers of Bahrain Defence Force Royal Medical Services. BAHRAIN MEDICAL BULLETIN 43, 580-587.

Norhayati, M.N., Che Yusof, R., Azman, M.Y., 2021. Depressive symptoms among frontline and non-frontline healthcare providers in response to the COVID-19 pandemic in Kelantan, Malaysia: A cross sectional study. PloS one 16, e0256932.

Ofori, A.A., Osarfo, J., Agbeno, E.K., Manu, D.O., Amoah, E., 2021. Psychological impact of COVID-19 on health workers in Ghana: A multicentre, cross-sectional study. SAGE open medicine 9, 20503121211000919.

Okajima, I., Chung, S., Suh, S., 2021. Validation of the Japanese version of Stress and Anxiety to Viral Epidemics-9 (SAVE-9) and relationship among stress, insomnia, anxiety, and depression in healthcare workers exposed to coronavirus disease 2019. Sleep Medicine 84, 397-402.

Onchonga, D., Ngetich, E., Makunda, W., Wainaina, P., Wangeshi, D., Viktoria, P., 2021. Anxiety and depression due to 2019 SARS-CoV-2 among frontier healthcare workers in Kenya. Heliyon 7, e06351.

Önen Sertöz, Ö., Kuman Tunçel, Ö., Sertöz, N., Hepdurgun, C., İşman Haznedaroglu, D., Bor, C., 2021. Burnout in healthcare professionals during the Covid-19 pandemic in a tertiary care university hospital: Evaluation of the need for psychological support. Türk Psikiyatri Dergisi 32, 75-86.

Ozdemir, S., Akca, H.S., Algin, A., ozkan, A., Eroglu, S.E., Kurtulus, S.A., Sevimli, H., Donmez, Z., 2021. The effect of the Coronavirus 2019 pandemic on the mental health of healthcare workers. ANNALS OF CLINICAL AND ANALYTICAL MEDICINE 12, 1348-1352.

Pachi, A., Sikaras, C., Ilias, I., Panagiotou, A., Zyga, S., Tsironi, M., Baras, S., Tsitrouli, L.A., Tselebis, A., 2022. Burnout, Depression and Sense of Coherence in Nurses during the Pandemic Crisis. HEALTHCARE 10.

Pan, L., Xu, Q., Kuang, X., Zhang, X., Fang, F., Gui, L., Li, M., Tefsen, B., Zha, L., Liu, H., 2021. Prevalence and factors associated with post-traumatic stress disorder in healthcare workers exposed to COVID-19 in Wuhan, China: A cross-sectional survey. BMC Psychiatry 21.

Pan, X., Xiao, Y., Ren, D., Xu, Z.M., Zhang, Q., Yang, L.Y., Liu, F., Hao, Y.S., Zhao, F., Bai, Y.H., 2020. Prevalence of mental health problems and associated risk factors among military healthcare workers in specialized COVID ‐19 hospitals in Wuhan, China: A cross‐sectional survey. Asia-Pacific Psychiatry.

Pang, Y., Fang, H., Li, L., Chen, M., Chen, Y., Chen, M., 2021. Predictive factors of anxiety and depression among nurses fighting coronavirus disease 2019 in China. International Journal of Mental Health Nursing 30, 524-532.

Pappa, S., Athanasiou, N., Sakkas, N., Patrinos, S., Sakka, E., Barmparessou, Z., Tsikrika, S., Adraktas, A., Pataka, A., Migdalis, I., Gida, S., Katsaounou, P., 2021. From Recession to Depression? Prevalence and Correlates of Depression, Anxiety, Traumatic Stress and Burnout in Healthcare Workers during the COVID-19 Pandemic in Greece: A Multi-Center, Cross-Sectional Study. International journal of environmental research and public health 18.

Park, C., Hwang, J.M., Jo, S., Bae, S.J., Sakong, J., 2020. COVID-19 Outbreak and Its Association with Healthcare Workers' Emotional Stress: a Cross-Sectional Study. Journal of Korean Medical Science 35.

Park, S., Lee, Y., Kim, T., Jung, S.J., 2021. Anxiety and COVID-19 Related Stressors Among Healthcare Workers Who Performed Shift Work at Four COVID-19 Dedicated Hospitals in Korea. Journal of Occupational & Environmental Medicine 63, 875-880.

Park, Y.S., Park, K.-H., Lee, J., 2021. Validation of the Korean Version of Impact of Event Scale-Revised (IES-R) in Korean Nurses during the COVID-19 Pandemic. International journal of environmental research and public health 18.

Parthasarathy, R., Ts, J., K, T., Murthy, P., 2021. Mental health issues among health care workers during the COVID-19 pandemic - A study from India. Asian journal of psychiatry 58, 102626.

Pascoe, A., Paul, E., Johnson, D., Putland, M., Willis, K., Smallwood, N., 2021. Differences in Coping Strategies and Help-Seeking Behaviours among Australian Junior and Senior Doctors during the COVID-19 Pandemic. International journal of environmental research and public health 18.

Patel, A.V., Kandre, D.D., Mehta, P., Prajapati, A., Patel, B., Prajapati, S., 2020. Multi-centric study of psychological disturbances among health care workers in tertiary care centers of western India during the COVID-19 pandemic. NEUROPSYCHIATRIA I NEUROPSYCHOLOGIA 15, 89-100.

Patel, V.K., Pandey, S., Jani, A.M., Tiwari, D.S., Patel, F.B., Thakrar, R.K.A., 2021. COVID-19 Outbreak: Impact on Psychological Well-Being of the Health-Care Workers of a Designated COVID-19 Hospital. JOURNAL OF MENTAL HEALTH AND HUMAN BEHAVIOUR 26, 20-27.

Patel, V.K., Pandey, S., Patel, F.B., Jani, A.M., Tiwari, D.S., Thakrar, R.K.A., 2021. Study of correlates of depression among health care workers during COVID-19 epidemic. ARCHIVES OF PSYCHIATRY AND PSYCHOTHERAPY 23, 29-35.

Peñacoba, C., Velasco, L., Catalá, P., Gil‐Almagro, F., García‐Hedrera, F.J., Carmona‐Monge, F.J., 2021. Resilience and anxiety among intensive care unit professionals during the COVID‐19 pandemic. Nursing in Critical Care 26, 501-509.

Peng, X., Meng, X., Li, L., Hu, C., Liu, W., Liu, Z., Ma, X., Xu, D., Xing, Z., Zhu, Z., Liu, B., Zhang, L., Peng, M., 2021. Depressive and Anxiety Symptoms of Healthcare Workers in Intensive Care Unit Under the COVID-19 Epidemic: An Online Cross-Sectional Study in China. Frontiers in public health 9, 603273.

Perera, B., Wickramarachchi, B., Samanmalie, C., Hettiarachchi, M., 2021. Psychological experiences of healthcare professionals in Sri Lanka during COVID-19. BMC psychology 9, 49.

Petrișor, C., Breazu, C., Doroftei, M., Mărieș, I., Popescu, C., 2021. Association of Moral Distress with Anxiety, Depression, and an Intention to Leave among Nurses Working in Intensive Care Units during the COVID-19 Pandemic. Healthcare (Basel, Switzerland) 9.

Pouralizadeh, M., Bostani, Z., Maroufizadeh, S., Ghanbari, A., Khoshbakht, M., Alavi, S.A., Ashrafi, S., 2020. Anxiety and depression and the related factors in nurses of Guilan University of Medical Sciences hospitals during COVID-19: A web-based cross-sectional study. International Journal of Africa Nursing Sciences 13, 100233.

Prasad, A., Civantos, A.M., Byrnes, Y., Chorath, K., Poonia, S., Chang, C., Graboyes, E.M., Bur, A.M., Thakkar, P., Deng, J., Seth, R., Trosman, S., Wong, A., Laitman, B.M., Shah, J., Stubbs, V., Long, Q., Choby, G., Rassekh, C.H., Thaler, E.R., Rajasekaran, K., 2020. Snapshot Impact of COVID-19 on Mental Wellness in Nonphysician Otolaryngology Health Care Workers: A National Study. OTO open 4, 2473974X20948835.

Proserpio, P., Zambrelli, E., Lanza, A., Dominese, A., Di Giacomo, R., Quintas, R., Tramacere, I., Rubino, A., Turner, K., Colosio, C., Cattaneo, F., Canevini, M.P., D'Agostino, A., Agostoni, E.C., Didato, G., Sleep disorders and mental health in hospital workers during the COVID-19 pandemic: a cross-sectional multicenter study in Northern Italy. NEUROLOGICAL SCIENCES.

Qasem Surrati, A.M., Asad Mansuri, F.M., Ayadh Alihabi, A.A., 2020. Psychological impact of the COVID-19 pandemic on health care workers. Journal of Taibah University Medical Sciences.

Quang, L.N., Kien, N.T., Anh, P.N., Anh, D.T.V., Nghi, T.D.B., Lan, P.P., Anh, N.T., Son, N.V., Lieu, N.T.T., 2021. The Level of Expression of Anxiety and Depression in Clinical Health Care Workers during the COVID-19 Outbreak in 2 Hospitals in Hanoi, Vietnam. Health Services Insights, 1-7.

Qutishat, M., Abu Sharour, L., Al-Dameery, K., Al-Harthy, I., Al-Sabei, S., 2021. COVID-19-Related Posttraumatic Stress Disorder Among Jordanian Nurses During the Pandemic. Disaster medicine and public health preparedness, 1-8.

Rahman, A., Deeba, F., Akhter, S., Bashar, F., Nomani, D., Koot, J., Koly, K.N., Bin Salah, F., Haverlag, K., Anwar, I., 2021. Mental health condition of physicians working frontline with COVID-19 patients in Bangladesh. BMC Psychiatry 21.

Ren, C., Zhou, D., Fan, Y., Li, B., Zhang, W., Shen, Y., Yu, S., Jiang, L., Yu, F., Duan, Y., Peng, D., Cheng, X., Wu, L., Wu, C., Ye, D., 2022. Prevalence and influencing factors of anxiety and depression symptoms among surgical nurses during COVID-19 pandemic: A large-scale cross-sectional study. NURSING OPEN 9, 752-764.

Ren, H., Luo, X., Wang, Y., Guo, X., Hou, H., Zhang, Y., Yang, P., Zhu, F., Hu, C., Wang, R., Sun, Y., Du, Y., Yin, Q., Xu, G., Zuo, H., Hu, Q., Wang, Y., 2021. Psychological responses among nurses caring for patients with COVID-19: a comparative study in China. Translational psychiatry 11, 273.

Roberts, T., Daniels, J., Hulme, W., Hirst, R., Horner, D., Lyttle, M.D., Samuel, K., Graham, B., Reynard, C., Barrett, M., Foley, J., Cronin, J., Umana, E., Vinagre, J., Carlton, E., 2021. Psychological distress and trauma in doctors providing frontline care during the COVID-19 pandemic in the United Kingdom and Ireland: a prospective longitudinal survey cohort study. BMJ open 11, e049680.

Saddik, B., Elbarazi, I., Temsah, M.-H., Saheb Sharif-Askari, F., Kheder, W., Hussein, A., Najim, H., Bendardaf, R., Hamid, Q., Halwani, R., 2021. Psychological Distress and Anxiety Levels Among Health Care Workers at the Height of the COVID-19 Pandemic in the United Arab Emirates. International journal of public health 66, 1604369.

Sadiq, R., Faizan, M., Wadood, F., Malik, M.A., Mustafa, J., Waheed, U., 2021. Mental Health Status of Health Care Professionals During COVID-19 Pandemic at a Tertiary Care Hospital of Central Punjab. PAKISTAN JOURNAL OF MEDICAL & HEALTH SCIENCES 15, 3132-3135.

Sagherian, K., Steege, L.M., Cobb, S.J., Cho, H., 2020. Insomnia, fatigue and psychosocial well-being during COVID-19 pandemic: A cross-sectional survey of hospital nursing staff in the United States. Journal of clinical nursing.

Sahin, S.K., Arslan, E., Atalay, Ü.M., Demir, B., Elboga, G., Altındağ, A., 2021. Psychological impact of covid-19 outbreak on health workers in a university hospital in turkey. Psychology, Health & Medicine.

Sanghavi, P.B., Yeung, K.A., Sosa, C.E., Veesenmeyer, A.F., Limon, J.A., Vijayan, V., 2020. Effect of the Coronavirus Disease 2019 (COVID-19) Pandemic on Pediatric Resident Well-Being. Journal of Medical Education and Curricular Development 7.

Sangra, P.S., Mir, S.A., Ribeiro, T.C., Esteban-Sepulveda, S., Pages, E.G., Barbeito, B.L., Moya-Prats, J.L.P., Perez, L.P., Llobet, J.A., 2022. Mental health assessment of Spanish healthcare workers during the SARS-CoV-2 pandemic. A cross-sectional study. COMPREHENSIVE PSYCHIATRY 112.

Saracoglu, K.T., Simsek, T., Kahraman, S., Bombaci, E., Sezen, O., Saracoglu, A., Demirhan, R., 2020. The Psychological Impact of COVID-19 Disease is more Severe on Intensive Care Unit Healthcare Providers: A Cross-sectional Study. Clinical Psychopharmacology and Neuroscience 18, 607-615.

Schmid, B., Schulz, S.M., Schuler, M., Göpfert, D., Hein, G., Heuschmann, P., Wurmb, T., Pauli, P., Meybohm, P., Rittner, H.L., 2021. Impaired psychological well-being of healthcare workers in a German department of anesthesiology is independent of immediate SARS-CoV-2 exposure - a longitudinal observational study. German medical science : GMS e-journal 19, Doc11.

Schneider, J.N., Hiebel, N., Kriegsmann-Rabe, M., Schmuck, J., Erim, Y., Morawa, E., Jerg-Bretzke, L., Beschoner, P., Albus, C., Hannemann, J., Weidner, K., Steudte-Schmiedgen, S., Radbruch, L., Brunsch, H., Geiser, F., 2021. Moral Distress in Hospitals During the First Wave of the COVID-19 Pandemic: A Web-Based Survey Among 3,293 Healthcare Workers Within the German Network University Medicine. Frontiers in psychology 12, 775204.

Serrano, J., 2021. Depression and anxiety prevalence in nursing staff during the COVID-19 pandemic. Nursing Management 52, 24-32.

Shah, J., Monroe-Wise, A., Talib, Z., Nabiswa, A., Said, M., Abeid, A., Ali Mohamed, M., Mohamed, S., Ali, S.K., 2021. Mental health disorders among healthcare workers during the COVID-19 pandemic: a cross-sectional survey from three major hospitals in Kenya. BMJ open 11, e050316.

Shah, N., Raheem, A., Sideris, M., Velauthar, L., Saeed, F., 2020. Mental health amongst obstetrics and gynaecology doctors during the COVID-19 pandemic: Results of a UK-wide study. European Journal of Obstetrics and Gynecology and Reproductive Biology 253, 90-94.

Shalhub, S., Mouawad, N.J., Malgor, R.D., Johnson, A.P., Wohlauer, M.V., Coogan, S.M., Loveland, K.A., Cuff, R.F., Leonardi, C., Coleman, D.M., Sheahan, M.G., Woo, K., 2020. Global vascular surgeons' experience, stressors, and coping during the coronavirus disease 2019 pandemic. Journal of Vascular Surgery.

Sharma, S.K., Mudgal, S.K., Thakur, K., Parihar, A., Chundawat, D.S., Joshi, J., 2021. Anxiety, depression and quality of life (QOL) related to COVID-19 among frontline health care professionals: A multicentric cross-sectional survey. Journal of family medicine and primary care 10, 1383-1389.

Shechter, A., Chiuzan, C., Shang, Y., Ko, G., Diaz, F., Venner, H.K., Shaw, K., Cannone, D.E., McMurry, C.L., Sullivan, A.M., Rivera, R.R., Vose, C., Shapiro, P.A., Abdalla, M., 2021. Prevalence, Incidence, and Factors Associated with Posttraumatic Stress at Three-Month Follow-Up among New York City Healthcare Workers after the First Wave of the COVID-19 Pandemic. International journal of environmental research and public health 19.

Shechter, A., Diaz, F., Moise, N., Anstey, D.E., Ye, S., Agarwal, S., Birk, J.L., Brodie, D., Cannone, D.E., Chang, B., Claassen, J., Cornelius, T., Derby, L., Dong, M., Givens, R.C., Hochman, B., Homma, S., Kronish, I.M., Lee, S.A.J., Manzano, W., Mayer, L.E.S., McMurry, C.L., Moitra, V., Pham, P., Rabbani, L., Rivera, R.R., Schwartz, A., Schwartz, J.E., Shapiro, P.A., Shaw, K., Sullivan, A.M., Vose, C., Wasson, L., Edmondson, D., Abdalla, M., 2020. Psychological distress, coping behaviors, and preferences for support among New York healthcare workers during the COVID-19 pandemic. General Hospital Psychiatry 66, 1-8.

Shen, M., Xu, H., Fu, J., Wang, T., Fu, Z., Zhao, X., Zhou, G., Jin, Q., Tong, G., 2020. Investigation of anxiety levels of 1637 healthcare workers during the epidemic of COVID-19. PLoS ONE 15.

Si, M.Y., Su, X.Y., Jiang, Y., Wang, W.J., Gu, X.F., Ma, L., Li, J., Zhang, S.K., Ren, Z.F., Ren, R., Liu, Y.L., Qiao, Y.L., 2020. Psychological impact of COVID-19 on medical care workers in China. Infectious Diseases of Poverty 9.

Sim, S.K., Lau, B.L., Zaila, S.R., Hazira, N., Aniqah, N.M., Panicker, J., Hamzah, A.S., 2021. Psychological symptoms among healthcare workers handling COVID-19 patients. The Medical journal of Malaysia 76, 138-144.

Simonetti, V., Durante, A., Ambrosca, R., Arcadi, P., Graziano, G., Pucciarelli, G., Simeone, S., Vellone, E., Alvaro, R., Cicolini, G., 2021. Anxiety, sleep disorders and self-efficacy among nurses during COVID-19 pandemic: A large cross-sectional study. Journal of clinical nursing 30, 1360-1371.

Smith, A.J., Wright, H., Griffin, B.J., Ehman, A.C., Shoji, K., Love, T.M., Morrow, E., Locke, A., Call, M., Kerig, P.K., Olff, M., Benight, C.C., Langenecker, S.A., 2021. Mental health risks differentially associated with immunocompromised status among healthcare workers and family members at the pandemic outset. Brain, behavior, & immunity - health 15, 100285.

Song, X., Fu, W., Liu, X., Luo, Z., Wang, R., Zhou, N., Yan, S., Lv, C., 2020. Mental health status of medical staff in emergency departments during the Coronavirus disease 2019 epidemic in China. Brain, Behavior, and Immunity 88, 60-65.

Stocchetti, N., Segre, G., Zanier, E.R., Zanetti, M., Campi, R., Scarpellini, F., Clavenna, A., Bonati, M., 2021. Burnout in Intensive Care Unit Workers during the Second Wave of the COVID-19 Pandemic: A Single Center Cross-Sectional Italian Study. International journal of environmental research and public health 18.

Stojanov, J., Malobabic, M., Stanojevic, G., Stevic, M., Milosevic, V., Stojanov, A., 2020. Quality of sleep and health-related quality of life among health care professionals treating patients with coronavirus disease-19. The International journal of social psychiatry, 20764020942800.

Styra, R., Hawryluck, L., Mc Geer, A., Dimas, M., Sheen, J., Giacobbe, P., Dattani, N., Lorello, G., Rac, V.E., Francis, T., Wu, P.E., Luk, W.-S., Ng, E., Nadarajah, J., Wingrove, K., Gold, W.L., 2021. Surviving SARS and living through COVID-19: Healthcare worker mental health outcomes and insights for coping. PloS one 16, e0258893.

Su, Q., Ma, X., Liu, S., Liu, S., Goodman, B.A., Yu, M., Guo, W., 2021. Adverse Psychological Reactions and Psychological Aids for Medical Staff During the COVID-19 Outbreak in China. Frontiers in psychiatry 12, 580067.

Sun, H., Wang, S., Wang, W., Han, G., Liu, Z., Wu, Q., Pang, X., 2020. Correlation between emotional intelligence and negative emotions of front-line nurses during the COVID-19 epidemic: a cross-sectional study. Journal of clinical nursing.

Sung, C.-W., Chen, C.-H., Fan, C.-Y., Chang, J.-H., Hung, C.C., Fu, C.-M., Wong, L.P., Huang, E.P.-C., Lee, T.S.-H., 2021. Mental health crisis in healthcare providers in the COVID-19 pandemic: a cross-sectional facility-based survey. BMJ open 11, e052184.

Suryavanshi, N., Kadam, A., Dhumal, G., Nimkar, S., Mave, V., Gupta, A., Cox, S.R., Gupte, N., 2020. Mental health and quality of life among healthcare professionals during the COVID‐19 pandemic in India. Brain and Behavior 10.

Takashi, O., Eiichi, T., Yuka, O., Kazuko, M., Ohue, T., Togo, E., Ohue, Y., Mitoku, K., 2021. Mental health of nurses involved with COVID-19 patients in Japan, intention to resign, and influencing factors. Medicine 100, 1-9.

Tamrakar, P., Pant, S.B., Acharya, S.P., 2021. Anxiety and depression among nurses in COVID and non-COVID intensive care units. Nursing in critical care.

Tatsuno, J., Unoki, T., Sakuramoto, H., Hamamoto, M., 2021. Effects of social support on mental health for critical care nurses during the coronavirus disease 2019 (COVID-19) pandemic in Japan: A web-based cross-sectional study. Acute medicine & surgery 8, e645.

Teo, I., Chay, J., Cheung, Y.B., Sung, S.C., Tewani, K.G., Yeo, L.F., Yang, G.M., Pan, F.T., Ng, J.Y., Abu Bakar Aloweni, F., Ang, H.G., Ayre, T.C., Chai-Lim, C., Chen, R.C., Heng, A.L., Nadarajan, G.D., Ong, M.E.H., See, B., Soh, C.R., Tan, B.K.K., Tan, B.S., Tay, K.X.K., Wijaya, L., Tan, H.K., 2021. Healthcare worker stress, anxiety and burnout during the COVID-19 pandemic in Singapore: A 6-month multi-centre prospective study. PloS one 16, e0258866.

Teo, W.Z.Y., Soo, Y.E., Yip, C., Lizhen, O., Chun-Tsu, L., 2020. The psychological impact of COVID-19 on 'hidden' frontline healthcare workers. The International journal of social psychiatry, 20764020950772.

Than, H.M., Nong, V.M., Nguyen, C.T., Dong, K.P., Ngo, H.T., Doan, T.T., Do, N.T., Nguyen, T.H.T., Van Do, T., Dao, C.X., Nguyen, T.Q., Pham, T.N., Do, C.D., 2020. Mental health and health-related quality-of-life outcomes among frontline health workers during the peak of covid-19 outbreak in Vietnam: A cross-sectional study. Risk Management and Healthcare Policy 13, 2927-2936.

Th'ng, F., Rao, K.A., Ge, L., Mao, D., Neo, H.N., Molina, J.A.D., Seow, E., 2021. A One-Year Longitudinal Study: Changes in Depression and Anxiety in Frontline Emergency Department Healthcare Workers in the COVID-19 Pandemic. International journal of environmental research and public health 18.

Thomaier, L., Teoh, D., Jewett, P., Beckwith, H., Parsons, H., Yuan, J., Blaes, A.H., Lou, E., Hui, J.Y.C., Vogel, R.I., 2020. Emotional health concerns of oncology physicians in the United States: fallout during the COVID-19 pandemic. MedRxiv : the preprint server for health sciences.

Tiete, J., Guatteri, M., Lachaux, A., Matossian, A., Hougardy, J.-M., Loas, G., Rotsaert, M., 2021. Mental Health Outcomes in Healthcare Workers in COVID-19 and Non-COVID-19 Care Units: A Cross-Sectional Survey in Belgium. Frontiers in psychology 11, 612241.

Tran, T.V., Nguyen, H.C., Pham, L.V., Nguyen, M.H., Ha, T.H., Phan, D.T., Dao, H.K., Nguyen, P.B., Trinh, M.V., Do, T.V., Nguyen, H.Q., Nguyen, T.T.P., Nguyen, N.P.T., Tran, C.Q., Tran, K.V., Duong, T.T., Pham, H.X., Nguyen, L.V., Vo, T.T., Do, B.N., Duong, T.H., Pham, M.K., Pham, T.T.M., Nguyen, K.T., Yang, S.H., Chao, J.C.J., Duong, T.V., 2020. Impacts and interactions of COVID-19 response involvement, health-related behaviours, health literacy on anxiety, depression and health-related quality of life among healthcare workers: A cross-sectional study. BMJ Open 10.

Tselebis, A., Lekka, D., Sikaras, C., Tsomaka, E., Tassopoulos, A., Ilias, I., Bratis, D., Pachi, A., 2020. Insomnia, Perceived Stress, and Family Support among Nursing Staff during the Pandemic Crisis. Healthcare (Basel, Switzerland) 8.

Tuan, N.Q., Phuong, N.D., Co, D.X., Son, D.N., Chinh, L.Q., Dung, N.H., Thach, P.T., Thai, N.Q., Thu, T.A., Tuan, N.A., San, B.V., Tung, V.S., An, N.V., Khanh, D.N., Long, V.H., Tai, N., Muoi, T., Vinh, N.D., Thien, N.T., Nhan, L.D., Tuan, N.V., 2021. Prevalence and Factors Associated with Psychological Problems of Healthcare Workforce in Vietnam: Findings from COVID-19 Hotspots in the National Second Wave. Healthcare (Basel, Switzerland) 9.

Turkmen, E., Kir, S., Sayarlioglu, H., Dilek, M., Arik, N., 2021. Prevalence and Influencing Factors of Anxiety in Healthcare Professionals in Hemodialysis During the COVID-19 Pandemic. TURKISH JOURNAL OF NEPHROLOGY 30, 158-164.

Uvais, N.A., Nalakath, M.J., Jose, K., 2021. Facing COVID-19: Psychological Impacts on Hospital Staff in a Tertiary Care Private Hospital in India. The primary care companion for CNS disorders 23.

Uyaroğlu, O.A., Başaran, N.Ç., Ozisik, L., Karahan, S., Tanriover, M.D., Guven, G.S., Oz, S.G., 2020. Evaluation of the effect of COVID-19 pandemic on anxiety severity of physicians working in the internal medicine department of a tertiary care hospital: a cross-sectional survey. Internal Medicine Journal.

Uz, B., Savasan, E., Soganci, D., 2022. Anxiety, Depression and Burnout Levels of Turkish Healthcare Workers at the End of the First Period of COVID-19 Pandemic in Turkey. CLINICAL PSYCHOPHARMACOLOGY AND NEUROSCIENCE 20, 97-108.

Vafaei, H., Roozmeh, S., Hessami, K., Kasraeian, M., Asadi, N., Faraji, A., Bazrafshan, K., Saadati, N., Aski, S.K., Zarean, E., Golshahi, M., Haghiri, M., Abdi, N., Tabrizi, R., Heshmati, B., Arshadi, E., 2020. Obstetrics Healthcare Providers' Mental Health and Quality of Life During COVID-19 Pandemic: Multicenter Study from Eight Cities in Iran. Psychology Research and Behavior Management 13, 563-571.

Valaine, L., Ancāne, G., Utināns, A., Briģis, Ģ., 2021. Mental Health and Associated Demographic and Occupational Factors among Health Care Workers during the COVID-19 Pandemic in Latvia. Medicina (Kaunas, Lithuania) 57.

Van Wert, M.J., Gandhi, S., Gupta, I., Singh, A., Eid, S.M., Burhanullah, M.H., Michtalik, H., Malik, M., Healthcare Worker Mental Health After the Initial Peak of the COVID-19 Pandemic: a US Medical Center Cross-Sectional Survey. JOURNAL OF GENERAL INTERNAL MEDICINE.

Vanni, G., Materazzo, M., Santori, F., Pellicciaro, M., Costesta, M., Orsaria, P., Cattadori, F., Pistolese, C.A., Perretta, T., Chiocchi, M., Meucci, R., Lamacchia, F., Assogna, M., Caspi, J., Granai, A.V., De Majo, A., Chiaravalloti, A., D'Angelillo, M.R., Barbarino, R., Ingallinella, S., Morando, L., Dalli, S., Portarena, I., Altomare, V., Tazzioli, G., Buonomo, O.C., 2020. The Effect of coronavirus (COVID-19) on breast cancer teamwork: A multicentric survey. In Vivo 34, 1685-1694.

Villalba-Arias, J., Estigarribia, G., Bogado, J.A., Méndez, J., Toledo, S., Barrios, I., Castaldelli-Maia, J.M., Ventriglio, A., Torales, J., 2021. Mental health issues and psychological risk factors among Paraguayan healthcare workers during the COVID-19 pandemic. Journal of mental health (Abingdon, England), 1-8.

Vitale, E., Galatola, V., Mea, R., 2021. Observational study on the potential psychological factors that affected Italian nurses involved in the COVID-19 health emergency. Acta bio-medica : Atenei Parmensis 92, e2021007.

Vitale, E., Mea, R., Di Dio, F., Canonico, A., Galatola, V., 2020. Anxiety, Insomnia and Body Mass Index scores in Italian nurses engaged in the care of COVID-19 patients. Endocrine, metabolic & immune disorders drug targets.

Wadasadawala, T., Kumar, A., Laskar, S.G., Gondhowiardjo, S., Mokal, S., Goswami, S., Giselvania, A., Kapoor, R., Das, A., Pradhan, S., Pujari, L., Acharya, B., Chapagain, S., Mahantshetty, U., Vadgaonkar, R., Hussain, Q.M., Akbarov, K., Agarwal, J.P., 2021. Multinational Study to Assess Stress Levels Among the Health Care Workers of Radiation Oncology Community at the Outset of the COVID-19 Pandemic. JCO global oncology 7, 464-473.

Walvik, L., Brejnebøl, M.W., Ravn, A.T., Jensen, R.G., Christensen, A.K., von Buchwald, C., 2021. The impact of the COVID-19 pandemic on mental health among healthcare workers in ear-nose-throat clinics. Danish medical journal 68.

Wang, H., Huang, D., Huang, H., Zhang, J., Guo, L., Liu, Y., Ma, H., Geng, Q., 2020. The psychological impact of COVID-19 pandemic on medical staff in Guangdong, China: a cross-sectional study. Psychological medicine, 1-9.

Wang, L., Xu, X., Shi, L., Hong, S., Wang, W., Fang, X., Chen, J., Zhang, Q., Ai, M., Kuang, L., 2022. A cross-sectional study of the psychological status of 33,706 hospital workers at the late stage of the COVID-19 outbreak. JOURNAL OF AFFECTIVE DISORDERS 297, 156-168.

Wang, L.Q., Zhang, M., Liu, G.M., Nan, S.Y., Li, T., Xu, L., Xue, Y., Wang, L., Qu, Y.D., Liu, F., 2020. Psychological impact of coronavirus disease (2019) (COVID-19) epidemic on medical staff in different posts in China: A multicenter study. Journal of Psychiatric Research 129, 198-205.

Wang, Q.Q., Fang, Y.Y., Huang, H.L., Lv, W.J., Wang, X.X., Yang, T.T., Yuan, J.M., Gao, Y., Qian, R.L., Zhang, Y.H., 2021. Anxiety, depression and cognitive emotion regulation strategies in Chinese nurses during the COVID‐19 outbreak. Journal of Nursing Management (John Wiley & Sons, Inc.) 29, 1263-1274.

Wang, S., Xie, L., Xu, Y., Yu, S., Yao, B., Xiang, D., 2020. Sleep disturbances among medical workers during the outbreak of COVID-2019. Occupational Medicine 70, 364-369.

Wang, S., Zhou, P., Yang, X., Wang, N., Jie, J., Li, Y., Cai, Q., Lyv, C., Li, Y., 2021. Can urban prosperity aid in recovery? The relationship between healthcare workers' mental health status and the city level during the COVID-19 epidemic. CITIES 118.

Wang, W., Song, W., Xia, Z., He, Y., Tang, L., Hou, J., Lei, S., 2020. Sleep Disturbance and Psychological Profiles of Medical Staff and Non-Medical Staff During the Early Outbreak of COVID-19 in Hubei Province, China. Frontiers in Psychiatry 11.

Wang, Y., Duan, Z., Peng, K., Li, D., Ou, J., Wilson, A., Wang, N., Si, L., Chen, R., 2020. Acute stress disorder among frontline health professionals during the COVID-19 outbreak: A structural equation modelling investigation. Psychosomatic medicine.

Wang, Y., Ma, S., Yang, C., Cai, Z., Hu, S., Zhang, B., Tang, S., Bai, H., Guo, X., Wu, J., Du, H., Kang, L., Tan, H., Li, R., Yao, L., Wang, G., Liu, Z., 2020. Acute psychological effects of Coronavirus Disease 2019 outbreak among healthcare workers in China: a cross-sectional study. Translational Psychiatry 10.

Wang, Y.X., Guo, H.T., Du, X.W., Song, W., Lu, C., Hao, W.N., 2020. Factors associated with post-traumatic stress disorder of nurses exposed to corona virus disease 2019 in China. Medicine 99.

Wanigasooriya, K., Palimar, P., Naumann, D.N., Ismail, K., Fellows, J.L., Logan, P., Thompson, C.V., Bermingham, H., Beggs, A.D., Ismail, T., 2020. Mental health symptoms in a cohort of hospital healthcare workers following the first peak of the COVID-19 pandemic in the UK. BJPsych Open 7.

Wilson, W., Raj, J.P., Rao, S., Ghiya, M., Nedungalaparambil, N.M., Mundra, H., Mathew, R., 2020. Prevalence and Predictors of Stress, anxiety, and Depression among Healthcare Workers Managing COVID-19 Pandemic in India: A Nationwide Observational Study. Indian journal of psychological medicine 42, 353-358.

Woon, L.S.C., Sidi, H., Jaafar, N.R.N., Bin Abdullah, M.F.I.L., 2020. Mental health status of university healthcare workers during the covid-19 pandemic: A post–movement lockdown assessment. International Journal of Environmental Research and Public Health 17, 1-20.

Wozniak, H., Benzakour, L., Moullec, G., Buetti, N., Nguyen, A., Corbaz, S., Roos, P., Vieux, L., Suard, J.-C., Weissbrodt, R., Pugin, J., Pralong, J.A., Cereghetti, S., 2021. Mental health outcomes of ICU and non-ICU healthcare workers during the COVID-19 outbreak: a cross-sectional study. Annals of intensive care 11, 106.

Wright, H.M., Griffin, B.J., Shoji, K., Love, T.M., Langenecker, S.A., Benight, C.C., Smith, A.J., 2020. Pandemic-related mental health risk among front line personnel. Journal of Psychiatric Research.

Xiao, X., Zhu, X., Fu, S., Hu, Y., Li, X., Xiao, J., 2020. Psychological impact of healthcare workers in China during COVID-19 pneumonia epidemic: A multi-center cross-sectional survey investigation. Journal of Affective Disorders 274, 405-410.

Xiaoming, X., Ming, A., Su, H., Wo, W., Jianmei, C., Qi, Z., Hua, H., Xuemei, L., Lixia, W., Jun, C., Lei, S., Zhen, L., Lian, D., Jing, L., Handan, Y., Haitang, Q., Xiaoting, H., Xiaorong, C., Ran, C., Qinghua, L., Xinyu, Z., Jian, T., Jing, T., Guanghua, J., Zhiqin, H., Nkundimana, B., Li, K., 2020. The psychological status of 8817 hospital workers during COVID-19 Epidemic: A cross-sectional study in Chongqing. Journal of Affective Disorders 276, 555-561.

Xing, L.-Q., Xu, M.-L., Sun, J., Wang, Q.-X., Ge, D.-D., Jiang, M.-M., Du, W., Li, Q., 2020. Anxiety and depression in frontline health care workers during the outbreak of Covid-19. The International journal of social psychiatry, 20764020968119.

Xu, L., You, D., Li, C., Zhang, X., Yang, R., Kang, C., Wang, N., Jin, Y., Yuan, J., Li, C., Wei, Y., Li, Y., Yang, J., 2022. Two-stage mental health survey of first-line medical staff after ending COVID-19 epidemic assistance and isolation. EUROPEAN ARCHIVES OF PSYCHIATRY AND CLINICAL NEUROSCIENCE 272, 81-93.

Xu, X., Wang, W., Chen, J., Ai, M., Shi, L., Wang, L., Hong, S., Zhang, Q., Hu, H., Li, X., Cao, J., Lv, Z., Du, L., Li, J., Yang, H., He, X., Chen, X., Chen, R., Luo, Q., Zhou, X., Tan, J., Tu, J., Jiang, G., Han, Z., Kuang, L., 2021. Suicidal and self-harm ideation among Chinese hospital staff during the COVID-19 pandemic: Prevalence and correlates. Psychiatry research 296, 113654.

Yadeta, T.A., Dessie, Y., Balis, B., 2021. Magnitude and Predictors of Health Care Workers Depression During the COVID-19 Pandemic: Health Facility-Based Study in Eastern Ethiopia. Frontiers in psychiatry 12, 654430.

Yang, H., Shi, R., Chi, Y., Qiao, Z., Wu, Y., Zhu, Z., Xiao, B., Feng, L., Wang, H., 2021. Knowledge, Anxiety, Depression, and Sleep Quality Among Medical Staff in Central South Areas of China During the Break of COVID-19: Does the Level of Hospitals Make a Difference? FRONTIERS IN PSYCHIATRY 12.

Yang, S., Kwak, S.G., Chang, M.C., Psychological impact of COVID-19 on hospital workers in nursing care hospitals. Nursing Open.

Yang, S., Kwak, S.G., Ko, E.J., Chang, M.C., 2020. The mental health burden of the covid-19 pandemic on physical therapists. International Journal of Environmental Research and Public Health 17.

Yang, X., Chen, D., Chen, Y., Wang, N., Lyv, C., Li, Y., Jie, J., Zhou, T., Li, Y., Zhou, P., 2021. Geographical distribution and prevalence of mental disorders among healthcare workers in China: A cross-sectional country-wide survey: A cross-sectional study to assess mental disorders of healthcare workers in China. International Journal of Health Planning & Management 36, 1561-1574.

Yang, X.P., Zhang, Y., Li, S., Chen, X., Risk factors for anxiety of otolaryngology healthcare workers in Hubei province fighting coronavirus disease 2019 (COVID-19). Social Psychiatry and Psychiatric Epidemiology.

Yang, Y., Lu, L., Chen, T., Ye, S., Kelifa, M.O., Cao, N., Zhang, Q., Liang, T., Wang, W., 2021. Healthcare Worker's Mental Health and Their Associated Predictors During the Epidemic Peak of COVID-19. Psychology research and behavior management 14, 221-231.

Yegin, G.F., Desdicioglu, R., Secen, E.I., Aydin, S., Bal, C., Goka, E., Keskin, H.L., 2022. Low Anti-Mullerian Hormone Levels Are Associated with the Severity of Anxiety Experienced by Healthcare Professionals During the COVID-19 Pandemic. REPRODUCTIVE SCIENCES 29, 627-632.

Yeo, I.-H., Kim, Y.-J., Kim, J.-K., Lee, D.-E., Choe, J.-Y., Kim, C.-H., Park, J.-B., Seo, K.-S., Park, S.-Y., Lee, S.-H., Cho, J.-K., Lee, S.-H., 2021. Impact of the COVID-19 Pandemic on Emergency Department Workload and Emergency Care Workers' Psychosocial Stress in the Outbreak Area. Medicina (Kaunas, Lithuania) 57.

Yin, Q., Chen, A., Song, X., Deng, G., Dong, W., 2021. Risk Perception and PTSD Symptoms of Medical Staff Combating Against COVID-19: A PLS Structural Equation Model. Frontiers in psychiatry 12, 607612.

Yitayih, Y., Mekonen, S., Zeynudin, A., Mengistie, E., Ambelu, A., 2020. Mental health of healthcare professionals during the early stage of the COVID-19 pandemic in Ethiopia. BJPsych Open 7.

Yoruk, S., Guler, D., The relationship between psychological resilience, burnout, stress, and sociodemographic factors with depression in nurses and midwives during the COVID-19 pandemic: A cross-sectional study in Turkey. Perspectives in Psychiatric Care.

Youngrong, L., Kwanghyun, K., Sungjin, P., Sun Jae, J., Lee, Y., Kim, K., Park, S., Jung, S.J., 2021. Associations Between General Perceptions of COVID-19 and Posttraumatic Stress Disorder in Korean Hospital Workers: Effect Modification by Previous Middle East Respiratory Syndrome Coronavirus Experience and Occupational Type. Journal of Preventive Medicine & Public Health 54, 86-95.

Yue, L., Zhao, R., Xiao, Q., Zhuo, Y., Yu, J., Meng, X., 2021. The effect of mental health on sleep quality of front-line medical staff during the COVID-19 outbreak in China: A cross-sectional study. PloS one 16, e0253753.

Zakeri, M.A., Dehghan, M., Ghaedi Heidari, F., Pakdaman, H., Mehdizadeh, M., Ganjeh, H., Sanji Rafsanjani, M., Hossini Rafsanjanipoor, S.M., 2021. Mental health outcomes among health-care workers during the COVID-19 outbreak in Iran. Mental Health Review Journal 26, 152-160.

Zakeri, M.A., Hossini Rafsanjanipoor, S.M., Sedri, N., Kahnooji, M., Sanji Rafsanjani, M., Zakeri, M., Zakeri Bazmandeh, A., Talebi, A., Dehghan, M., 2021. Psychosocial status during the prevalence of covid-19 disease: The comparison between healthcare workers and general population. Current Psychology: A Journal for Diverse Perspectives on Diverse Psychological Issues.

Zakeri, M.A., Hossini Rafsanjanipoor, S.M., Zakeri, M., Dehghan, M., 2021. The relationship between frontline nurses' psychosocial status, satisfaction with life and resilience during the prevalence of COVID-19 disease. Nursing open 8, 1829-1839.

Zartash, S., Javed, M., Hashmi, M., Qayyum, A., Rana, M.A., Munir, F., 2020. COVID-19 Induced anxiety among health care professionals. Pakistan Journal of Medical and Health Sciences 14, 846-849.

Zebley, B., Wolk, D., McAllister, M., Lynch, C.J., Mikofsky, R., Liston, C., 2021. Individual Differences in the Affective Response to Pandemic-Related Stressors in COVID-19 Health Care Workers. Biological psychiatry global open science 1, 336-344.

Zhan, Y., Liu, Y., Liu, H., Li, M., Shen, Y., Gui, L., Zhang, J., Luo, Z., Tao, X., Yu, J., 2020. Factors associated with insomnia among Chinese front‐line nurses fighting against COVID‐19 in Wuhan: A cross‐sectional survey. Journal of Nursing Management 28, 1525-1535.

Zhang, C., Peng, D.H., Lv, L., Zhuo, K.M., Yu, K., Shen, T., Xu, Y.F., Wang, Z., 2020. Individual Perceived Stress Mediates Psychological Distress in Medical Workers During COVID-19 Epidemic Outbreak in Wuhan. Neuropsychiatric Disease and Treatment 16, 2529-2537.

Zhang, C., Yang, L., Liu, S., Ma, S., Wang, Y., Cai, Z., Du, H., Li, R., Kang, L., Su, M., Zhang, J., Liu, Z., Zhang, B., 2020. Survey of Insomnia and Related Social Psychological Factors Among Medical Staff Involved in the 2019 Novel Coronavirus Disease Outbreak. Frontiers in Psychiatry 11.

Zhang, H., Shi, Y., Jing, P., Zhan, P., Fang, Y., Wang, F., 2020. Posttraumatic stress disorder symptoms in healthcare workers after the peak of the COVID-19 outbreak: A survey of a large tertiary care hospital in Wuhan. Psychiatry Research 294.

Zhang, H.-H., Zhao, Y.-J., Wang, C., Zhang, Q., Yu, H.-Y., Cheung, T., Hall, B.J., An, F.-R., Xiang, Y.-T., 2021. Depression and its relationship with quality of life in frontline psychiatric clinicians during the COVID-19 pandemic in China: a national survey. International journal of biological sciences 17, 683-688.

Zhang, J., Wang, Y., Xu, J., You, H., Li, Y., Liang, Y., Li, S., Ma, L., Lau, J.T.-F., Hao, Y., Chen, S., Zeng, J., Li, J., Gu, J., 2021. Prevalence of mental health problems and associated factors among front-line public health workers during the COVID-19 pandemic in China: an effort-reward imbalance model-informed study. BMC psychology 9, 55.

Zhang, L., Wang, S., Shen, J., Wang, Y., Huang, X., Wu, F., Zheng, X., Zeng, P., Qiu, D., 2020. The mental health of Chinese healthcare staff in non-epicenter of COVID-19: a cross-sectional study. Annals of palliative medicine 9, 4127-4136.

Zhang, R., Hou, T., Kong, X., Wang, G., Wang, H., Xu, S., Xu, J., He, J., Xiao, L., Wang, Y., Du, J., Huang, Y., Su, T., Tang, Y., 2021. PTSD Among Healthcare Workers During the COVID-19 Outbreak: A Study Raises Concern for Non-medical Staff in Low-Risk Areas. Frontiers in psychiatry 12, 696200.

Zhang, X., Zhao, K., Zhang, G., Feng, R., Chen, J., Xu, D., Liu, X., Ngoubene-Atioky, A.J., Huang, H., Liu, Y., Chen, L., Wang, W., 2020. Occupational Stress and Mental Health: A Comparison Between Frontline Medical Staff and Non-frontline Medical Staff During the 2019 Novel Coronavirus Disease Outbreak. Frontiers in Psychiatry 11.

Zhang, X., Zou, R., Liao, X., Bernardo, A.B.I., Du, H., Wang, Z., Cheng, Y., He, Y., 2020. Perceived Stress, Hope, and Health Outcomes Among Medical Staff in China During the COVID-19 Pandemic. Frontiers in Psychiatry 11.

Zhang, Z., Hu, Y., Chen, Y., Liao, Z., Zheng, Y., Ding, L., 2021. Sleep disorders and related factors among frontline medical staff supporting Wuhan during the COVID-19 outbreak. Bulletin of the Menninger Clinic 85, 254-270.

Zheng, R., Zhou, Y., Fu, Y., Xiang, Q., Cheng, F., Chen, H., Xu, H., Fu, L., Wu, X., Feng, M., Ye, L., Tian, Y., Deng, R., Liu, S., Jiang, Y., Yu, C., Li, J., 2020. Prevalence and associated factors of depression and anxiety among nurses during the outbreak of COVID-19 in China: A cross-sectional study. International journal of nursing studies 114, 103809.

Zheng, R., Zhou, Y., Qiu, M., Yan, Y., Yue, J., Yu, L., Lei, X., Tu, D., Hu, Y., 2020. Prevalence and associated factors of depression, anxiety, and stress among Hubei pediatric nurses during COVID-19 pandemic. Comprehensive psychiatry 104, 152217.

Zhu, J., Sun, L., Zhang, L., Wang, H., Fan, A., Yang, B., Li, W., Xiao, S., 2020. Prevalence and Influencing Factors of Anxiety and Depression Symptoms in the First-Line Medical Staff Fighting Against COVID-19 in Gansu. Frontiers in Psychiatry 11.

Zhu, Z., Xu, S., Wang, H., Liu, Z., Wu, J., Li, G., Miao, J., Zhang, C., Yang, Y., Sun, W., Zhu, S., Fan, Y., Chen, Y., Hu, J., Liu, J., Wang, W., 2020. COVID-19 in Wuhan: Sociodemographic characteristics and hospital support measures associated with the immediate psychological impact on healthcare workers. EClinicalMedicine 24, 100443.
