## Supplements for "The prevalence of probable mental health disorders among hospital healthcare workers during COVID-19: A systematic review and meta-analysis": S3C Appendix.Study's Measurement tools and cut off scores.docx

**S3 Appendix. Measurement tools and cut off scores**

| Depression Measurement Tools | Sensitivity | Specificity |
| --- | --- | --- |
| Hospital Anxiety and Depression Scale - Depression Subscale ≥ 8 ^(1, 2)^ | 0.715 | 0.856 |
| Hospital Anxiety and Depression Scale- Depression Subscale ≥ 7 (Turkish sample) ^(3)^ | N/A | N/A |
| Patient Health Questionnaire- Nine item ≥ 10 ^(4)^ | 0.88 | 0.88 |
| Patient Health Questionnaire- Two item ≥ 2-3 ^(5, 6)^ | 0.79 | 0.86 |
| Zung Self-Rating Depression Scale ≥ 50 ^(7)^ | 86 (73 to 100) | 76 (57 to 95) |
| Zung Self-Rating Depression Scale ≥ 53 (Chinese population) ^(8, 9)^ | 96.5 | 59.3 |
| Center for Epidemiological Studies- Depression ≥ 16 ^(7)^ | 84 (70 -89) | 74 (68-80) |
| Center for Epidemiological Studies- Depression ≥ 10 ^(7, 10)^ | .89 | .47 |
| Depression Anxiety Stress Scale- Depression Subscale ≥ 5 (Raw score)/ 10 and DASS-42-D ≥ 10 ^(11)^ | 0.91 | 0.40 |
| Beck Depression Inventory ≥ 10 ^(12)^ | 100 | 75 |
| Beck Depression Inventory ≥ 17 (Turkish population) ^(13)^ | N/A | N/A |
| Beck Depression Inventory -Second Edition ≥ 14 ^(14)^ | 0.79 | 0.76 |
| Goldberg Anxiety and Depression Scale-Depression Subscale ≥ 2 ^(15)^ | 0.85 | N/A |
| Case finding instruments : Positive response on both question/ ≥ 2^(16)^ | 75-96 | 57-88 |
| Major depression inventory ≥ 26 | 0.66 | 0.63 |
| HSCL-25 depression >1.75 ^(17, 18)^ | 0.84 | 0.48 |
| Inventory of Depressive Symptomology 30 (IDS-30)^(19)^ | 76 | 87 |

| Anxiety Measurement Tools | Sensitivity | Specificity |
| --- | --- | --- |
| Hospital Anxiety and Depression Scale – Anxiety Subscale ≥ 8 ^(1)^ | 0.780 | 0.742 |
| Hospital Anxiety and Depression Scale- Anxiety Subscale ≥ 10 (Turkish sample) ^(3)^ | N/A | N/A |
| General Anxiety Disorder-Seven Item ≥ (8 – 10) ^(20, 21)^ | 0.68 | 0.88 |
| General Anxiety Disorder-Two Item ≥ 3 ^(22)^ | 0.80 | 0.81 |
| Zung Self-Rating Depression Scale ≥ 50 ^(9)^ | 62.5 | 90.4 |
| Depression Anxiety Stress Scale- Anxiety Subscale ≥ 4 (Raw score) and DASS-42-D ≥ 8 (Raw score) ^(11)^ | 0.76 | 0.54 |
| Beck Anxiety Inventory ≥ 16 ^(23)^ | 0.62 | 0.69 |
| Goldberg Anxiety and Depression Scale-Anxiety Subscale ≥ 5 ^(24)^ | 0.91 | 0.68 |
| HSCL-25 anxiety >1.75 ^(17, 18)^ | 0.86 | 0.3 |
| PROMIS: T-Score ≥ 62.3 ^(25)^ | N/A | N/A |
| LEEDS SAD ≥ 7 ^(26)^ | N/A | N/A |

| PTSD Measurement tools | Sensitivity | Specificity |
| --- | --- | --- |
| Impact of Event Scale ≥ (24 – 27)^(27)^ | 0.87 to 0.80 | 0.87 to 0.93 |
| Impact of Event Scale- Revised ≥ 33 ^(28)^ | 0.91 | 0.82 |
| Impact of Event Scale- Revised (Korean population) ≥ (25 – 26) ^(29)^ | 0.90-0.93 | 0.81-0.82 |
| Impact of Event Scale- Six Item≥ 10/1.75 ^(30, 31)^ | 0.8 | 0.92 |
| PTSD Checklist for DSM-5 ≥ 31-33 ^(32)^ | 0.88 | 0.69 |
| Turkish PTSD Checklist for DSM-5 ≥ 47 ^(33)^ | .76 | .69 |
| PTSD Checklist – Civilian Version ≥ (38- 50) ^(34)^ | 0.52-0.79 | 0.93-0.80 |
| Acute Stress Disorder Scale ≥ 56 ^(35)^ | 0.91 | 0.93 |
| PTSD Checklist – Civilian Version- Six item ≥ 14 ^(36)^ | 0.92 | 0.72 |
| Trauma Screening Questionnaire ≥ 6 ^(37)^ | 0.86 | 0.93 |
| Short Post-Traumatic Stress Disorder Rating Interview ≥ 14 ^(38)^ | 0.95 | 9.96 |
| primary care PTSD screen ≥ 3 ^(39)^ | 0.95 | 0.85 |
| Stanford Acute Stress Reaction Questionnaire ≥ 3 ^(40)^ | 0.45 | 0.91 |
| The trauma and loss spectrum (TALS-SR): A full diagnosis of PTSD was defined as meeting all the  DSM-5 criteria B, C, D, E for PTSD. ^(41)^ | N/A | N/A |
| Post traumatic diagnostic scale- 5 ≥ 28 ^(42)^ | .79 | .77 |
| PTSD-SS/ NSESSS-PTSD ≥ 24 ^(43)^ | .91 | .77 |
| Essen trauma inventory ≥ 27 ^(44)^ | N/A | N/A |
| DTS ≥ 40 ^(45)^ | N/A | N/A |

| Insomnia Measurement Tools | Sensitivity | Specificity |
| --- | --- | --- |
| Insomnia Severity Index ≥ 14-15 ^(46)^ | 0.765-0.824 | 0.821 |
| Insomnia Severity Index ≥ 10 ^(47)^ | 86.1 | 87.7 |
| Athens insomnia index ≥6 ^(48)^ | 93 | 85 |
| Sleep Condition Indicator ≥16 ^(49)^ | 0.95 | 0.75 |

| Alcohol Measurement Tools | Sensitivity | Specificity |
| --- | --- | --- |
| Alcohol Use Disorders Identification Test Consumption ≥ 3 women and ≥ 4 men ^(50)^ | 0.86 | 0.89 |
| CAGE-AID ≥ 2/3 ^(51)^ | N/A | N/A |
