## Supplements for "The prevalence of probable mental health disorders among hospital healthcare workers during COVID-19: A systematic review and meta-analysis": S9 Appendix. Funnel plots.docx

**S8A Appendix.** **Funnel plot of depression estimates**

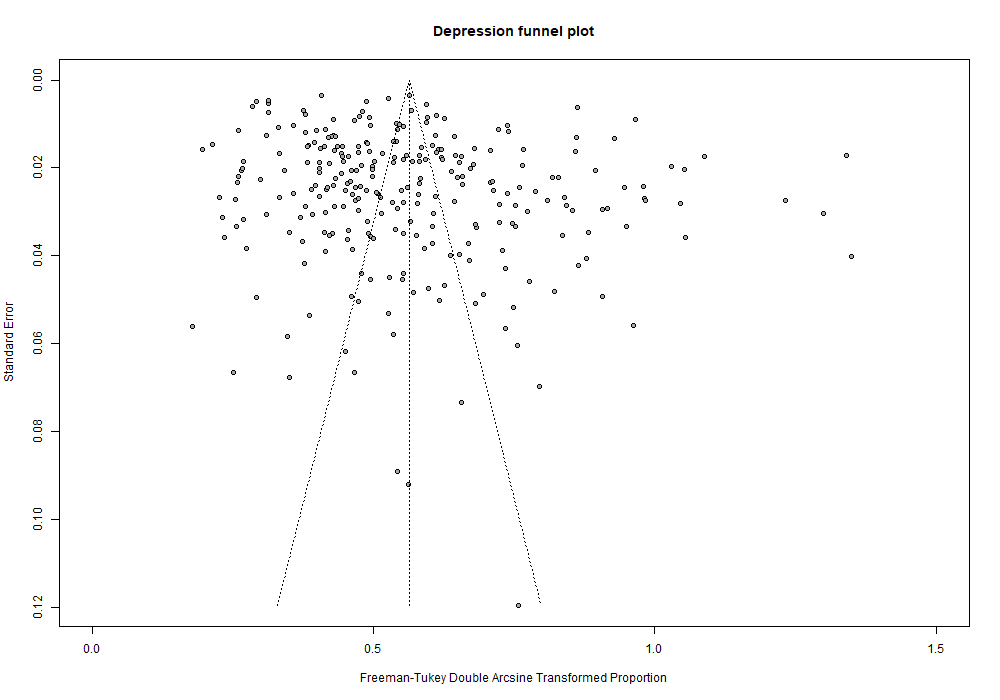

Egger’s test: p < 0.0001

**S8B Appendix.** **Funnel plot of anxiety estimates**

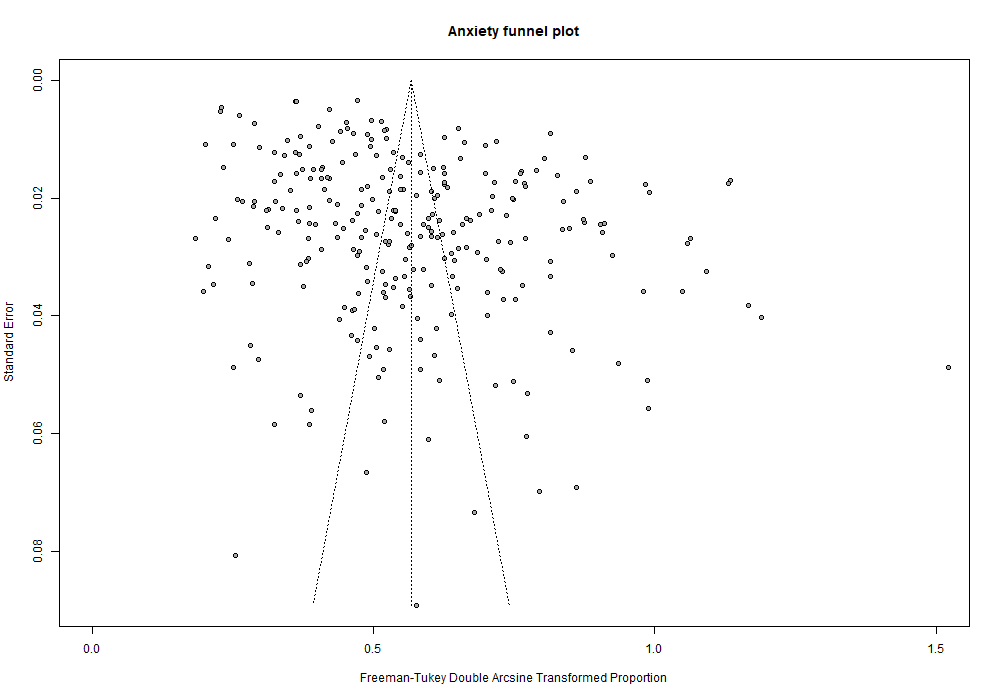

Egger’s test: p < 0.0001

**S8C Appendix.** **Funnel plot of PTSD estimates**

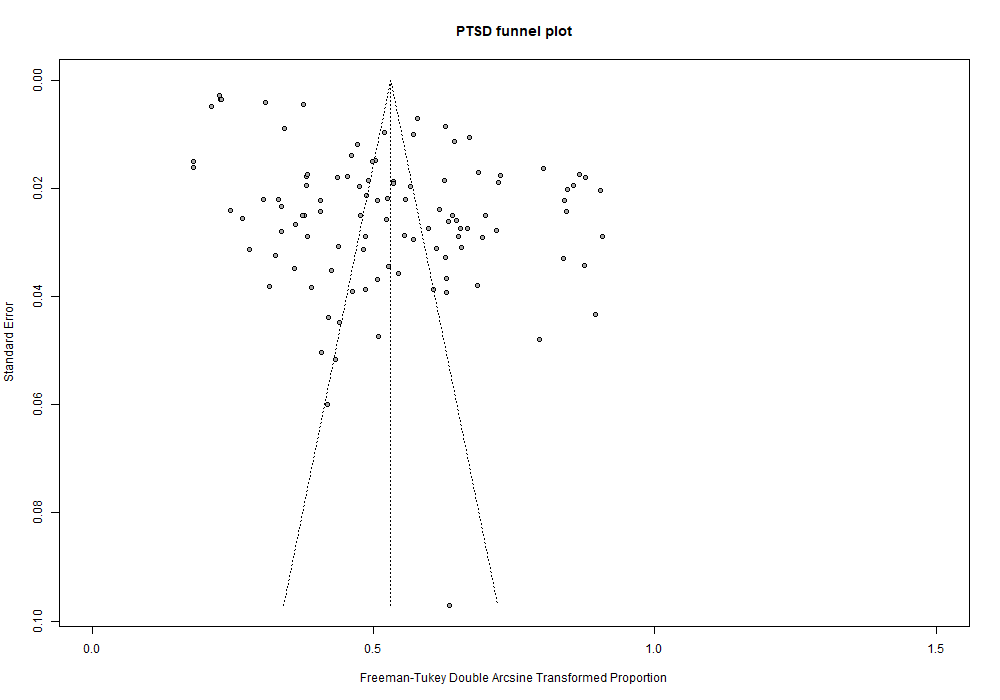

Egger’s test: p < 0.0001

**S8D Appendix.** **Funnel plot of insomnia estimates**

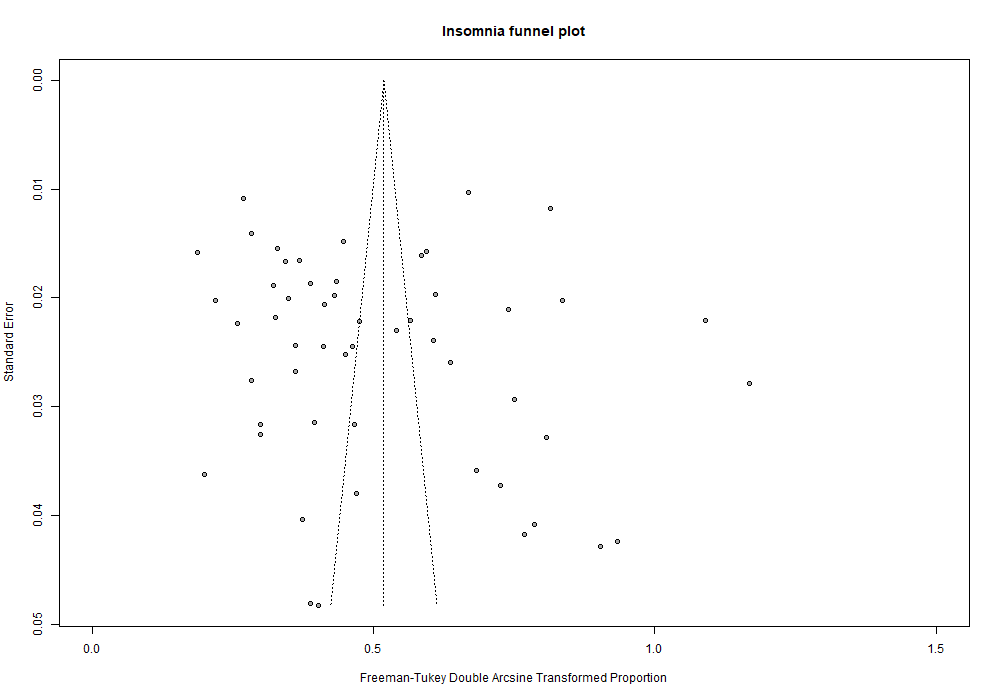

Egger’s test: p = 0.40
