## Supplementary figures and images for "The prevalence of probable mental health disorders among hospital healthcare workers during COVID-19: A systematic review and meta-analysis"

### S4A.Dep.Overall.tiff

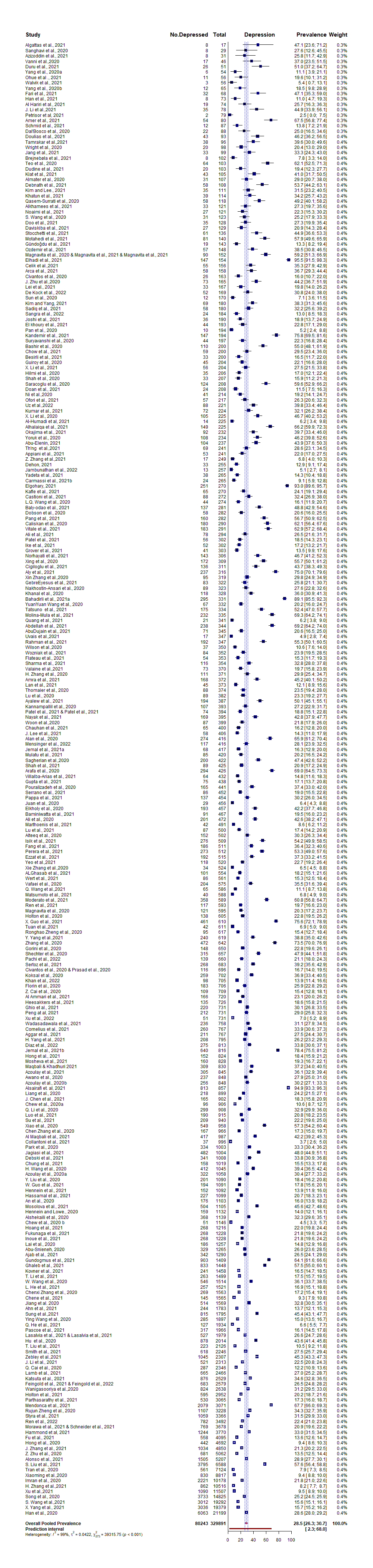

### S4B.Dep.Overall_by_Country.tiff

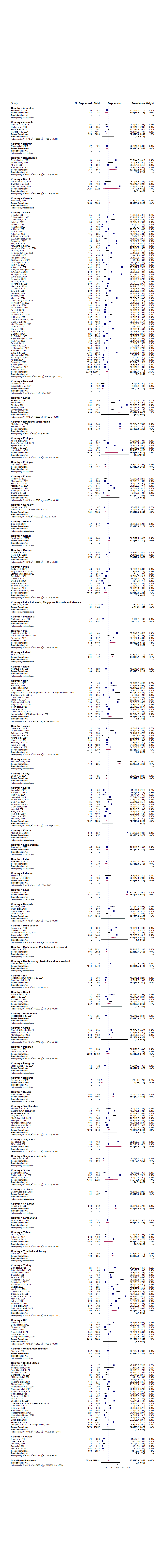

### S4C.Dep.Nurse.tiff

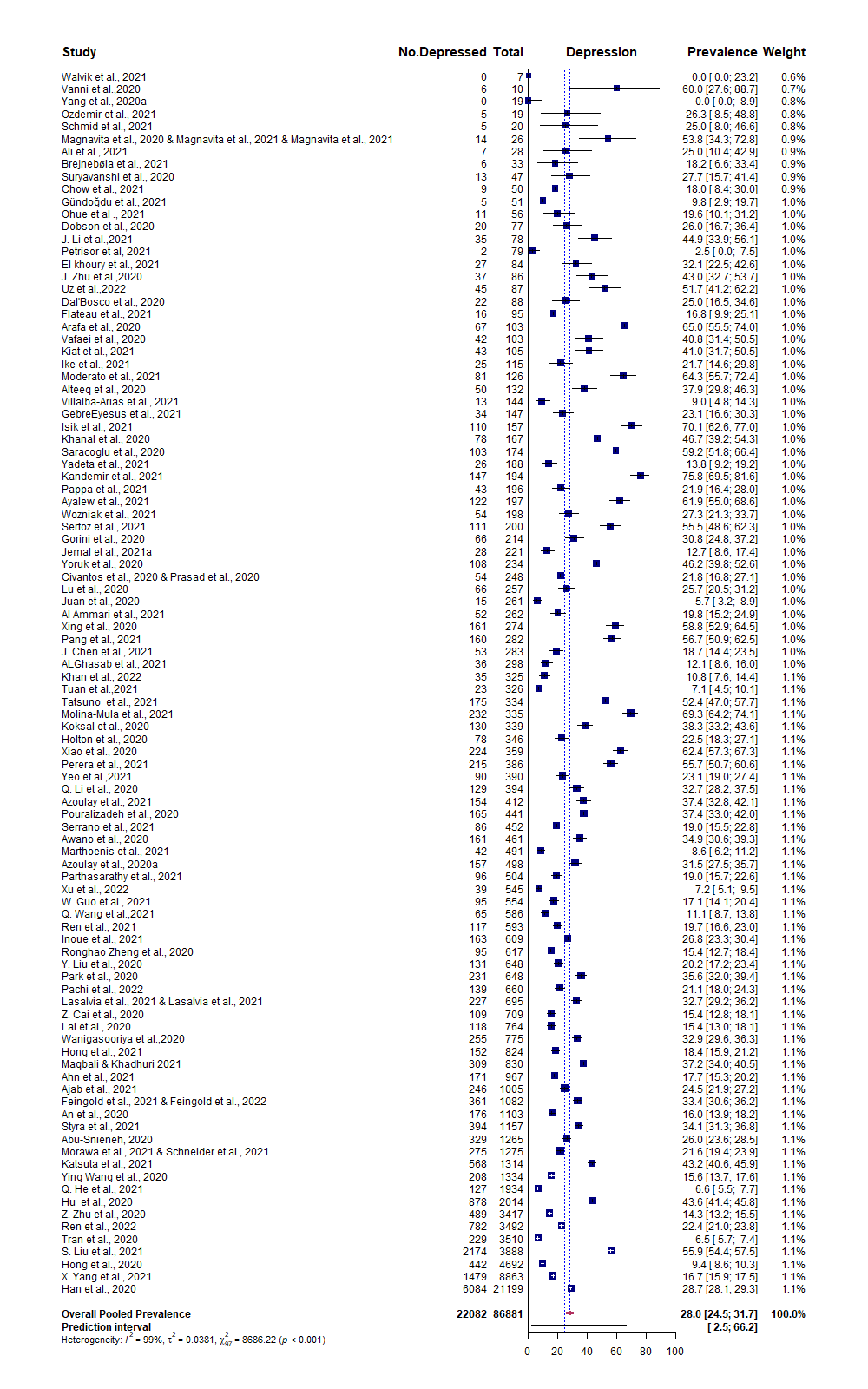

### S4D.Dep.Doc.tiff

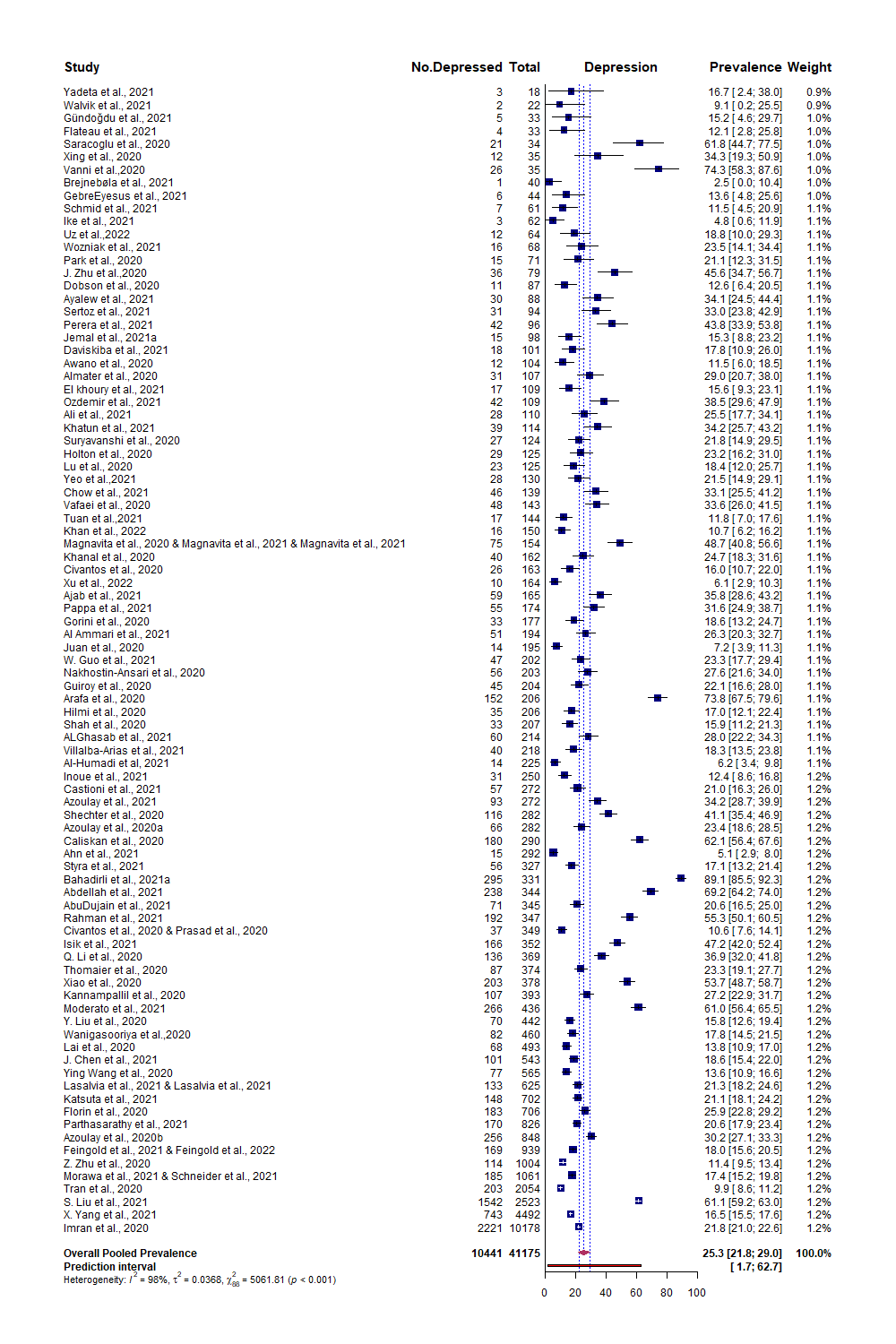

### S4E.Dep.ah.tiff

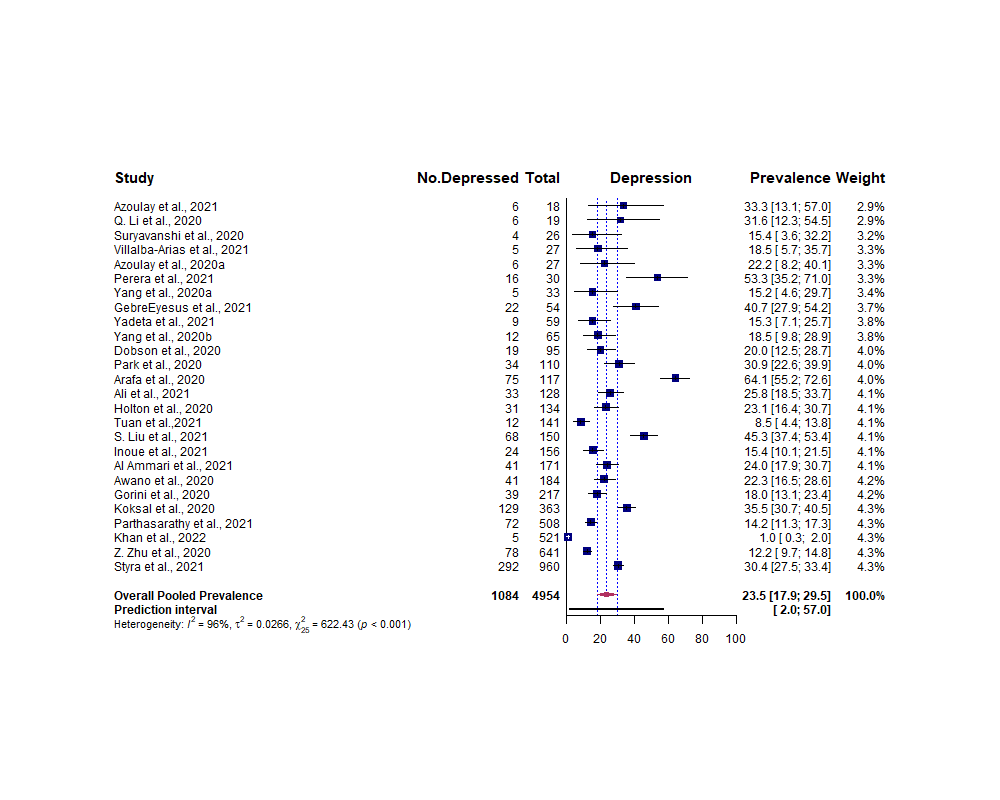

### S4F.Dep.nonmed.tiff

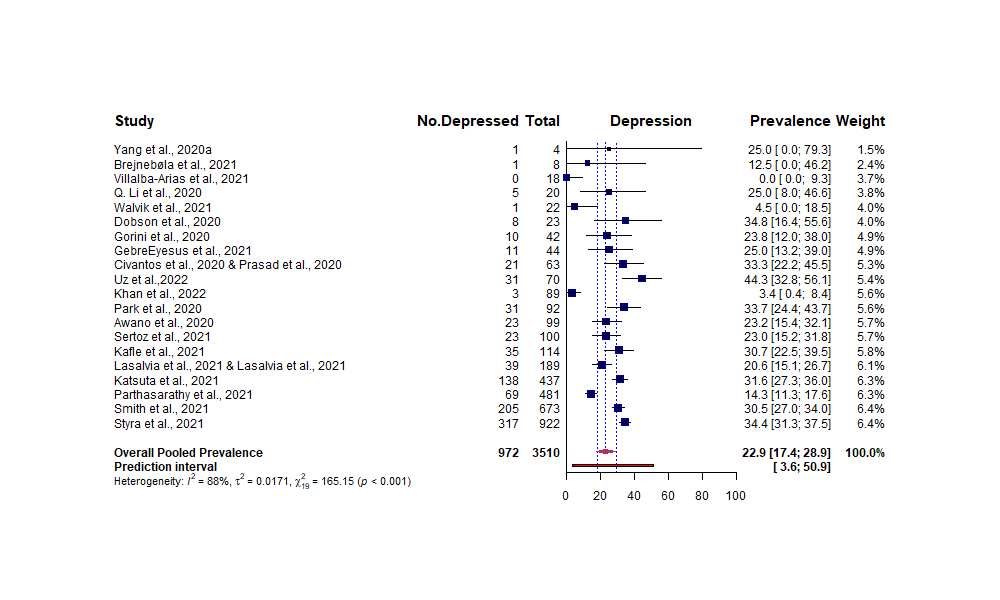

### S4G.Dep.Support.tiff

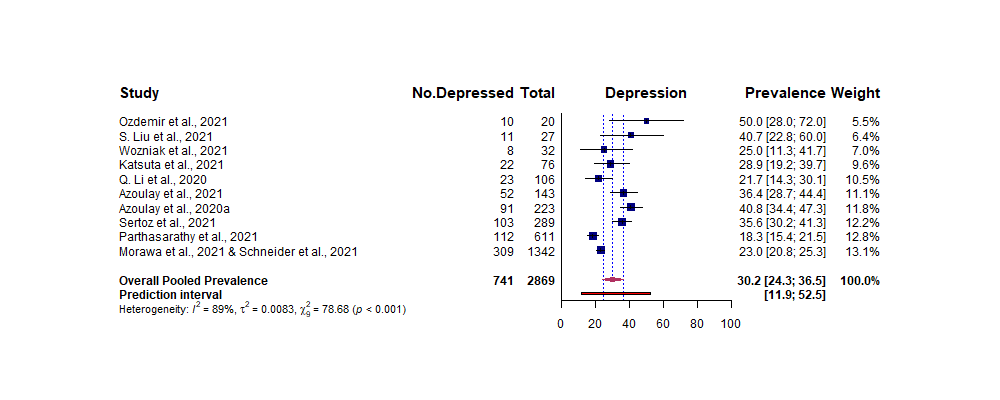

### S4H.Dep.student.tiff

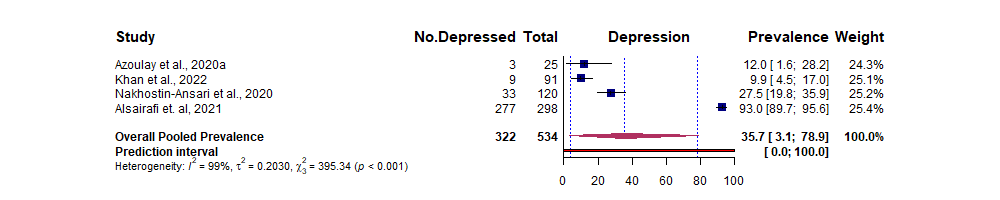

### S5A.Anx.Overall.tiff

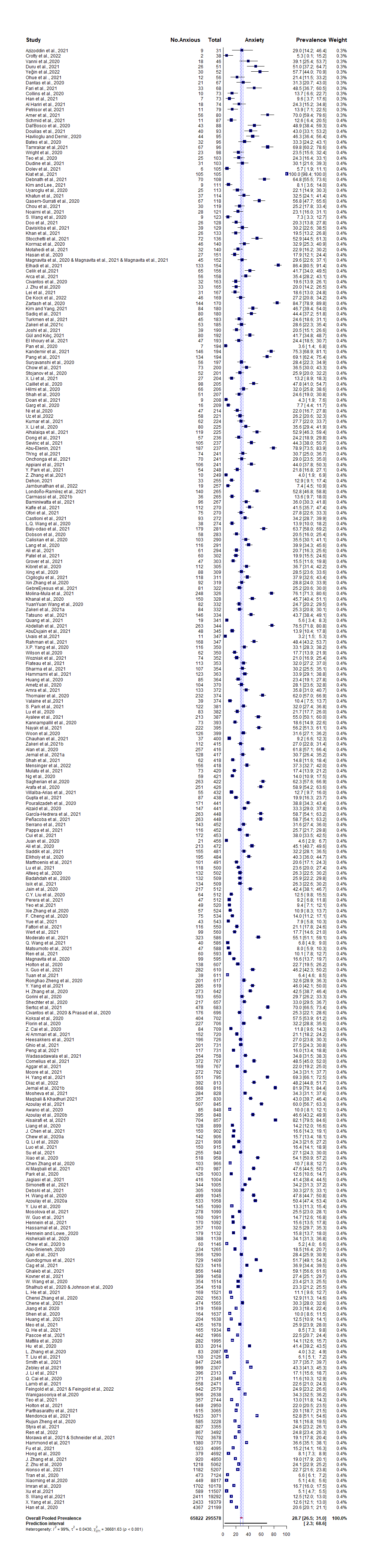

### S5B.Anx.Overall_by_Country.tiff

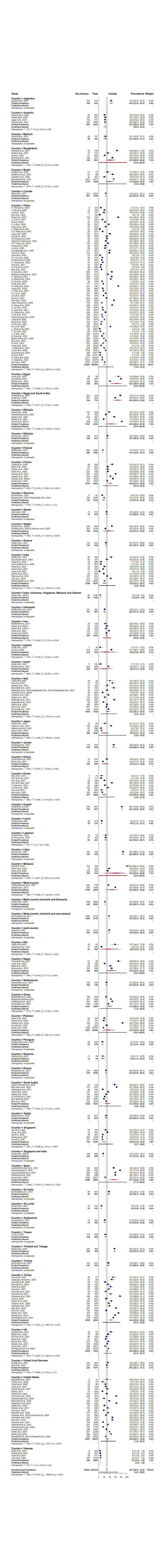

### S5C.Anx.Nurse.tiff

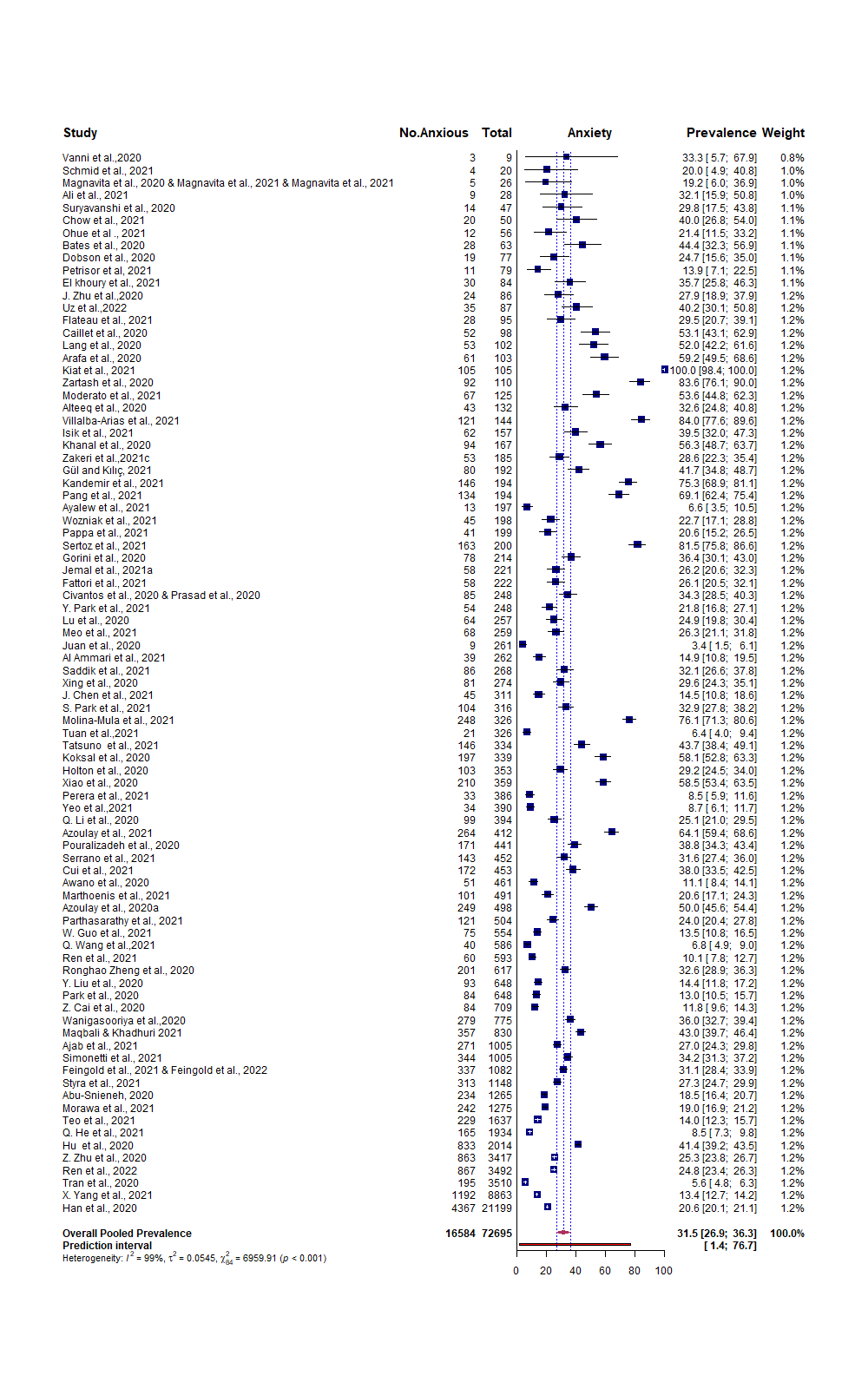

### S5D.Anx.Doc.tiff

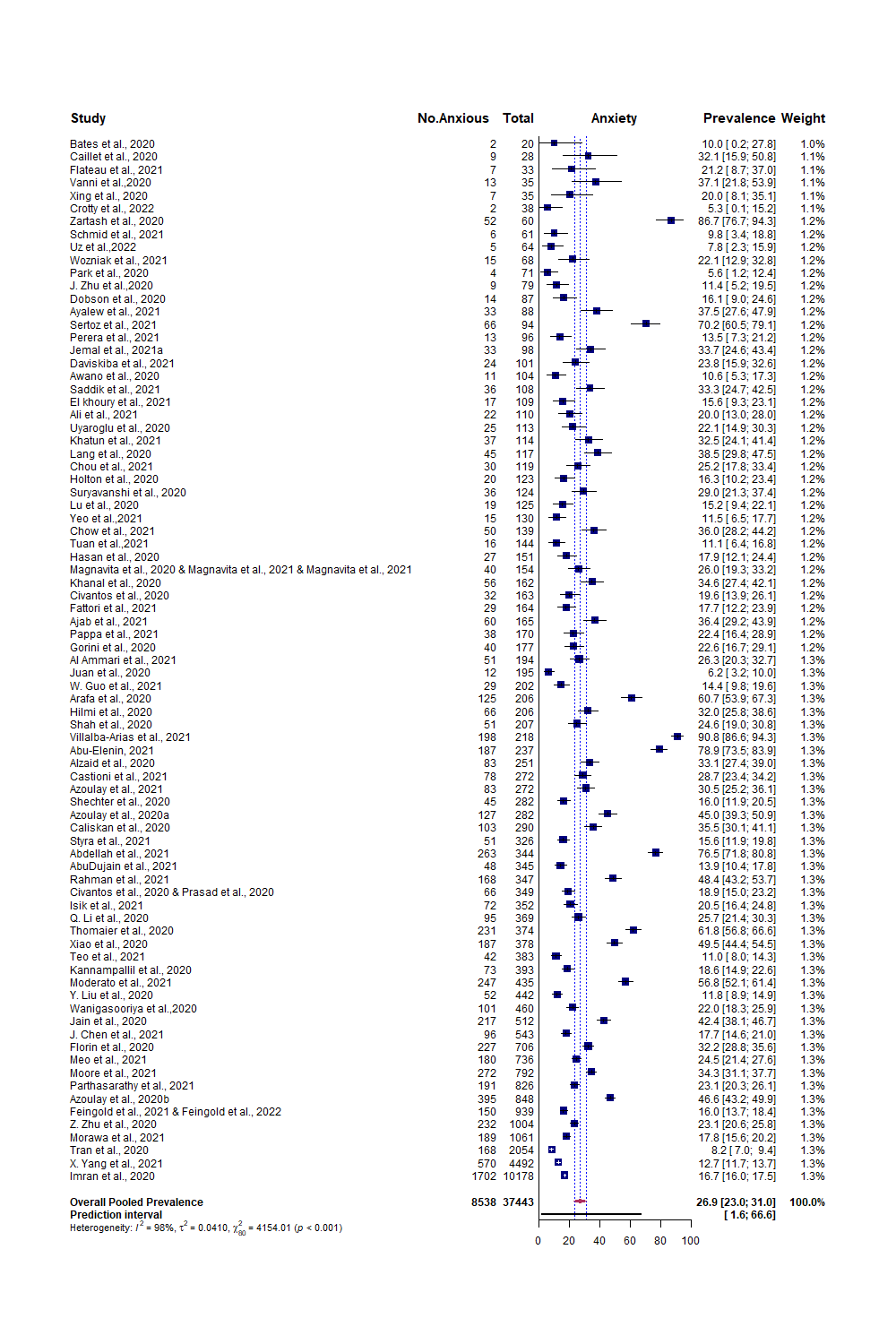

### S5E.Anx.ah.tiff

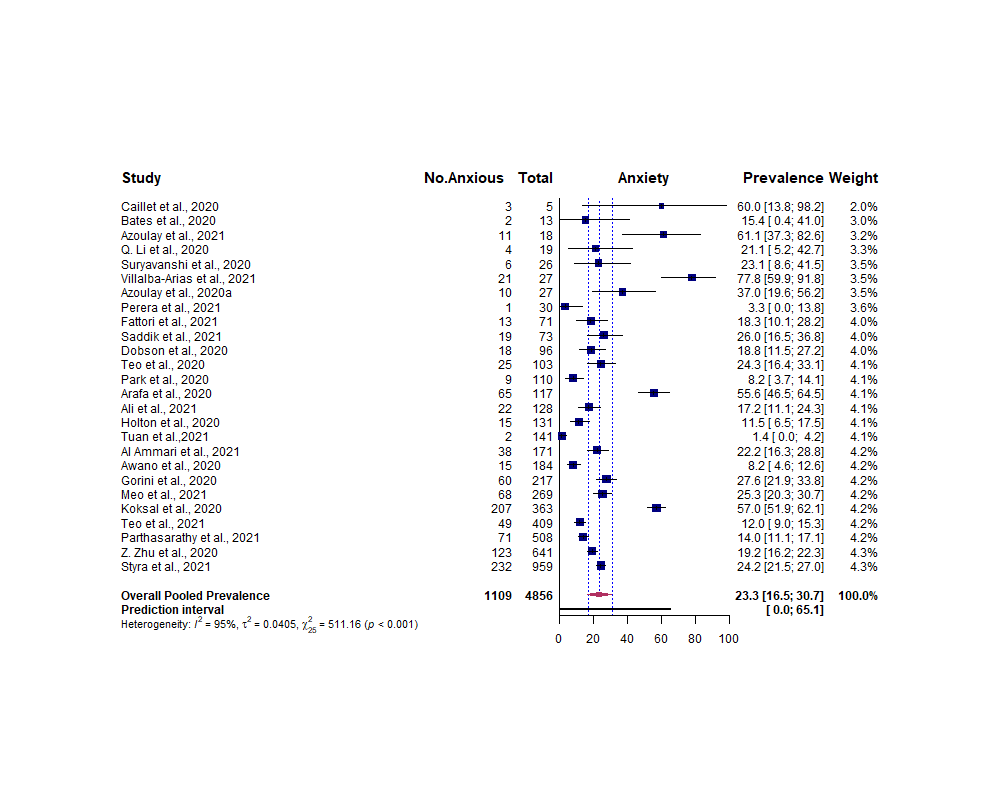

### S5F.Anx.nonmed.tiff

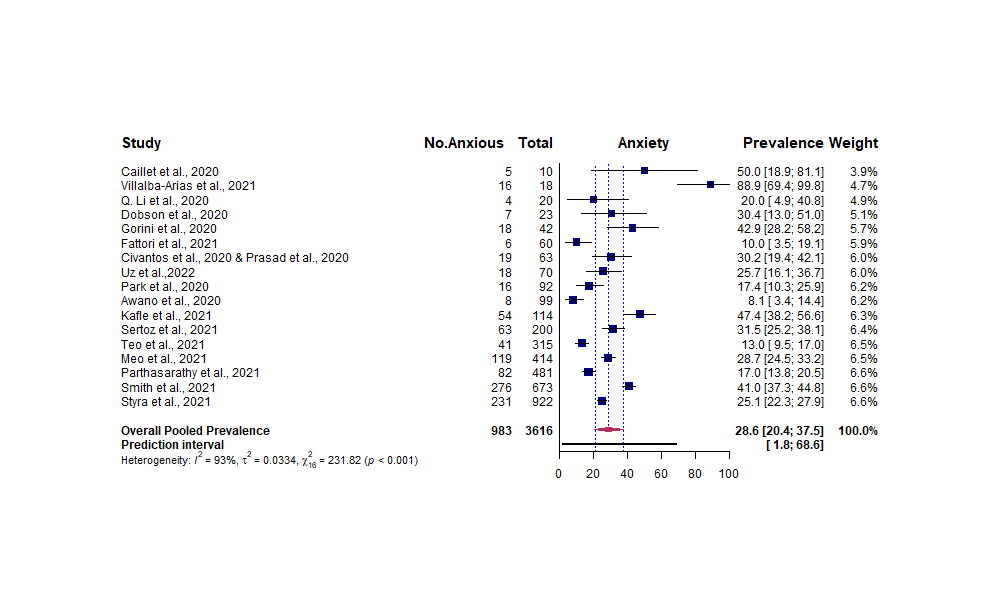

### S5G.Anx.support.tiff

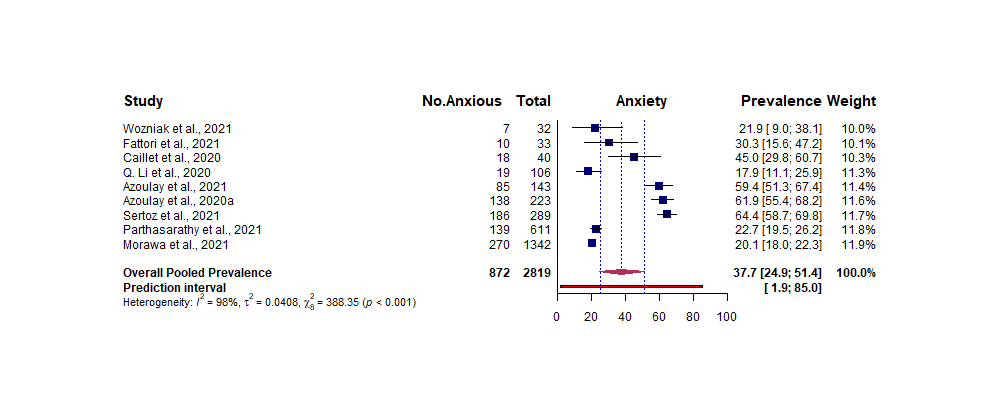

### S5H.Anx.student.tiff

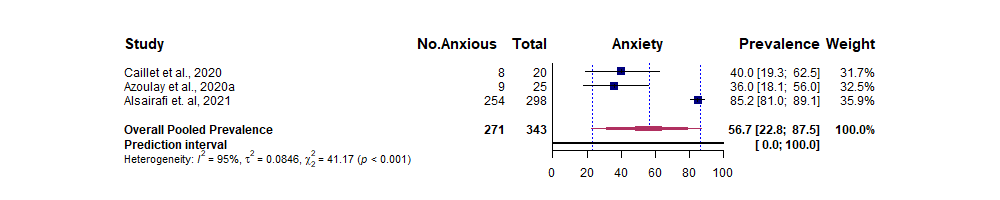

### S6A.Overall.tiff

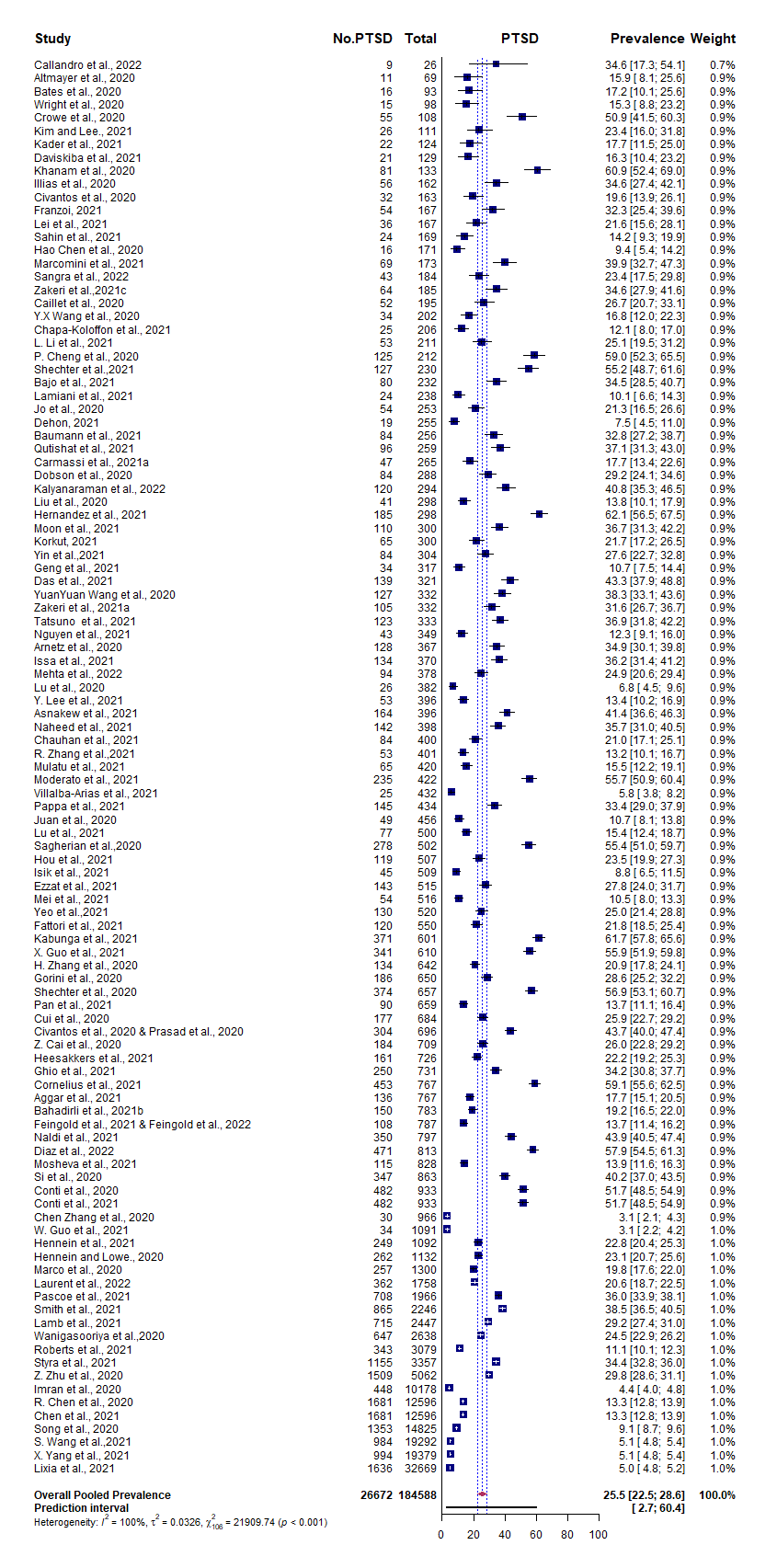

### S6B.Overall_by_Country.tiff

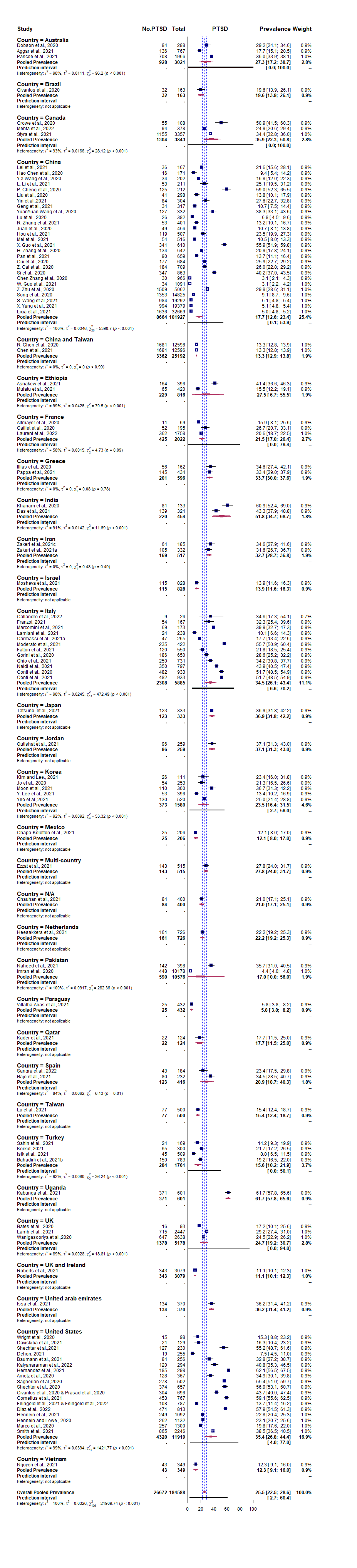

### S6C.Nurse.tiff

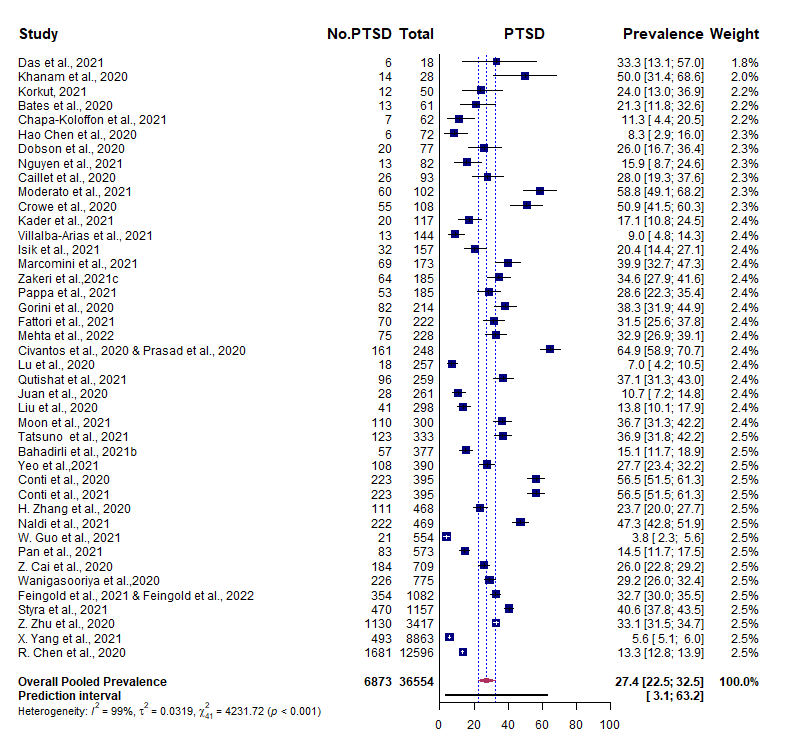

### S6D.Doc.tiff

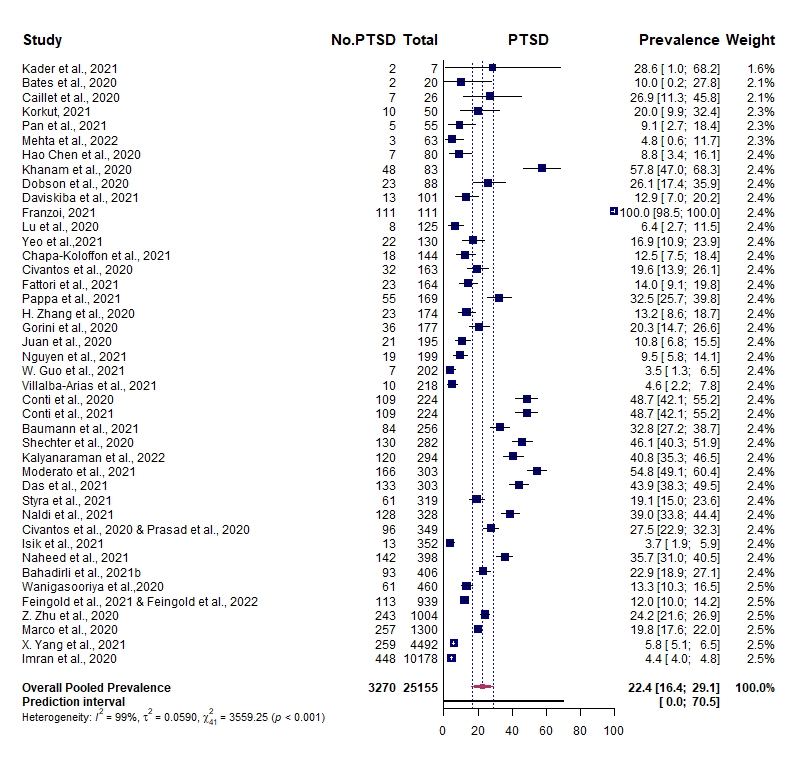

### S6E.ah.tiff

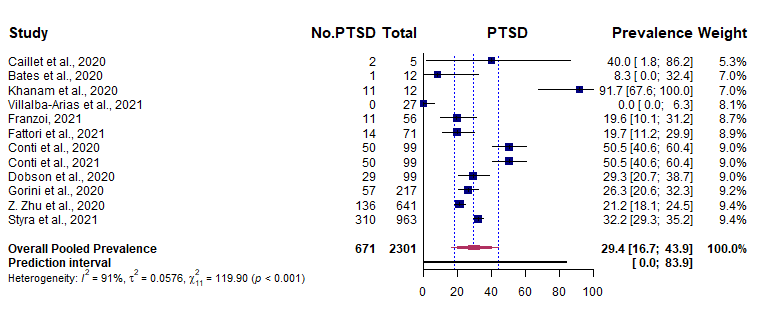

### S6F.nonmed.tiff

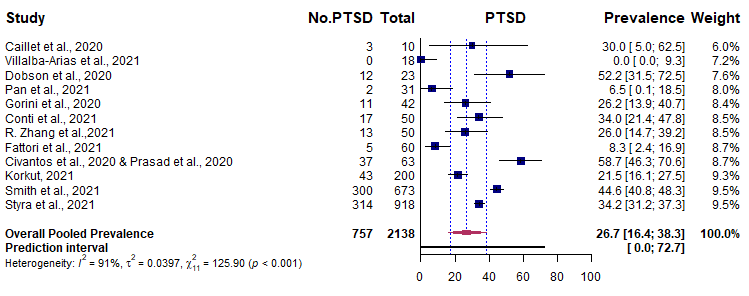

### S6G.support.tiff

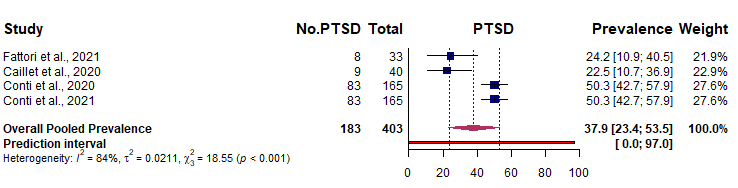

### S6H.student.tiff

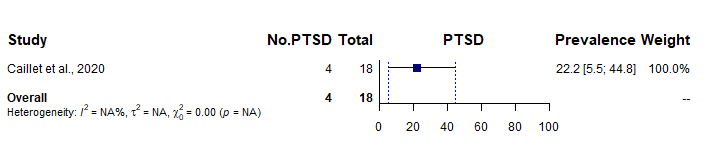

### S7A.Overall.tiff

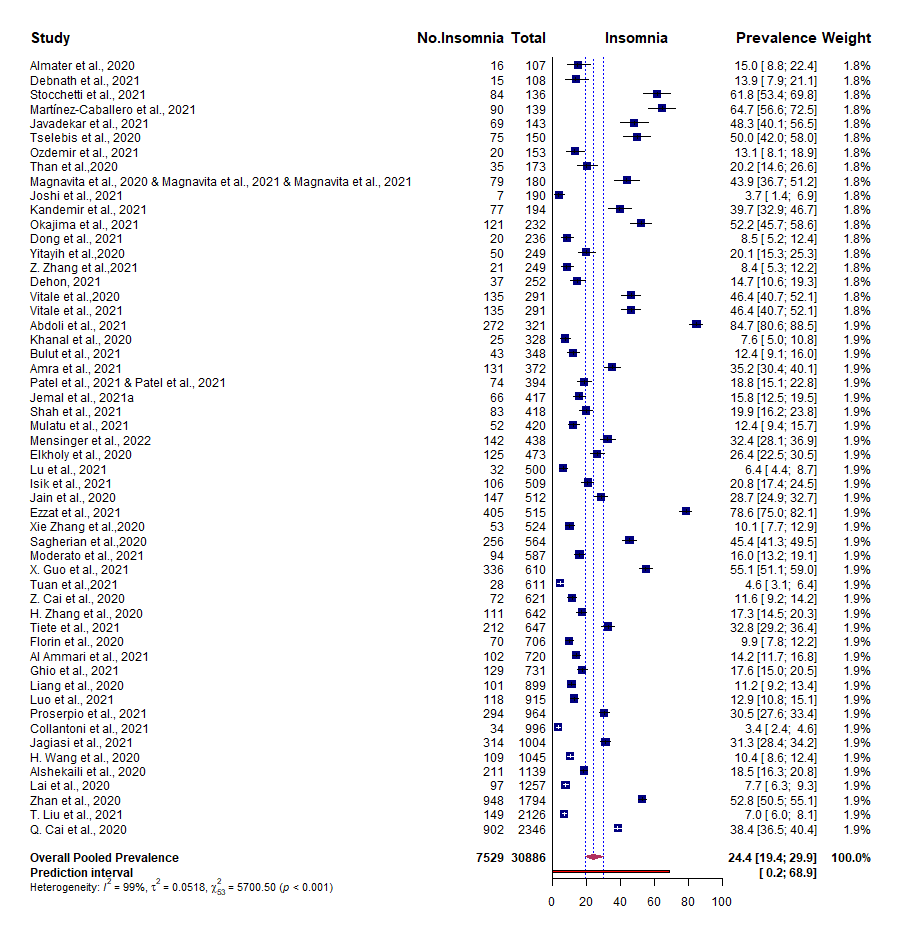

### S7B.Overall_by_Country.tiff

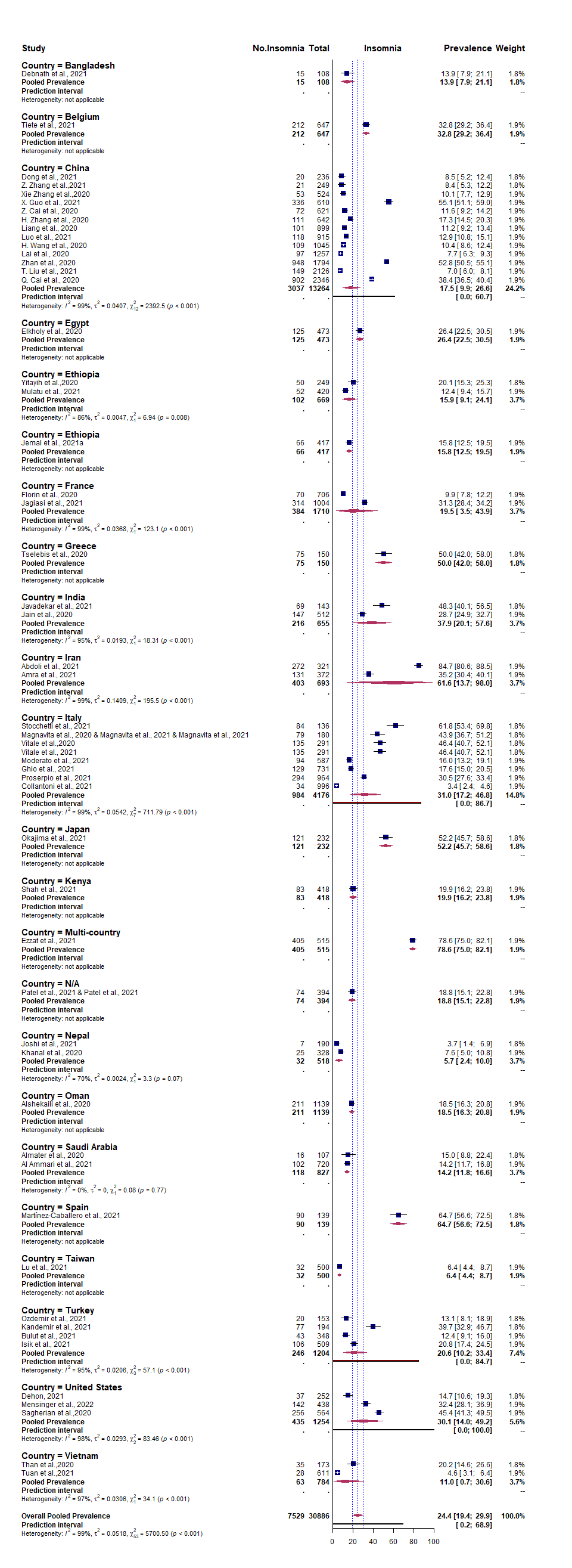

### S7C.Nurse.tiff

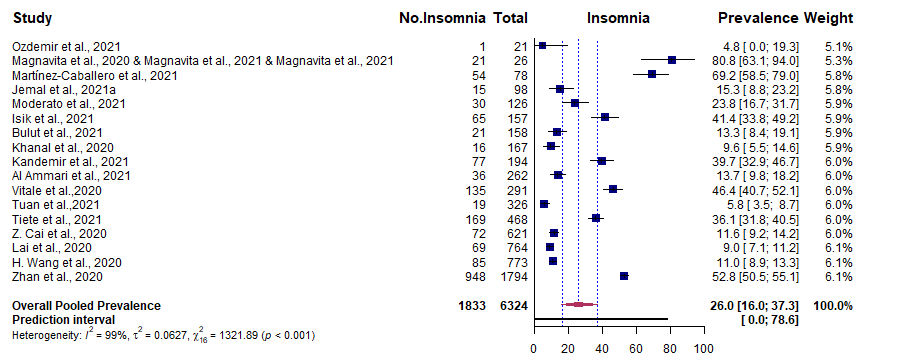

### S7D.Doc.tiff

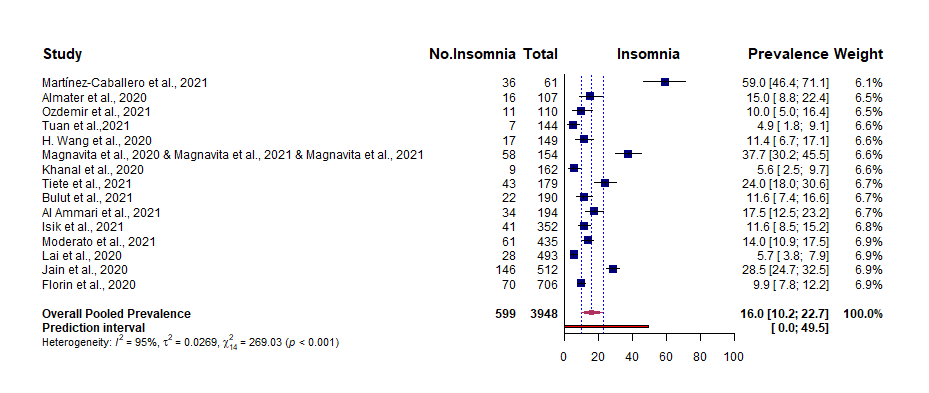

### S7E.ah.tiff

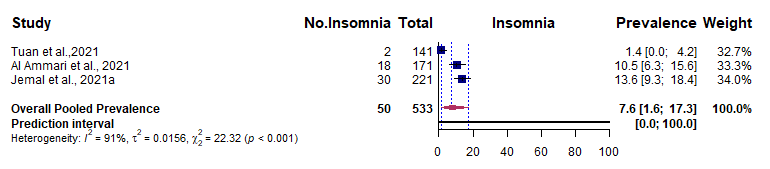

### S7F.support.tiff

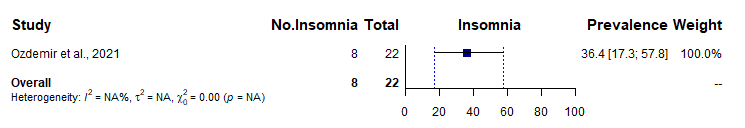

### S8A.Overall.tiff

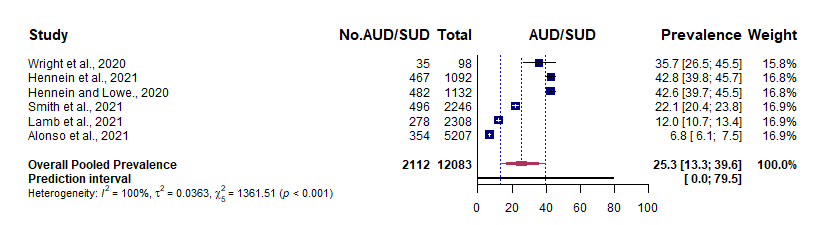

### S8B.Overall_by_Country.tiff

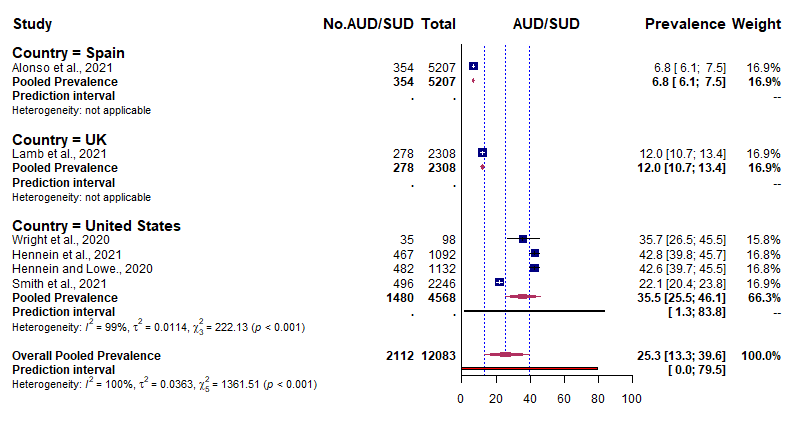
